# EHR-based Case Identification of Pediatric Long COVID: A Report from the RECOVER EHR Cohort

**DOI:** 10.1101/2024.05.23.24307492

**Authors:** Morgan Botdorf, Kimberley Dickinson, Vitaly Lorman, Hanieh Razzaghi, Nicole Marchesani, Suchitra Rao, Colin Rogerson, Miranda Higginbotham, Asuncion Mejias, Daria Salyakina, Deepika Thacker, Dima Dandachi, Dimitri A Christakis, Emily Taylor, Hayden Schwenk, Hiroki Morizono, Jonathan Cogen, Nathan M Pajor, Ravi Jhaveri, Christopher B. Forrest, L. Charles Bailey, the RECOVER Consortium

## Abstract

**Objective:** Long COVID, marked by persistent, recurring, or new symptoms post-COVID-19 infection, impacts children’s well-being yet lacks a unified clinical definition. This study evaluates the performance of an empirically derived Long COVID case identification algorithm, or computable phenotype, with manual chart review in a pediatric sample. This approach aims to facilitate large-scale research efforts to understand this condition better.

**Methods:** The algorithm, composed of diagnostic codes empirically associated with Long COVID, was applied to a cohort of pediatric patients with SARS-CoV-2 infection in the RECOVER PCORnet EHR database. The algorithm classified 31,781 patients with conclusive, probable, or possible Long COVID and 307,686 patients without evidence of Long COVID. A chart review was performed on a subset of patients (n=651) to determine the overlap between the two methods. Instances of discordance were reviewed to understand the reasons for differences.

**Results:** The sample comprised 651 pediatric patients (339 females, *M_age_* = 10.10 years) across 16 hospital systems. Results showed moderate overlap between phenotype and chart review Long COVID identification (accuracy = 0.62, PPV = 0.49, NPV = 0.75); however, there were also numerous cases of disagreement. No notable differences were found when the analyses were stratified by age at infection or era of infection. Further examination of the discordant cases revealed that the most common cause of disagreement was the clinician reviewers’ tendency to attribute Long COVID-like symptoms to prior medical conditions. The performance of the phenotype improved when prior medical conditions were considered (accuracy = 0.71, PPV = 0.65, NPV = 0.74).

**Conclusions:** Although there was moderate overlap between the two methods, the discrepancies between the two sources are likely attributed to the lack of consensus on a Long COVID clinical definition. It is essential to consider the strengths and limitations of each method when developing Long COVID classification algorithms.

## Introduction

Long COVID, also known as post-acute sequelae of SARS-CoV-2 infection (PASC), is a significant health concern characterized by ongoing, relapsing, or new symptoms emerging four or more weeks after the acute infection phase^1^. While post-viral syndromes like chronic fatigue syndrome following mononucleosis are well-documented in children^2–3^, understanding the clinical manifestations of Long COVID in pediatric patients remains incomplete. The variability of symptoms in children compared to adults complicates diagnosis and treatment^4–9^. Symptoms can range from fatigue and headache to loss of taste and smell and chest pain^4–9^. Although rare, diagnosed conditions associated with Long COVID include myocarditis, myositis, postural tachycardia syndrome (POTS), and myalgic encephalomyelitis/chronic fatigue syndrome (ME/CFS), among other conditions^10^. Despite certain symptoms and conditions clearly attributable to a SARS-CoV-2 infection, like multisystem inflammatory syndrome (MIS-C), much remains to be understood about others^11–12^. These symptoms and conditions impose a substantial burden on children and their families, leading to missed school and the need for service referrals^13–14^. This highlights the importance of improved detection and treatment strategies.

Identifying children who suffer from Long COVID in research studies is crucial to better understand this disorder and ensuring timely detection and treatments in clinical settings. However, this task is challenging due to the inconsistency and heterogeneity of associated symptoms. To address this challenge, researchers have used large observational cohort studies that use repositories of electronic health record (EHR) data to identify patients^5, 8, 9, 15, 16^. These studies have primarily relied on EHR-based diagnosis codes^15–16^. The ICD-10-CM U09.9 code, introduced in October 2021^17–18^, allows clinicians to assign a Long COVID diagnosis; however, its utilization remains inconsistent and potentially biased across patients and healthcare settings ^16^. Additionally, relying solely on this code may not adequately capture all patients due to the variety of symptoms associated with Long COVID. This poses a risk of misclassification if researchers exclusively use the U09.9 code for phenotyping.

To improve identification of patients with Long COVID, computable phenotyping techniques, which involve developing a set of rules to identify patients with a disorder, have been used in Long COVID studies. Long COVID phenotypes for adult^19^ and pediatric^20^ patients have been developed using machine-learning approaches that leverage large numbers of clinical features. For example, in a recent pediatric study, a machine learning algorithm demonstrated high precision in classifying both general and MIS-C-specific forms of PASC, with recall rates of up to 70% ^20^. Training these supervised learning models requires a labeled cohort of patients who likely have Long COVID based on healthcare utilization or Long COVID diagnosis codes. Since there is no gold-standard definition of Long COVID, it is difficult to produce an unbiased labeled training set, which limits the generalizability of the models.

In this study, we aimed to 1) identify children with Long COVID by utilizing a rules-based computable phenotype approach and 2) assess the performance of this computable phenotype for Long COVID in a subset of children. This approach involves analyzing specific diagnosis coding and symptoms that occur more frequently after a COVID-19 infection. By doing so, we can more accurately identify a larger number of children with Long COVID. In addition, we have included clinician reviews of patient charts to gain a comprehensive understanding of patients’ experience with Long COVID. This combined approach represents a significant step in the automation of Long COVID clinical phenotypes using EHR data in the absence of a consensus definition.

## Methods

### Data Source

This retrospective cohort study is part of the NIH Researching COVID to Enhance Recovery (RECOVER) Initiative, which seeks to understand, treat, and prevent the post-acute sequelae of SARS-CoV-2 infection^21^. The RECOVER PCORnet EHR cohort includes clinical data from patients in 40 hospital systems across the United States. Data were extracted from version 6 of the pediatric RECOVER database, comprising more than 9 million children who were tested for SARS-CoV-2, diagnosed with COVID-19, or received a COVID-19 vaccine between 2019 and December 2022. Institutional Review Board (IRB) approval was obtained under Biomedical Research Alliance of New York (BRANY) protocol #21-08-508. BRANY waived the need for consent and HIPAA authorization.

### Study Population

Inclusion criteria for our pediatric sample were as follows: 1) SARS-CoV-2 infection confirmed via clinical diagnosis or PCR, antigen, or qualifying serology test^22^ between March 2020 and December 2022, 2) age less than 21 years at first COVID-19 infection, and 3) at least two contacts with the healthcare system (at least one being in-person or telehealth) to ensure adequate follow-up during the post-acute phase (28-179 days following infection). We defined clinically meaningful time periods surrounding the initial COVID-19 infection as shown in Figure 1. The acute phase spanned until the 27th day post-infection. The post-acute phase, which was the primary focus of our analyses, spanned from day 28 through day 179 post-infection, ensuring that symptoms were not directly related to the acute COVID-19 infection. For patients with a specific COVID-19 diagnosis or viral test, the initial infection date was the date of diagnosis or test. For patients with diagnoses indicating “history of” or “complication of” COVID-19 or with a positive serology test, we used 28 days prior to the earliest diagnosis or test evidence of COVID-19 as a proxy for initial infection date.

**Figure 1.**
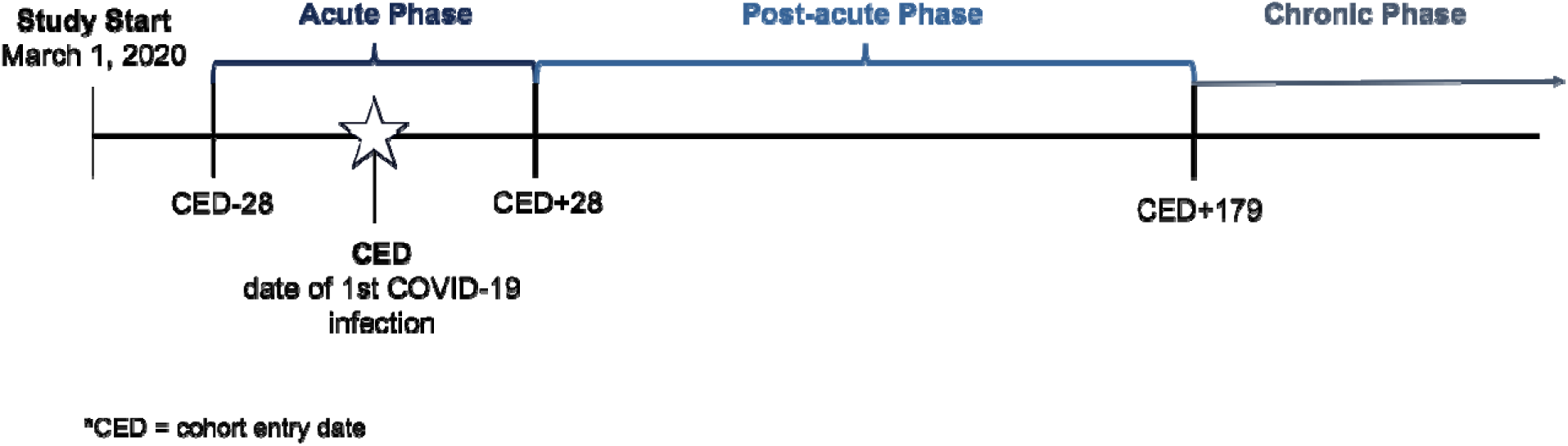
Study Timeline.

### Phenotype classification

Patients were identified as having conclusive, probable, or possible Long COVID according to the algorithm described in Figure 2, which used criteria documented in the EHR in the post-acute period. The algorithm accounts for diagnoses of Long COVID (ICD-10-CM code U09.9), diagnoses of MIS-C (ICD-10-CM code M35.81), diagnoses of sequelae of specified infectious and parasitic diseases (ICD-10-CM code B94.8), and 23 diagnosis clusters identified as probable indicators of Long COVID based on our prior work^5,9^ (Supplemental File 1). The diagnosis clusters were formed using a data mining approach that identified conditions more common in U09.9-diagnosed patients than in non-U09.9 diagnosed COVID-19+ patients in the post-acute period^5^. Clinicians then reviewed the diagnosis codes to create clusters of ICD-10-CM codes. Clusters included abdominal pain, abnormal liver enzymes, acute kidney injury, acute respiratory distress syndrome, arrythmias, autonomic dysfunction, cardiovascular signs/symptoms, changes in taste/smell, chest pain, cognitive function, generalized pain, fatigue/malaise, fever, fluid/electrolyte balance, headache, heart disease, myocarditis, musculoskeletal symptoms, myositis, respiratory signs/symptoms, and thrombophlebitis/thromboembolism. Any patient with two or more diagnoses within the same cluster separated by at least 28 days during the post-acute period was labeled as having probable Long COVID, regardless of whether the patient had a specific Long COVID or MIS-C diagnosis code. Figure 2 depicts the steps applied to classify patients according to the certainty of them having Long COVID. Any patient with conclusive, probable, or possible Long COVID detected by the phenotype was labeled as “Long COVID Evidence” and all others were labelled as “No Long COVID Evidence”.

**Figure 2.**
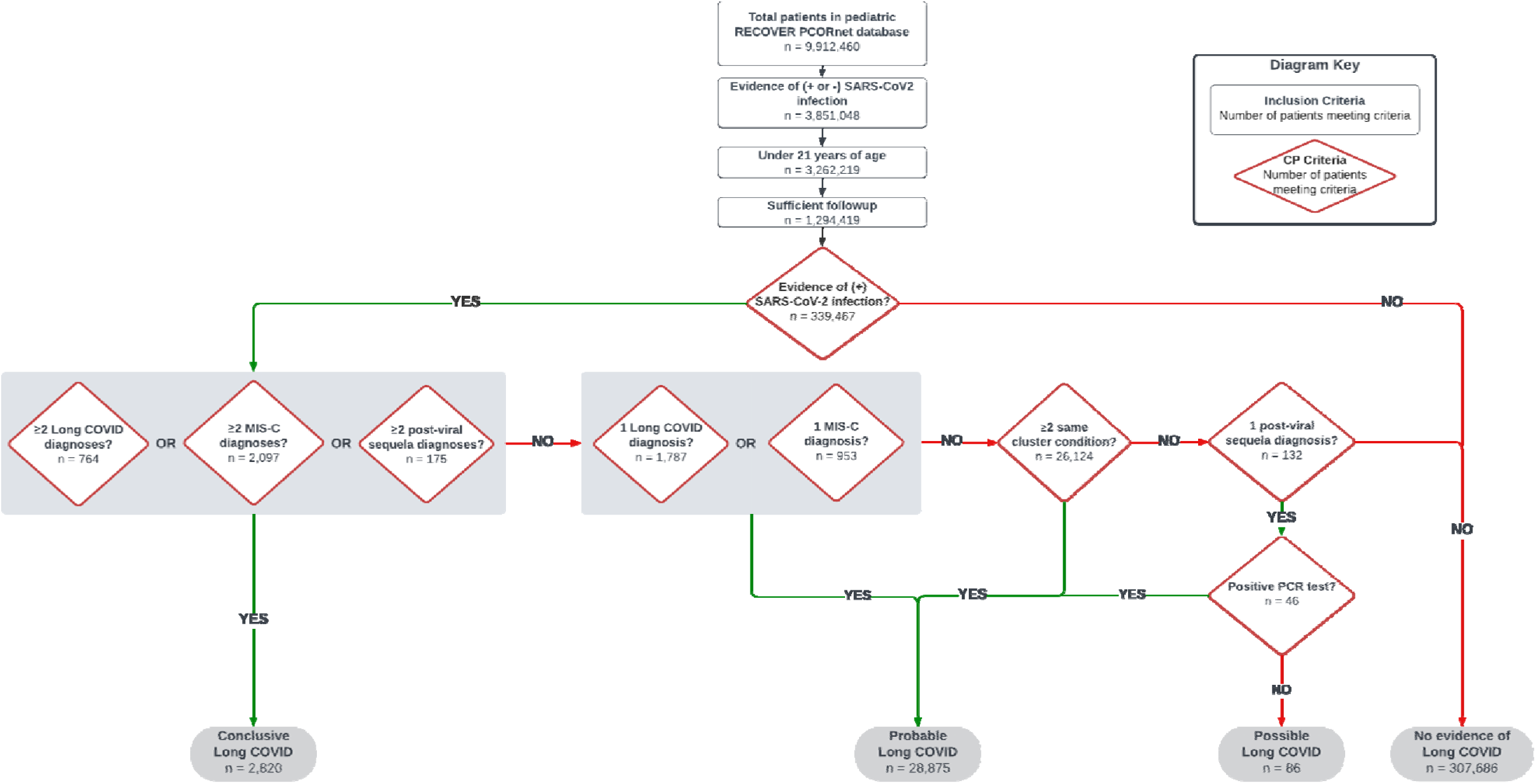
Flow chart depicting attrition and algorithm used to identify patients with Long COVID.

### Chart Review Sampling

A manual chart review was performed on a subset of the study population at 16 institutions. We sampled 702 patients split between the Long COVID Evidence and No Long COVID Evidence groups to ensure there was adequate representation across sites. The sampling strategy is laid out in Figure 3. Approximately 22 Long COVID Evidence patients were randomly sampled per institution. Each Long COVID Evidence sampled patient was matched 1:1 without replacement with a No Long COVID Evidence patient using exact matching on institution, age at time of infection, calendar quarter of infection, and acute period hospitalization (yes/no). Ninety percent of the No Long COVID Evidence sample had SARS-CoV-2 infection while the remaining ten percent (35 patients) were patients with no evidence of SARS-CoV-2 but with at least two diagnoses of cluster conditions separated by 28 to 150 days. The latter group were additional patients included in the chart review to gather insight on the attribution of cluster diagnoses to conditions other than SARS-CoV-2 infection. A total of 651 children were ultimately included in analyses based on additional exclusions which will be discussed. The sampling strategy is laid out in Figure 3.

**Figure 3.**
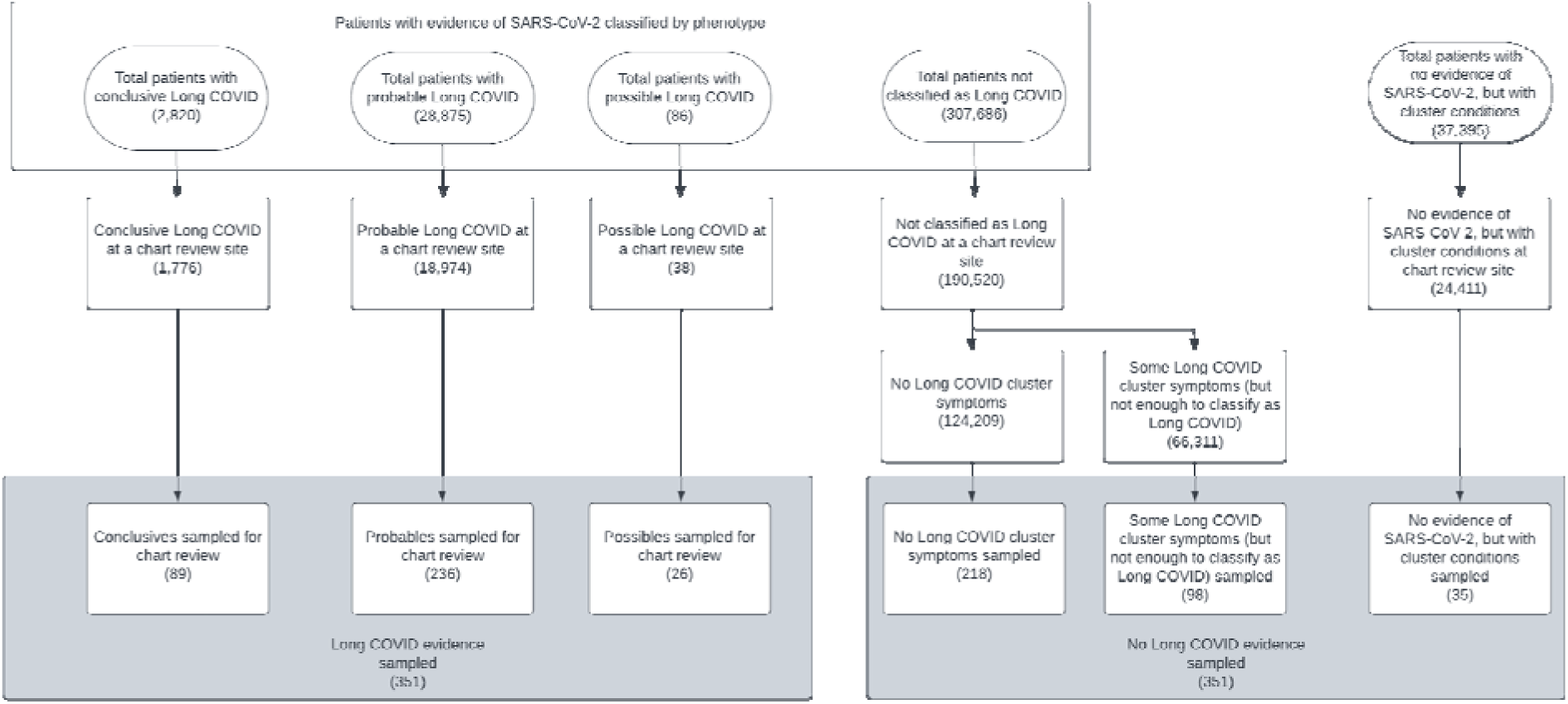
Sampling strategy for the chart review cohort. The numbers reported in parentheses represent sample sizes.

### Chart Review Procedure

Clinical research teams from each participating institution conducted chart reviews using a REDCap^23^ (Research Electronic Data Capture) instrument with questions including information on COVID-19 diagnoses and testing, demographics, COVID-19 prevention and treatment strategies, vaccines, functional outcomes, and conditions post COVID-19 captured in the patient’s medical record. Each site had between 1 and 5 reviewers for a total of 44 reviewers across sites. Table 1 contains a summary of patient information extracted from chart review, and the full case report form is included in Supplemental File 2.

**Table 1.** Demographics of children with and without Long COVID based on the computable phenotype definition.

|  | CP-Detected Long<br>COVID<br>(N=318) | No CP-Detected Long<br>COVID<br>(N=333) | Overall<br>(N=651) |
| --- | --- | --- | --- |
| *Approx. CED age (years)* |  |  |  |
| Mean (SD) | 10.10 (6.32) | 10.10 (6.28) | 10.10 (6.30) |
| Median [Min, Max] | 11.0 [0, 21.0] | 10.3 [0.1, 21.0] | 10.9 [0, 21.0] |
| *CED Age Group (years)* |  |  |  |
| <1 | 24 (7.5%) | 25 (7.5%) | 49 (7.5%) |
| 1-4 | 70 (22.0%) | 76 (22.8%) | 146 (22.4%) |
| 5-9 | 52 (16.4%) | 55 (16.5%) | 107 (16.4%) |
| 10-15 | 98 (30.8%) | 100 (30.0%) | 198 (30.4%) |
| 16-20 | 74 (23.3%) | 77 (23.1%) | 151 (23.2%) |
| *Patient Sex* |  |  |  |
| Male | 147 (46.2%) | 167 (50.2%) | 314 (48.2%) |
| Female | 171 (53.8%) | 166 (49.8%) | 337 (51.8%) |
| *Race* |  |  |  |
| Asian/Native<br>Hawaiian/Pacific<br>Islander | 12 (3.8%) | 14 (4.2%) | 26 (4.0%) |
| Black | 57 (17.9%) | 58 (17.4%) | 115 (17.7%) |
| White | 175 (55.0%) | 182 (54.7%) | 357 (54.8%) |
| Multiracial | 12 (3.8%) | 9 (2.7%) | 21 (3.2%) |
| Unknown | 62 (19.5%) | 70 (21.0%) | 132 (20.3%) |

| *Ethnicity* |  |  |  |
| --- | --- | --- | --- |
| Hispanic | 78 (24.5%) | 99 (29.7%) | 177 (27.2%) |
| Non-Hispanic | 214 (67.3%) | 210 (63.1%) | 424 (65.1%) |
| Unknown | 26 (8.2%) | 24 (7.2%) | 50 (7.7%) |
| *Payer\** |  |  |  |
| Private | 139 (43.7%) | 143 (42.9%) | 282 (43.3%) |
| Public | 124 (39.0%) | 134 (40.2%) | 258 (39.6%) |
| Other/Unknown | 55 (17.3%) | 56 (16.8%) | 111 (17.1%) |

*Note. CP = computable phenotype. \*=at time of COVID-19 infection.*

A secondary review was completed by a clinician who reviewed information extracted by the primary chart reviewer and answered questions regarding the level of confidence with which Long COVID could be assigned to the patient. The clinician was first asked if the patient met criteria for Long COVID based on the NIH definition^21^ which describes Long COVID as signs, symptoms, and conditions that continue or develop after initial COVID-19 or SARS-CoV-2 infection, are present four weeks [28 days] or more after the initial phase of infection; may be multisystemic; and may present with a relapsing-remitting pattern and progression or worsening over time, with the possibility of severe and life-threatening events even months or years after infection. They were then asked if the patient met criteria for Long COVID based on the computable phenotype definition. The response to these questions (i.e., conclusive, probable, possible, no evidence) was used to assess concordance with the computable phenotype. The first question, which analyses focused on, asked the clinician to exercise clinical judgment, while the second question was focused on assessing the validity of the structured EHR data.

For ease of assessing the performance of the computable phenotype compared with chart review, patients were collapsed into four overlapping groups: computable phenotype-positive (CP-positive), computable phenotype-negative (CP-negative), clinician review-positive (CR-positive), and clinician review-negative (CR-negative). Patients identified by the phenotype as having conclusive or probable Long COVID were placed in the CP-positive group. Conversely, patients found to have possible evidence or no evidence of Long COVID were placed in the CP- negative group. Patients with possible Long COVID were included in the CP-negative sample as the study team concluded that having only one post-viral sequelae code without a positive PCR test to confirm SARS-CoV-2 infection did not provide enough evidence to conclude that the patient’s post-viral sequelae was caused by Long COVID. On the other hand, when examining the chart review, we determined that the reasons reviewers used to classify patients as possible for Long COVID were more similar to a positive than a negative Long COVID classification. Therefore, the CR-positive group consisted of patients labeled as conclusive, probable, or possible for Long COVID by the clinician reviewer. In contrast, the CR-negative group consisted of patients labeled as having no evidence of Long COVID by the clinician reviewer.

### Performance Assessment

The performance of the computable phenotype was evaluated across various key metrics including sensitivity, specificity, positive predictive value (PPV), and negative predictive value (NPV). Additionally, we examined the accuracy and F1 score of the phenotype. Accuracy assesses the proportion of CR-positive patients who were also CP-positive. The F1 score combines precision and recall providing insight into the overall effectiveness of the phenotype.

Next, we assessed whether concordance between the phenotype and clinician review classification differed by age (i.e., under vs. over 12 years old), variant period (i.e., alpha, delta, omicron), and number of symptom clusters through stratified analyses. Analyses were conducted using R version 4.1.2 (2021-11-01; 24).

We conducted an assessment to identify discrepancies between the phenotype and chart review identification of Long COVID. We reviewed cases where the phenotype identified Long COVID, but the chart review did not, and vice versa. To understand the reasons behind these discrepancies, we reviewed the chart review form for each discordant patient, along with the clinician reviewer’s explanation for assigning or not assigning Long COVID. We then generated themes that accounted for the discrepancy and assigned those themes to the remaining cases. We next aimed to modify our model based the most common themes and perform a sensitivity analysis to assess whether the performance of our modified model was superior to our original model.

To describe the sample and investigate the impact of patients with complex medical histories on the performance of the phenotype, we used the Pediatric Medical Complexity Algorithm^25^ (PMCA). We determined the chronic condition status of each patient by applying the more conservative version of the algorithm. This version requires that a patient have one diagnosis of a progressive or malignant condition or at least two diagnoses per body system for at least two body systems in the three years prior to the SARS-CoV-2 infection.

## Results

Among patients with a positive SARS-CoV-2 infection (1,007,867), the computable phenotype detected Long COVID in 31,781 (3.2%) patients. Seven hundred and two patients were included in the chart review sample; however, sixteen charts were not completed due to limitations in the chart reviewers’ access to records and were excluded from the sample. In addition, the 35 patients with no evidence of SARS-CoV-2 infection according to the computable phenotype were not included in comparative analyses as a full chart review was not completed on them. Thus, our final sample consisted of 651 patients. Sample demographics and descriptive statistics are presented in Table 1. Sociodemographic characteristics were similar among those classified with and without Long COVID by the phenotype and by the clinician chart reviewer (Supplemental Table S1).

There was 73.33% agreement between the responses to the two questions the clinician chart reviewers answered. Table 2 presents statistics assessing the performance of the computable phenotype with chart review. The two methods had substantial but incomplete overlap. Analyses assessing whether concordance differed by selected variables showed similar results across age, era associated with infection, and number of symptom clusters (Supplemental Tables S2-4).

**Table 2.** Statistics comparing computable phenotype and clinician review identification of Long COVID.

|  | CR-Positive<br>(N = 239) | CR-Negative<br>(N = 412) |  |  |  |  |
| --- | --- | --- | --- | --- | --- | --- |
| CP-Positive (N = 318) | 156 | 162 |  |  |  |  |
| CP-Negative (N = 333) | 83 | 250 |  |  |  |  |
| Performance Statistics* |  |  |  |  |  |  |
|  | Accuracy | Sensitivity | Specificity | PPV | NPV | F1 |
|  | 0.624 | 0.653 | 0.607 | 0.491 | 0.751 | 0.560 |

*Note.* PPV = positive predictive value. NPV = negative predictive value. CP = computable phenotype. CR = chart review.
\*Reported as CP relative to CR.

### Computable Phenotype-Only and Clinician Review-Only Long COVID Positive Review

A review was conducted to assess the reasons for disagreement between the two methods. The initial focus was cases where the phenotype identified Long COVID, but the chart review did not (CP+/CR-cases) as there were many of these cases. Results are presented in Figure 4. In most CP+/CR- cases, the clinician reviewer agreed with the symptoms the computable phenotype identified but attributed those symptoms to another viral infection or preexisting disorder (Figure 4a). This was especially true for symptoms common to other respiratory infections and symptoms with occurrences both pre and post COVID-19 infection. In other less common cases, the reviewer did not see the diagnostic codes that the phenotype saw, or the reviewer made a conclusion based on incomplete information.

**Figure 4.**
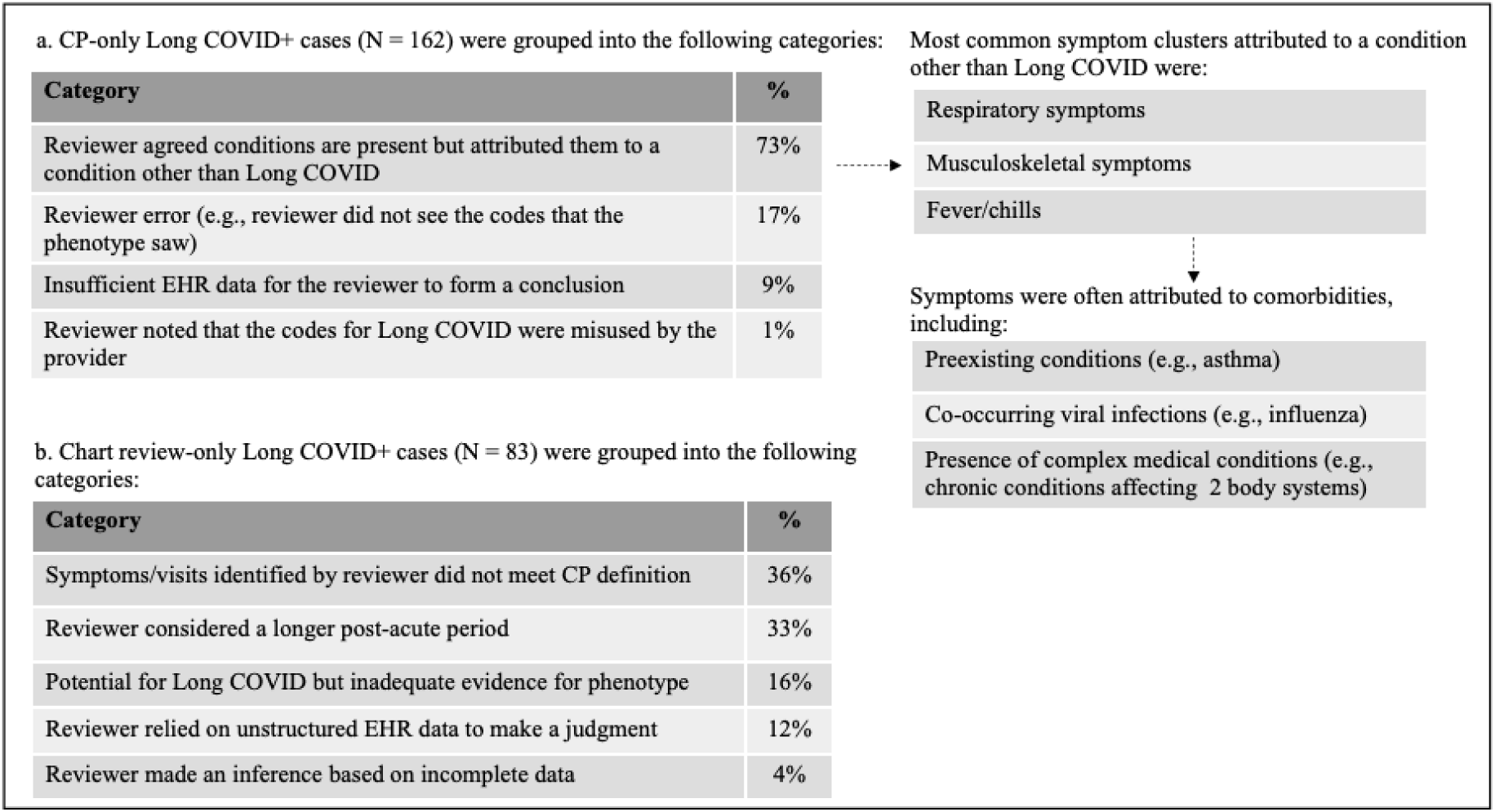
Qualitative review of a.) CP+/CR- Long COVID patients and b.) CR+/CP- Long COVID patients.

An assessment of CR+/CP- cases showed that in many cases the reviewer considered symptoms, visits, and time frames that differed from our phenotype (Figure 4b). For example, clinician reviewers considered symptoms beyond 180 days post-infection (up to 11 months in some cases) while the computable phenotype considered symptoms in the one month to 180 days following infection. In addition, reviewers considered symptoms beyond those included in the computable phenotype definition, such as mental health symptoms. For example, clinician reviewers used clinical judgment and identified anxiety as a qualifying symptom, but it is not included in the phenotype definition due to inconsistent recording in structured data.

### Sensitivity Analysis

The review of CP+/CR- patients showed that comorbidities were a large factor contributing to discordance between the two methods. Given the difficulty of distinguishing symptoms due to preexisting conditions and symptoms due to Long COVID, we performed a sensitivity analysis to assess concordance with a modified model in which preexisting conditions and comorbidities were accounted for. First, we censored prior symptoms. In other words, we assessed phenotype classification when non-incident diagnoses were excluded. Patients who met criteria for the computable phenotype with only preexisting symptom clusters that persisted after their COVID-19 diagnosis were labeled as having no evidence of Long COVID (n = 32 patients). Second, given the high prevalence of symptoms reported in the setting of non-COVID-19 respiratory infections, we excluded respiratory or fever cluster diagnoses that occurred 2 weeks prior to or after a diagnosis of a non-COVID-19 respiratory infection. This led to exclusion of 31 patients who did not otherwise meet the criteria for probable long COVID. Finally, it was difficult to attribute post-acute symptoms to a COVID-19 diagnosis versus underlying medical conditions in patients with multiple chronic medical conditions. Therefore, we identified patients with complex medical histories using the PMCA^25^. We reclassified patients with a complex chronic condition as having no evidence of Long COVID (n = 67 patients) as we believed that our phenotype could not accurately identify these patients as having Long COVID.

Concordance was assessed again after incorporating these alterations into a modified model, and results showed a higher positive predictive value and specificity but lower sensitivity (Table 3). The negative predictive value remained high. Given the difficulty in adequately attributing diagnoses to Long COVID in patients with complex medical histories, we also completed a sensitivity analysis where these patients were removed from the sample, and performance was reassessed (Table 3). Results showed similar performance to the modified model overall, but a higher F1 score.

**Table 3.**
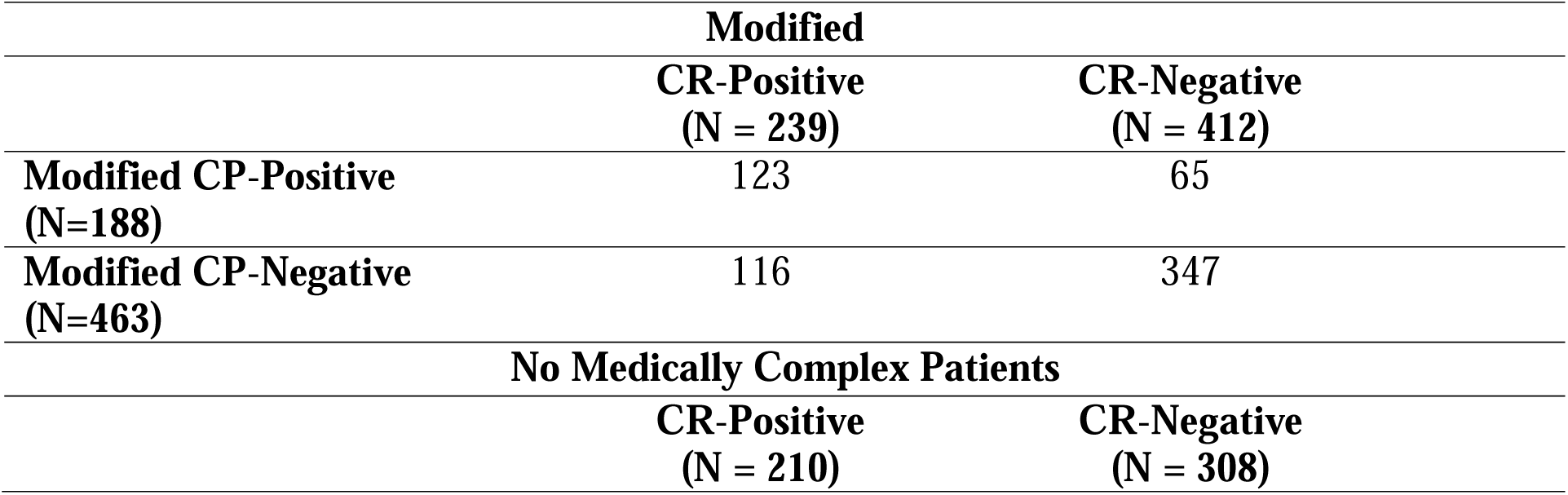
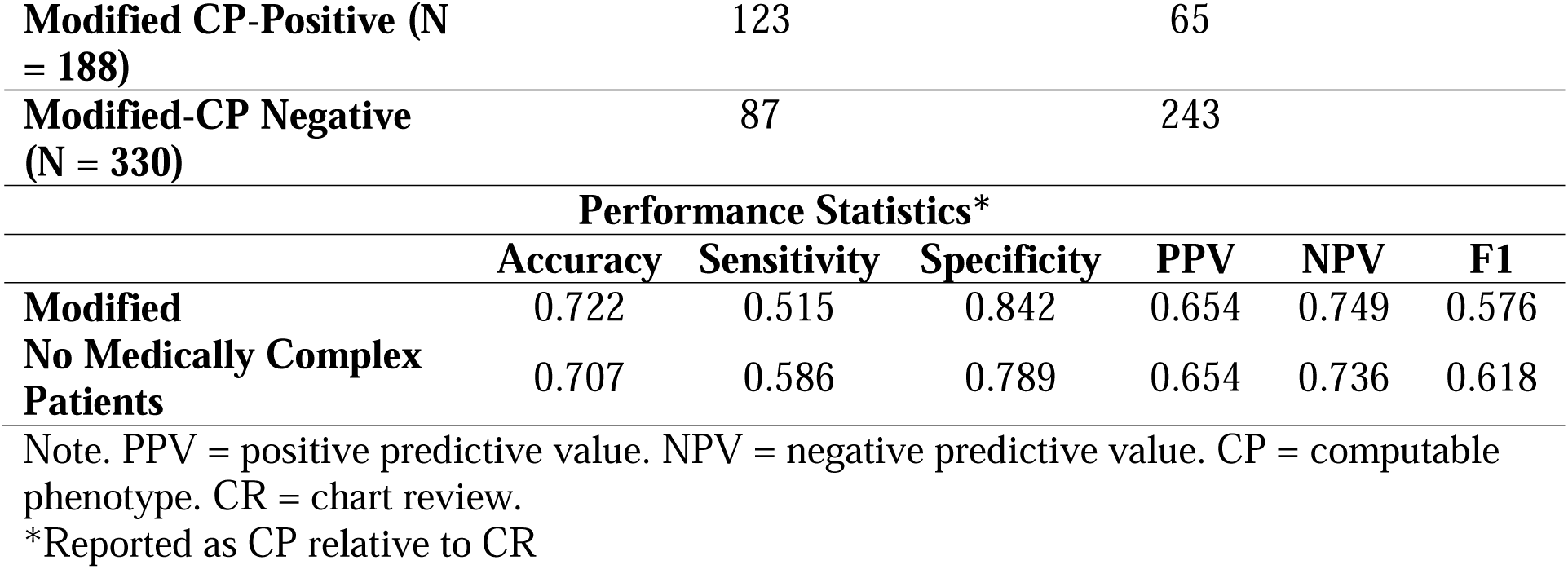
Statistics comparing modified computable phenotype and clinician review identification of Long COVID.

|  | **Modified** |  |
| --- | --- | --- |
|  | **CR-Positive**<br>**(N = 239)** | **CR-Negative**<br>**(N = 412)** |
| **Modified CP-Positive**<br>**(N=188)** | 123 | 65 |
| **Modified CP-Negative**<br>**(N=463)** | 116 | 347 |
| **No Medically Complex Patients** |  |  |
|  | **CR-Positive**<br>**(N = 210)** | **CR-Negative**<br>**(N = 308)** |

|  |  |  |  |  |  |  |
| --- | --- | --- | --- | --- | --- | --- |
| Modified CP-Positive (N = 188) | 123 | 65 |  |  |  |  |
| Modified-CP Negative (N = 330) | 87 | 243 |  |  |  |  |
| Performance Statistics* |  |  |  |  |  |  |
|  | Accuracy | Sensitivity | Specificity | PPV | NPV | F1 |
| Modified | 0.722 | 0.515 | 0.842 | 0.654 | 0.749 | 0.576 |
| No Medically Complex Patients | 0.707 | 0.586 | 0.789 | 0.654 | 0.736 | 0.618 |

Note. PPV = positive predictive value. NPV = negative predictive value. CP = computable phenotype. CR = chart review.
\*Reported as CP relative to CR

## Discussion

We conducted a study to assess the performance of a rules-based computable phenotype for identifying pediatric patients with Long COVID in a large EHR database compared to clinician chart review. Results showed moderate overlap between the two methods. Specifically, the computable phenotype was moderately sensitive in detecting patients with Long COVID and specific in detecting those without Long COVID, in comparison to chart review. However, there were several cases where the methods disagreed, with some patients being classified as having Long COVID by the phenotype but not by chart review, and vice versa. The main reason for these discrepancies was due to underlying comorbidities and subsequent respiratory infections.

Patients with comorbidities posed a challenge for the computable phenotype and the clinician reviewer. This was likely due to the lack of clinical guidelines for attribution and the difficulty in discerning exacerbation of preexisting symptoms. Clinicians were more likely to attribute post-COVID-19 symptoms to preexisting conditions when comorbidities were present, which likely resulted in the misattribution of Long COVID symptoms and may have been influenced by the provider involved in clinical care. However, the overlap between the two methods increased when the CP accounted for preexisting medical conditions by focusing on incident diagnoses and censoring existing conditions. Nevertheless, because our phenotype was not initially designed to assess exacerbation of preexisting conditions, we caution against its use to diagnose Long COVID in patients with medical complexity. Another source of disagreement between the computable phenotype and chart review stemmed from subsequent non-COVID-19 respiratory infections, which are common in children. Although there may be an increased risk of secondary infections due to SARS-CoV-2^26–27^, the symptoms are caused by a different agent. Therefore, we removed these circumstances as indicators of Long COVID in our phenotype.

An analysis of chart review-only positives (i.e., those the clinician reviewer classified as having Long COVID that the phenotype did not) showed that differences in the computable phenotype guidelines and the clinician’s framework for identifying Long COVID were the main reasons for discordance. While the computable phenotype only assessed symptoms up to 180 days post SARS-CoV-2 infection, clinicians may use a longer time window in practice. This suggests an extended time frame for assessing post-acute symptoms may be necessary, but also may increase the risk of later onset of symptoms not being clearly attributable to a SARS-CoV-2 infection. In addition, clinician reviewers identified conditions beyond those included in our phenotype as providing evidence for Long COVID. For example, the computable phenotype did not consider mental health conditions due to reporting inconsistency of these conditions using diagnosis codes and the difficulty in distinguishing between biologic and social causes of mental health conditions. However, clinicians tended to include them in their framework for identifying Long COVID. Therefore, constructing computable phenotypes that incorporate subphenotypes of interest (e.g., physiological vs psychological manifestations of Long COVID) may be useful in accounting for different manifestations of Long COVID.

Many of the differences in identifying Long COVID are due to the lack of a clear and consistently used definition for Long COVID. The novelty of Long COVID in children, as well as the overlap of symptoms with other acute and chronic disorders such as headache and fatigue, contribute to these differences. Similar difficulties have been encountered when defining other post-viral syndromes. Although chart review is often viewed as the best method for detecting patients with a specific disorder, the lack of a consistent definition for Long COVID by healthcare providers poses significant challenges. Moreover, the chart review and EHR research are prone to errors such as biases, missed codes, misdiagnoses, incomplete information due to fragmented care, and a lack of availability of a unique medical code for certain Long COVID-related conditions. For example, a unique ICD code for POTS did not exist until October 2022^28^, and many clinicians remain unaware of its existence, making it difficult to pick up the presence of POTS in the phenotype or chart review. Therefore, comparing our computable phenotype with chart review provided insight into the clinician’s view of a patient’s status, but it did not allow us to validate against a gold standard, as we cannot confidently conclude that either method accurately detects Long COVID.

Our two-pronged approach to identifying Long COVID using clinician chart review and a computable phenotype is a strength of the study as previous research that used diagnosis codes or machine learning algorithms did not incorporate a review of patient charts^19–20^. By incorporating both methods, we were able to qualitatively review cases of discordance. In addition, we focused on pediatric patients, in whom Long COVID is understudied. Research suggests that Long COVID has a lower prevalence in children; however, the current diagnostic tools may not be sensitive enough to detect all cases. Our study design is a strength as it uses data-driven symptom clusters for identifying Long COVID and specific diagnosis codes. This approach allowed us to capture patients who may not have a clear Long COVID presentation but have Long COVID-like symptoms. Although it allowed for our phenotype to be more inclusive, it also resulted in a computable phenotype that was less inclusive than a clinician may be when subjectively assessing whether a patient has Long COVID. As more symptoms of Long COVID are identified, it may be necessary to update the symptom clusters.

Our study has limitations, but it also brings attention to some significant areas for future work to focus on. Due to the challenge of distinguishing the progression of a chronic condition from symptom exacerbation due to COVID-19 using EHR data and chart review, we were unable to evaluate the worsening of preexisting symptoms. Instead, we examined differences in concordance after excluding pre-COVID-19 symptoms. This approach provided increased certainty that symptoms were due to Long COVID but may have been too restrictive. Future research should consider cluster-specific washout windows and develop reliable methods to identify patients with Long COVID-related worsening of preexisting conditions. Additionally, our sampling strategy focused on edge cases and rare occurrences to develop and refine the phenotype. This approach was useful for identifying patients with a range of Long COVID-related symptoms and diagnoses, but limits generalizability and underestimates the performance of the phenotype. Future iterations should use random sampling to obtain a more generalizable patient sample. Finally, because our study was based on EHR data and we imposed a two-visit requirement in the post-acute period, our sample may be biased towards patients who have the means to obtain healthcare at the population level.

## Conclusion

This study describes a computable phenotype approach to identify children with Long COVID in EHR data. Our study highlights the complexity of identifying and diagnosing Long COVID due to its heterogeneity and overlap with other conditions, which leads to substantial differences observed across methods. To address this challenge, future work could include additional data sources, such as unstructured data, and further refine algorithms with clinical expertise to develop a reliable definition of Long COVID. It is also essential to develop a revised phenotype that can identify Long COVID through the worsening of pre-existing conditions. The development of a reliable CP for Long COVID in children allows for studying large data networks, which has future applications for both observational studies and clinical trials. Further research assessing the presentation of Long COVID in children and the interplay between Long COVID and comorbidities is vital to continue to understand this emerging chronic illness and evaluate interventions that can prevent or mitigate its effects.

## Data Availability

All data produced in the present study are available upon reasonable request to the authors.

## Acknowledgements

This study is part of the NIH Researching COVID to Enhance Recovery (RECOVER) Initiative, which seeks to understand, treat, and prevent the post-acute sequelae of SARS-CoV-2 infection (PASC). For more information on RECOVER, visit https://recovercovid.org/

We would like to thank the National Community Engagement Group (NCEG), all patient, caregiver, and community Representatives, and all the participants enrolled in the RECOVER Initiative.

## Supporting Information

**Table S1.** Demographics of children with and without Long COVID based on the computable phenotype definition.

**Table S2.** Statistics comparing CP and chart review identification of Long COVID presented separately for patients younger and older than 12 years of age.

**Table S3.** Start and end dates associated with each era of infection.

**Table S4.** Statistics comparing CP and chart review identification of Long COVID presented separately by era of infection.

**Table S5.** Statistics comparing CP and chart review identification of Long COVID presented separately by the number of clusters identified by the CP.

