## Supplemental File 2 for "EHR-based Case Identification of Pediatric Long COVID: A Report from the RECOVER EHR Cohort"

*Data Dictionary Codebook*

### Round 2- RECOVER Pediatric PCORnet Chart Review Form (PID: 56148)

01/10/2024 5:18pm

**Instruments**

| # | Variable / Field Name | Field Label<br>*Field Note* | Field Attributes (Field Type, Validation, Choices, Calculations, etc.) |
|---|---|---|---|
| 1 | record_id | Record ID | text |
| 2 | chart_reviewer | Chart Reviewer Information:Please enter your first and last name; if the Coordinating Center has questions about the chart review, we may reach out with additional questions. | text, Required |
| 3 | approx_entry | Section Header: *Section 1. Inclusion/Exclusion Criteria*<br>Approximate Date of Cohort Entry: | text (date_mdy), Required<br>Field Annotation: @HIDEBUTTON |
| 4 | covid | Did this patient have COVID-19? | yesno, Required<br>1 Yes<br>0 No |
| 5 | coviddiagnosis_method<br><br>Show the field ONLY if:<br>[covid] = '1' | How was the diagnosis made? | dropdown, Required<br>1 Based on PCR result<br>2 Based on antigen result<br>3 Based on serology testing<br>4 Based on diagnosis code only<br>5 Self-reported by patient<br>6 Unsure/Not available |
| 6 | coviddiagnosis_date<br><br>Show the field ONLY if:<br>[covid] = '1' | Enter the date the first COVID-19 diagnosis was made: (Note: this date will be what you use as the Index Date in below sections) | text (date_mdy), Required<br>Field Annotation: @HIDEBUTTON |
| 7 | pcr_test_type<br><br>Show the field ONLY if:<br>[covid] = '1' and [coviddiagnosis_method] = '1' | Enter the type of PCR test: | dropdown, Required<br>1 Single PCR test for SARS-CoV-2<br>2 Multiplex PCR test which tested other respiratory pathogens in addition to SARS-CoV-2<br>3 Unknown/Unavailable |
| 8 | other_pathogen<br><br>Show the field ONLY if:<br>[covid] = '1' and [coviddiagnosis_method] = '1' | Did the patient have another respiratory pathogen identified within 7 days of the SARS CoV2 result? | dropdown, Required<br>1 Yes, same day<br>2 Yes, not on same day but within 7 days of test<br>3 No |

| | | | |
|---|---|---|---|
| 9 | respiratory_pathogens<br><br>Show the field ONLY if:<br>[other_pathogen] = '1' or [other_pathogen] = '2' | If yes, include name(s) of other respiratory pathogen (select all that apply from the following): | checkbox, Required<br>1 respiratory_pathogens___1 Adenovirus<br>2 respiratory_pathogens___2 Seasonal Coro[illegible] (229E, HKU1, N[illegible] OC43)<br>3 respiratory_pathogens___3 Human Metapneumov[illegible]<br>4 respiratory_pathogens___4 Human Rhinovirus/Ent[illegible]<br>5 respiratory_pathogens___5 Parainfluenza [illegible] 1,2,3 or 4<br>6 respiratory_pathogens___6 Respiratory sy[illegible] virus<br>7 respiratory_pathogens___7 Bacteria (Bord[illegible] parapertussis, Bordetella pert[illegible] Chlamydia pneumoniae o[illegible] Mycoplasma pneumoniae)<br>8 respiratory_pathogens___8 Unknown/Una[illegible] |
| 10 | coviddiagnosis_age<br><br>Show the field ONLY if:<br>[covid] = '1' | Select the patient's age range at first COVID-19 diagnosis: | dropdown, Required<br>1 Under 3 years old<br>2 3 years or older |
| 11 | coviddiagnosis_multiple<br><br>Show the field ONLY if:<br>[covid] = '1' | Did the patient have repeated COVID-19 (more than one SARS-CoV-2 infection)? | yesno, Required<br>1 Yes<br>0 No |
| 12 | coviddiagnosis_method_2<br><br>Show the field ONLY if:<br>[covid] = '1' and [coviddiagnosis_multiple] = '1' | How was the second diagnosis made? | dropdown, Required<br>1 Based on PCR result<br>2 Based on antigen result<br>3 Based on serology testing<br>4 Based on diagnosis code only<br>5 Self-reported by patient<br>6 Unsure/Not available |
| 13 | coviddiagnosis_date_2<br><br>Show the field ONLY if:<br>[covid] = '1' and [coviddiagnosis_multiple] = '1' | Enter the date the second COVID-19 diagnosis was made: | text (date_mdy), Required<br>Field Annotation: @HIDEBUTTON |
| 14 | pcr_test_type_2<br><br>Show the field ONLY if: | Enter the type of PCR test: | dropdown, Required<br>1 Single PCR test for SARS-CoV-2 |

| | | | |
|---|---|---|---|
| | [covid] = '1' and [coviddiagnosis_method_2] = '1' | | 2 Multiplex PCR test which tested other respiratory pathogens in addition to SARS-CoV-2<br>3 Unknown/Unavailable |
| 15 | other_pathogen_2<br>Show the field ONLY if:<br>[covid] = '1' and [coviddiagnosis_method_2] = '1' | Did the patient have another respiratory pathogen identified within 7 days of the SARS CoV2 result? | dropdown, Required<br>1 Yes, same day<br>2 Yes, not on same day but within 7 days of test<br>3 No |
| 16 | respiratory_pathogens_2<br>Show the field ONLY if:<br>[other_pathogen_2] = '1' or [other_pathogen_2] = '2' | If yes, include name(s) of other respiratory pathogen (select all that apply from the following): | checkbox, Required<br>1 respiratory_pathogens_2___1 Adenovirus<br>2 respiratory_pathogens_2___2 Seasonal Co[illegible] (229E, HKU1, [illegible] OC43)<br>3 respiratory_pathogens_2___3 Human Metapneum[illegible]<br>4 respiratory_pathogens_2___4 Human Rhinovirus/E[illegible]<br>5 respiratory_pathogens_2___5 Parainfluenz[illegible] 1,2,3 or 4<br>6 respiratory_pathogens_2___6 Respiratory s[illegible] virus<br>7 respiratory_pathogens_2___7 Bacteria (Bor[illegible] parapertussi[illegible] Bordetella pe[illegible] Chlamydia pneumoniae[illegible] Mycoplasma pneumoniae[illegible]<br>8 respiratory_pathogens_2___8 Unknown/Ur[illegible] |
| 17 | coviddiagnosis_method_3<br>Show the field ONLY if:<br>[covid] = '1' and [coviddiagnosis_multiple] = '1' | How was the third diagnosis made? | dropdown<br>1 Based on PCR result<br>2 Based on antigen result<br>3 Based on serology testing<br>4 Based on diagnosis code only<br>5 Self-reported by patient<br>6 Unsure/Not available |
| 18 | coviddiagnosis_date_3<br>Show the field ONLY if:<br>[covid] = '1' and [coviddiagnosis_multiple] = '1' | Enter the date the third COVID-19 diagnosis was made: | text (date_mdy)<br>Field Annotation: @HIDEBUTTON |

| | | | |
|---|---|---|---|
| 19 | pcr_test_type_3<br>Show the field ONLY if:<br>[covid] = '1' and [coviddiagnosis_method_3] = '1' | Enter the type of PCR test: | dropdown, Required<br>1 \| Single PCR test for SARS-CoV-2<br>2 \| Multiplex PCR test which tested other respiratory pathogens in addition to SARS-CoV-2<br>3 \| Unknown/Unavailable |
| 20 | other_pathogen_3<br>Show the field ONLY if:<br>[covid] = '1' and [coviddiagnosis_method_3] = '1' | Did the patient have another respiratory pathogen identified within 7 days of the SARS CoV2 result? | dropdown, Required<br>1 \| Yes, same day<br>2 \| Yes, not on same day but within 7 days of test<br>3 \| No |
| 21 | respiratory_pathogens_3<br>Show the field ONLY if:<br>[other_pathogen_3] = '1' or [other_pathogen_3] = '2' | If yes, include name(s) of other respiratory pathogen (select all that apply from the following): | checkbox, Required<br>1 \| respiratory_pathogens_3___1 \| Adenovirus<br>2 \| respiratory_pathogens_3___2 \| Seasonal Cor[illegible] (229E, HKU1, [illegible] OC43)<br>3 \| respiratory_pathogens_3___3 \| Human Metapneum[illegible]<br>4 \| respiratory_pathogens_3___4 \| Human Rhinovirus/E[illegible]<br>5 \| respiratory_pathogens_3___5 \| Parainfluenz[illegible] 1,2,3 or 4<br>6 \| respiratory_pathogens_3___6 \| Respiratory s[illegible] virus<br>7 \| respiratory_pathogens_3___7 \| Bacteria (Bor[illegible] parapertussi[illegible] Bordetella pe[illegible] Chlamydia pneumoniae [illegible] Mycoplasma pneumoniae[illegible]<br>8 \| respiratory_pathogens_3___8 \| Unknown/Ur[illegible] |
| 22 | covid_multiplereinfections<br>Show the field ONLY if:<br>[covid] = '1' and [coviddiagnosis_multiple] = '1' | Did the patient have more than three repeated COVID-19 infections? | yesno, Required<br>1 \| Yes<br>0 \| No |
| 23 | coviddiagnosis_method_4<br>Show the field ONLY if:<br>[covid] = '1' and [coviddiagnosis_multiple] = '1' and [covid_multiplereinfections] = '1' | How was the fourth diagnosis made? | dropdown, Required<br>1 \| Based on PCR result<br>2 \| Based on antigen result<br>3 \| Based on serology testing<br>4 \| Based on diagnosis code only<br>5 \| Self-reported by patient |

| | | | |
|---|---|---|---|
| | | | 6 Unsure/Not available |
| 24 | coviddiagnosis_date_4<br>Show the field ONLY if:<br>[covid] = '1' and [coviddiagnosis_multiple] = '1' and [covid_multiplereinfections] = '1' | Enter the date the fourth COVID-19 diagnosis was made: | text (date_mdy), Required<br>Field Annotation: @HIDEBUTTON |
| 25 | pcr_test_type_4<br>Show the field ONLY if:<br>[covid] = '1' and [coviddiagnosis_method_4] = '1' | Enter the type of PCR test: | dropdown, Required<br>1 Single PCR test for SARS-CoV-2<br>2 Multiplex PCR test which tested other respiratory pathogens in addition to SARS-CoV-2<br>3 Unknown/Unavailable |
| 26 | other_pathogen_4<br>Show the field ONLY if:<br>[covid] = '1' and [coviddiagnosis_method_4] = '1' | Did the patient have another respiratory pathogen identified within 7 days of the SARS CoV2 result? | dropdown, Required<br>1 Yes, same day<br>2 Yes, not on same day but within 7 days of test<br>3 No |
| 27 | respiratory_pathogens_4<br>Show the field ONLY if:<br>[other_pathogen_4] = '1' or [other_pathogen_4] = '2' | If yes, include name(s) of other respiratory pathogen (select all that apply from the following): | checkbox, Required<br>1 respiratory_pathogens_4___1 Adenovirus<br>2 respiratory_pathogens_4___2 Seasonal Cor (229E, HKU1, OC43)<br>3 respiratory_pathogens_4___3 Human Metapneumo<br>4 respiratory_pathogens_4___4 Human Rhinovirus/E<br>5 respiratory_pathogens_4___5 Parainfluenz 1,2,3 or 4<br>6 respiratory_pathogens_4___6 Respiratory s virus<br>7 respiratory_pathogens_4___7 Bacteria (Bor parapertussis Bordetella pe Chlamydia pneumoniae Mycoplasma pneumoniae<br>8 respiratory_pathogens_4___8 Unknown/Ur |
| 28 | coviddiagnosis_method_5<br>Show the field ONLY if:<br>[covid] = '1' and [coviddi | How was the fifth diagnosis made? | dropdown<br>1 Based on PCR result<br>2 Based on antigen result |

| | | | |
|---|---|---|---|
| | agnosis_multiple] = '1' and [covid_multiplereinfections] = '1' | | 3 Based on serology testing<br>4 Based on diagnosis code only<br>5 Self-reported by patient<br>6 Unsure/Not available |
| 29 | coviddiagnosis_date_5<br>Show the field ONLY if:<br>[covid] = '1' and [coviddiagnosis_multiple] = '1' and [covid_multiplereinfections] = '1' | Enter the date the fifth COVID-19 diagnosis was made: | text (date_mdy)<br>Field Annotation: @HIDEBUTTON |
| 30 | pcr_test_type_5<br>Show the field ONLY if:<br>[covid] = '1' and [coviddiagnosis_method_5] = '1' | Enter the type of PCR test: | dropdown, Required<br>1 Single PCR test for SARS-CoV-2<br>2 Multiplex PCR test which tested other respiratory pathogens in addition to SARS-CoV-2<br>3 Unknown/Unavailable |
| 31 | other_pathogen_5<br>Show the field ONLY if:<br>[covid] = '1' and [coviddiagnosis_method_5] = '1' | Did the patient have another respiratory pathogen identified within 7 days of the SARS CoV2 result? | dropdown, Required<br>1 Yes, same day<br>2 Yes, not on same day but within 7 days of test<br>3 No |
| 32 | respiratory_pathogens_5<br>Show the field ONLY if:<br>[other_pathogen_5] = '1' or [other_pathogen_5] = '2' | If yes, include name(s) of other respiratory pathogen (select all that apply from the following): | checkbox, Required<br>1 respiratory_pathogens_5___1 Adenovirus<br>2 respiratory_pathogens_5___2 Seasonal Cor (229E, HKU1, OC43)<br>3 respiratory_pathogens_5___3 Human Metapneum<br>4 respiratory_pathogens_5___4 Human Rhinovirus/E<br>5 respiratory_pathogens_5___5 Parainfluenz 1,2,3 or 4<br>6 respiratory_pathogens_5___6 Respiratory s virus<br>7 respiratory_pathogens_5___7 Bacteria (Bor parapertussi Bordetella pe Chlamydia pneumoniae Mycoplasma pneumoniae<br>8 respiratory_pathogens_5___8 Unknown/Ur |

| | | | |
|---|---|---|---|
| 33 | end_date<br><br>Show the field ONLY if:<br>[covid] = '1' | End Date: Enter the study Follow-up Period End Date for this patient. The End Date will be either a) 1 year after the last diagnosis of COVID-19, or b) if the patient has less than 1 year of follow-up from their last COVID diagnosis in the chart, the latest date available. | text (date_mdy), Required<br>Field Annotation: @HIDEBUTTON |
| 34 | end_date_reason<br><br>Show the field ONLY if:<br>[covid] = '1' | Enter the reason for the Follow-up Period End Date for this patient: | dropdown, Required<br>1 End of 1 year of follow-up from the patient's last COVID-19 diagnosis<br>2 End of available information in the patient's chart; less than 1 year of follow-up available for this patient<br><br>Field Annotation: @HIDEBUTTON |
| 35 | sex<br><br>Show the field ONLY if:<br>[covid] = '1' | Section Header: *Section 2. Demographics*<br>Patient Sex: | radio, Required<br>1 Male<br>2 Female |
| 36 | race<br><br>Show the field ONLY if:<br>[covid] = '1' | Race: | radio, Required<br>1 American Indian/Alaska Native<br>2 Asian/Native Hawaiian/Pacific Islander<br>3 Black/African-American<br>4 White<br>5 Multiracial<br>6 Other/Missing |
| 37 | ethnicity<br><br>Show the field ONLY if:<br>[covid] = '1' | Ethnicity: | radio, Required<br>1 Hispanic<br>2 Non-Hispanic<br>3 Unknown |
| 38 | payer<br><br>Show the field ONLY if:<br>[covid] = '1' | Payer: | radio, Required<br>1 Private/commercial<br>2 Public (Medicaid/SCHIP)<br>3 Other<br>4 Unknown/Unavailable |
| 39 | dob<br><br>Show the field ONLY if:<br>[covid] = '1' | Patient Date of Birth: | text (date_mdy), Required<br>Field Annotation: @HIDEBUTTON |
| 40 | covic_clinic<br><br>Show the field ONLY if:<br>[covid] = '1' | Section Header: *Section 3. COVID Treatments*<br>Was this patient seen in a long covid clinic/other clinic dedicated to the care of patients with PASC? | dropdown, Required<br>1 Yes<br>2 No<br>3 Unsure/Unavailable |

| | | | |
|---|---|---|---|
| 41 | evusheld<br><br>Show the field ONLY if:<br>[covid] = '1' | Did the patient receive evusheld (tixagevimab co-packaged with cilgavimab) as a prevention strategy against SARS-CoV-2? | dropdown, Required<br>1 Yes<br>2 No<br>3 Unsure/Unavailable |
| 42 | evusheld_doses<br><br>Show the field ONLY if:<br>[covid] = '1' and [evusheld] = '1' | If yes, how many doses of evusheld did the patient receive during the study period? | dropdown, Required<br>1 1<br>2 2<br>3 3<br>4 4<br>5 5<br>6 More than 5<br>9 Unsure/Unavailable |
| 43 | evusheld_dose1_date<br><br>Show the field ONLY if:<br>[covid] = '1' and [evusheld] = '1' and ([evusheld_doses] = '1' or [evusheld_doses] = '2' or [evusheld_doses] = '3' or [evusheld_doses] = '4' or [evusheld_doses] = '5' or [evusheld_doses] = '6') | What was the date of administration of the first dose of evusheld? | text (date_mdy), Required<br>Field Annotation: @HIDEBUTTON |
| 44 | evusheld_dose2_date<br><br>Show the field ONLY if:<br>[covid] = '1' and [evusheld] = '1' and ([evusheld_doses] = '2' or [evusheld_doses] = '3' or [evusheld_doses] = '4' or [evusheld_doses] = '5' or [evusheld_doses] = '6') | What was the date of administration of the second dose of evusheld? | text (date_mdy), Required<br>Field Annotation: @HIDEBUTTON |
| 45 | evusheld_dose3_date<br><br>Show the field ONLY if:<br>[covid] = '1' and [evusheld] = '1' and ([evusheld_doses] = '3' or [evusheld_doses] = '4' or [evusheld_doses] = '5' or [evusheld_doses] = '6') | What was the date of administration of the third dose of evusheld? | text (date_mdy), Required<br>Field Annotation: @HIDEBUTTON |
| 46 | evusheld_dose4_date<br><br>Show the field ONLY if: | What was the date of administration of the fourth dose of evusheld? | text (date_mdy), Required<br>Field Annotation: @HIDEBUTTON |

| | | | |
|---|---|---|---|
| | [covid] = '1' and [evusheld] = '1' and ([evusheld_doses] = '4' or [evusheld_doses] = '5' or [evusheld_doses] = '6') | | |
| 47 | evusheld_dose5_date<br><br>Show the field ONLY if:<br>[covid] = '1' and [evusheld] = '1' and ([evusheld_doses] = '5' or [evusheld_doses] = '6') | What was the date of administration of the fifth dose of evusheld? | text (date_mdy), Required<br>Field Annotation: @HIDEBUTTON |
| 48 | monoclonal<br><br>Show the field ONLY if:<br>[covid] = '1' | Did the patient receive monoclonal antibodies as their treatment for SARS CoV2? | dropdown, Required<br>1 Yes<br>2 No<br>3 Unsure/Unavailable |
| 49 | monoclonal_doses<br><br>Show the field ONLY if:<br>[covid] = '1' and [monoclonal] = '1' | If yes, how many doses of monoclonal antibodies did the patient receive during the study period? | dropdown, Required<br>1 1<br>2 2<br>3 3<br>4 4<br>5 5<br>6 More than 5<br>9 Unsure/Unavailable |
| 50 | monoclonal_date_1<br><br>Show the field ONLY if:<br>[covid] = '1' and [monoclonal] = '1' and ([monoclonal_doses] = '1' or [monoclonal_doses] = '2' or [monoclonal_doses] = '3' or [monoclonal_doses] = '4' or [monoclonal_doses] = '5' or [monoclonal_doses] = '6') | What was the date of administration of the first dose of monoclonal antibodies? | text (date_mdy), Required<br>Field Annotation: @HIDEBUTTON |
| 51 | monoclonal_type_1<br><br>Show the field ONLY if:<br>[covid] = '1' and [monoclonal] = '1' and ([monoclonal_doses] = '1' or [monoclonal_doses] = '2' or [monoclonal_doses] = '3' or [monoclonal_doses] = '4' or [monoclonal_doses] = '5' or [monoclonal_d | What type of monoclonal antibodies did the patient receive as their first dose of treatment? | dropdown, Required<br>1 Bebtelovimab<br>2 Bamlanivimab plus etesevimab<br>3 Casirivimab plus imdevimab<br>4 Sotrovimab<br>5 Unknown/Unavailable<br><br>Field Annotation: @HIDEBUTTON |

| | | | |
|---|---|---|---|
| | oses] = '6') | | |
| 52 | monoclonal_date_2<br><br>Show the field ONLY if:<br>[covid] = '1' and [monoclonal] = '1' and ([monoclonal_doses] = '2' or [monoclonal_doses] = '3' or [monoclonal_doses] = '4' or [monoclonal_doses] = '5' or [monoclonal_doses] = '6') | What was the date of administration of the second dose of monoclonal antibodies? | text (date_mdy), Required<br>Field Annotation: @HIDEBUTTON |
| 53 | monoclonal_type_2<br><br>Show the field ONLY if:<br>[covid] = '1' and [monoclonal] = '1' and ([monoclonal_doses] = '2' or [monoclonal_doses] = '3' or [monoclonal_doses] = '4' or [monoclonal_doses] = '5' or [monoclonal_doses] = '6') | What type of monoclonal antibodies did the patient receive as their second dose of treatment? | dropdown, Required<br>1 Bebtelovimab<br>2 Bamlanivimab plus etesevimab<br>3 Casirivimab plus imdevimab<br>4 Sotrovimab<br>5 Unknown/Unavailable<br><br>Field Annotation: @HIDEBUTTON |
| 54 | monoclonal_date_3<br><br>Show the field ONLY if:<br>[covid] = '1' and [monoclonal] = '1' and ([monoclonal_doses] = '3' or [monoclonal_doses] = '4' or [monoclonal_doses] = '5' or [monoclonal_doses] = '6') | What was the date of administration of the third dose of monoclonal antibodies? | text (date_mdy), Required<br>Field Annotation: @HIDEBUTTON |
| 55 | monoclonal_type_3<br><br>Show the field ONLY if:<br>[covid] = '1' and [monoclonal] = '1' and ([monoclonal_doses] = '3' or [monoclonal_doses] = '4' or [monoclonal_doses] = '5' or [monoclonal_doses] = '6') | What type of monoclonal antibodies did the patient receive as their third dose of treatment? | dropdown, Required<br>1 Bebtelovimab<br>2 Bamlanivimab plus etesevimab<br>3 Casirivimab plus imdevimab<br>4 Sotrovimab<br>5 Unknown/Unavailable<br><br>Field Annotation: @HIDEBUTTON |
| 56 | monoclonal_date_4<br><br>Show the field ONLY if:<br>[covid] = '1' and [monoclonal] = '1' and ([monoclonal_doses] = '4' or [monoclonal_doses] = '5' or [monoclonal_doses] = '6') | What was the date of administration of the fourth dose of monoclonal antibodies? | text (date_mdy), Required<br>Field Annotation: @HIDEBUTTON |
| 57 | monoclonal_type_4 | What type of monoclonal antibodies did the patient receive as their fourth dose of treatment? | dropdown, Required |

| | | | |
|---|---|---|---|
| | Show the field ONLY if: [covid] = '1' and [monoclonal] = '1' and ([monoclonal_doses] = '4' or [monoclonal_doses] = '5' or [monoclonal_doses] = '6') | | 1 Bebtelovimab<br>2 Bamlanivimab plus etesevimab<br>3 Casirivimab plus imdevimab<br>4 Sotrovimab<br>5 Unknown/Unavailable<br><br>Field Annotation: @HIDEBUTTON |
| 58 | monoclonal_date_5<br><br>Show the field ONLY if: [covid] = '1' and [monoclonal] = '1' and ([monoclonal_doses] = '5' or [monoclonal_doses] = '6') | What was the date of administration of the fifth dose of monoclonal antibodies? | text (date_mdy), Required<br>Field Annotation: @HIDEBUTTON |
| 59 | monoclonal_type_5<br><br>Show the field ONLY if: [covid] = '1' and [monoclonal] = '1' and ([monoclonal_doses] = '5' or [monoclonal_doses] = '6') | What type of monoclonal antibodies did the patient receive as their fifth dose of treatment? | dropdown, Required<br>1 Bebtelovimab<br>2 Bamlanivimab plus etesevimab<br>3 Casirivimab plus imdevimab<br>4 Sotrovimab<br>5 Unknown/Unavailable<br><br>Field Annotation: @HIDEBUTTON |
| 60 | vaccine_status<br><br>Show the field ONLY if: [covid] = '1' | Section Header: *Section 4. Vaccination*<br><br>Did this patient ever receive a COVID-19 vaccine? | dropdown, Required<br>1 Yes<br>2 No<br>3 Unknown |
| 61 | vaccine_date<br><br>Show the field ONLY if: [covid] = '1' and [vaccine_status] = '1' | Enter the date of their first COVID-19 vaccine: | text (date_mdy), Required<br>Field Annotation: @HIDEBUTTON |
| 62 | vaccine_brand<br><br>Show the field ONLY if: [covid] = '1' and [vaccine_status] = '1' | Select the vaccine they received as their first dose: | dropdown, Required<br>1 Pfizer/BioNTech<br>2 Moderna<br>3 J&J/Janssen<br>4 Unknown/Unavailable |
| 63 | vaccine_number<br><br>Show the field ONLY if: [covid] = '1' and [vaccine_status] = '1' | Did this patient receive multiple doses of the COVID-19 vaccine? | yesno, Required<br>1 Yes<br>0 No |
| 64 | vaccine_date_2<br><br>Show the field ONLY if: [covid] = '1' and [vaccine | Enter the date of their second COVID-19 vaccine: | text (date_mdy), Required<br>Field Annotation: @HIDEBUTTON |

| | | | |
|---|---|---|---|
| | _status] = '1' and [vaccine_number] = '1' | | |
| 65 | vaccine_brand_2<br>Show the field ONLY if:<br>[covid] = '1' and [vaccine_status] = '1' and [vaccine_number] = '1' | Select the vaccine they received as their second dose: | dropdown, Required<br>1 Pfizer/BioNTech<br>2 Moderna<br>3 J&J/Janssen<br>4 Unknown/Unavailable |
| 66 | vaccine_date_3<br>Show the field ONLY if:<br>[covid] = '1' and [vaccine_status] = '1' and [vaccine_number] = '1' | Enter the date of their third COVID-19 vaccine: | text (date_mdy)<br>Field Annotation: @HIDEBUTTON |
| 67 | vaccine_brand_3<br>Show the field ONLY if:<br>[covid] = '1' and [vaccine_status] = '1' and [vaccine_number] = '1' | Select the vaccine they received as their third dose: | dropdown<br>1 Pfizer/BioNTech<br>2 Moderna<br>3 J&J/Janssen<br>4 Unknown/Unavailable |
| 68 | vaccine_date_4<br>Show the field ONLY if:<br>[covid] = '1' and [vaccine_status] = '1' and [vaccine_number] = '1' | Enter the date of their fourth COVID-19 vaccine: | text (date_mdy)<br>Field Annotation: @HIDEBUTTON |
| 69 | vaccine_brand_4<br>Show the field ONLY if:<br>[covid] = '1' and [vaccine_status] = '1' and [vaccine_number] = '1' | Select the vaccine they received as their fourth dose: | dropdown<br>1 Pfizer/BioNTech<br>2 Moderna<br>3 J&J/Janssen<br>4 Unknown/Unavailable |
| 70 | vaccine_date_5<br>Show the field ONLY if:<br>[covid] = '1' and [vaccine_status] = '1' and [vaccine_number] = '1' | Enter the date of their fifth COVID-19 vaccine: | text (date_mdy)<br>Field Annotation: @HIDEBUTTON |
| 71 | vaccine_brand_5<br>Show the field ONLY if:<br>[covid] = '1' and [vaccine_status] = '1' and [vaccine_number] = '1' | Select the vaccine they received as their fifth dose: | dropdown<br>1 Pfizer/BioNTech<br>2 Moderna<br>3 J&J/Janssen<br>4 Unknown/Unavailable |
| 72 | functional_available<br>Show the field ONLY if: | Section Header: *Section 5. Functional Outcomes This section is looking for results from the period starting 28 days after the first date of COVID-19 infection.*<br>Is there any documentation available in the patient's | dropdown, Required<br>1 Yes<br>0 No |

| | | | |
|---|---|---|---|
| | [covid] = '1' | chart on functional outcomes (Behavioral/Psychological symptoms, school missingness, extracurricular activity changes, etc) | 2 Unsure/Unavailable |
| 73 | behavioral_symptoms<br>Show the field ONLY if:<br>[covid] = '1' and [functional_available] = '1' | Has the patient had behavioral and/or psychological symptoms post-COVID-19 infection? | dropdown, Required<br>1 Yes<br>0 No<br>2 Unsure/Unavailable |
| 74 | behavioral_symptoms_describe<br>Show the field ONLY if:<br>[covid] = '1' and [behavioral_symptoms] = '1' | Please describe: | notes, Required |
| 75 | behavior_symptoms_prior<br>Show the field ONLY if:<br>[covid] = '1' and [behavioral_symptoms] = '1' | Were any of these symptoms present prior to COVID-19 infection? | radio, Required<br>1 Yes<br>2 No<br>3 Unknown/Unavailable |
| 76 | behavior_symptoms_prior_detail<br>Show the field ONLY if:<br>[covid] = '1' and [behavioral_symptoms] = '1' and [behavior_symptoms_prior] = '1' | Describe symptoms present prior to COVID: | notes, Required |
| 77 | behavior_symptoms_change<br>Show the field ONLY if:<br>[covid] = '1' and [behavioral_symptoms] = '1' and [behavior_symptoms_prior] = '1' | Did the symptoms change post COVID19? | radio, Required<br>1 Yes- symptoms became worse post-COVID<br>2 Yes- symptoms improved post COVID<br>3 No, no change |
| 78 | behavior_change_description<br>Show the field ONLY if:<br>[covid] = '1' and [behavioral_symptoms] = '1' and [behavior_symptoms_prior] = '1' and [behavior_symptoms_change] = '1' or [behavior_symptoms_change] = '2' | Please describe how symptoms changed post COVID. | notes, Required |
| 79 | psych_referral<br>Show the field ONLY if:<br>[covid] = '1' and [functional_available] = '1' | Has the patient been referred to psychological or behavior services since COVID-19 infection? | yesno, Required<br>1 Yes<br>0 No |
| 80 | psych_referral_desc | Please describe the psychological or behavior | notes, Required |

| | | | |
|---|---|---|---|
| | ribe<br>Show the field ONLY if:<br>[covid] = '1' and [functional_available] = '1' and [psych_referral] = '1' | services the patient was referred to since COVID-19 infection: | |
| 81 | extracurriculars<br>Show the field ONLY if:<br>[covid] = '1' and [functional_available] = '1' | Is there any evidence the patient has missed extracurricular activities (sports, band, dance, etc.) since COVID-19 infection? | yesno, Required<br>1 Yes<br>0 No |
| 82 | extracurriculars_describe<br>Show the field ONLY if:<br>[covid] = '1' and [functional_available] = '1' and [extracurriculars] = '1' | Please describe the evidence the patient has missed extracurricular activities (sports, band, dance, etc.) since COVID-19 infection. | notes, Required |
| 83 | therapy_postcovid<br>Show the field ONLY if:<br>[covid] = '1' and [coviddiagnosis_age] = '1' and [functional_available] = '1' | Has the patient required any of the following since COVID-19 infection? | checkbox, Required<br>1 therapy_postcovid___1 Physical therapy (PT)<br>2 therapy_postcovid___2 Occupational therapy (OT)<br>3 therapy_postcovid___3 Speech language therapy (SLP)<br>4 therapy_postcovid___4 Special instruction<br>5 therapy_postcovid___5 Early intervention (EI) referral/Individual Family Service Plan (IFSP)<br>6 therapy_postcovid___6 None of the above |
| 84 | pt_date<br>Show the field ONLY if:<br>[covid] = '1' and [coviddiagnosis_age] = '1' and [functional_available] = '1' and [therapy_postcovid(1)] = '1' | Enter earliest date Physical Therapy treatment began post COVID-19 infection. | text (date_mdy), Required<br>Field Annotation: @HIDEBUTTON |
| 85 | pt_status<br>Show the field ONLY if:<br>[covid] = '1' and [coviddiagnosis_age] = '1' and [functional_available] = '1' and [therapy_postcovid(1)] = '1' | Enter current status of Physical Therapy treatment: | radio, Required<br>1 Ongoing<br>2 Completed<br>3 Unsure<br>Field Annotation: @HIDEBUTTON |

| | | | |
|---|---|---|---|
| 86 | pt_last_date<br><br>Show the field ONLY if:<br>[covid] = '1' and [covidiagnosis_age] = '1' and [functional_available] = '1' and [therapy_postcovid(1)] = '1' and [pt_status] = '2' | Enter date of last Physical Therapy treatment: | text (date_mdy), Required<br>Field Annotation: @HIDEBUTTON |
| 87 | ot_date<br><br>Show the field ONLY if:<br>[covid] = '1' and [covidiagnosis_age] = '1' and [functional_available] = '1' and [therapy_postcovid(2)] = '1' | Enter earliest date Occupational Therapy treatment began post COVID-19 infection. | text (date_mdy), Required<br>Field Annotation: @HIDEBUTTON |
| 88 | ot_status<br><br>Show the field ONLY if:<br>[covid] = '1' and [covidiagnosis_age] = '1' and [functional_available] = '1' and [therapy_postcovid(2)] = '1' | Enter current status of Occupational Therapy treatment: | radio, Required<br>1 Ongoing<br>2 Completed<br>3 Unsure<br><br>Field Annotation: @HIDEBUTTON |
| 89 | ot_last_date<br><br>Show the field ONLY if:<br>[covid] = '1' and [covidiagnosis_age] = '1' and [functional_available] = '1' and [therapy_postcovid(2)] = '1' and [ot_status] = '2' | Enter date of last Occupational Therapy treatment: | text (date_mdy), Required<br>Field Annotation: @HIDEBUTTON |
| 90 | speech_therapy_date<br><br>Show the field ONLY if:<br>[covid] = '1' and [covidiagnosis_age] = '1' and [functional_available] = '1' and [therapy_postcovid(3)] = '1' | Enter earliest date Speech Language Therapy treatment began post COVID-19 infection. | text (date_mdy), Required<br>Field Annotation: @HIDEBUTTON |
| 91 | speech_therapy_status<br><br>Show the field ONLY if:<br>[covid] = '1' and [covidiagnosis_age] = '1' and [functional_available] = '1' and [therapy_postcovid(3)] = '1' | Enter current status of Speech Language Therapy treatment: | radio, Required<br>1 Ongoing<br>2 Completed<br>3 Unsure<br><br>Field Annotation: @HIDEBUTTON |
| 92 | speech_therapy_lastdate | Enter date of last Speech Language Therapy treatment: | text (date_mdy), Required<br>Field Annotation: @HIDEBUTTON |

| | | | |
|---|---|---|---|
| | Show the field ONLY if: [covid] = '1' and [covidiagnosis_age] = '1' and [functional_available] = '1' and [therapy_postcovid(3)] = '1' and [speech_therapy_status] = '2' | | |
| 93 | special_instruction_date<br><br>Show the field ONLY if: [covid] = '1' and [covidiagnosis_age] = '1' and [functional_available] = '1' and [therapy_postcovid(4)] = '1' | Enter earliest date Special Instruction treatment began post COVID-19 infection. | text (date_mdy), Required<br>Field Annotation: @HIDEBUTTON |
| 94 | special_instruction_status<br><br>Show the field ONLY if: [covid] = '1' and [covidiagnosis_age] = '1' and [functional_available] = '1' and [therapy_postcovid(4)] = '1' | Enter current status of Special Instruction treatment: | radio, Required<br>1 Ongoing<br>2 Completed<br>3 Unsure<br><br>Field Annotation: @HIDEBUTTON |
| 95 | special_instruction_lastdate<br><br>Show the field ONLY if: [covid] = '1' and [covidiagnosis_age] = '1' and [functional_available] = '1' and [therapy_postcovid(4)] = '1' and [special_instruction_status] = '2' | Enter date of last Special Instruction treatment: | text (date_mdy), Required<br>Field Annotation: @HIDEBUTTON |
| 96 | early_intervene_date<br><br>Show the field ONLY if: [covid] = '1' and [covidiagnosis_age] = '1' and [functional_available] = '1' and [therapy_postcovid(5)] = '1' | Enter earliest date Early intervention (EI) referral/Individual Family Service Plan (IFSP) began post COVID-19 infection. | text (date_mdy), Required<br>Field Annotation: @HIDEBUTTON |
| 97 | early_intervene_status<br><br>Show the field ONLY if: [covid] = '1' and [covidiagnosis_age] = '1' and [functional_available] = '1' and [therapy_postcovid(5)] = '1' | Enter current status of Early intervention (EI) referral/Individual Family Service Plan (IFSP) treatment. | radio, Required<br>1 Ongoing<br>2 Completed<br>3 Unsure<br><br>Field Annotation: @HIDEBUTTON |
| 98 | early_intervene_las | Enter date of last Early intervention (EI) | text (date_mdy), Required |

| | | | |
|---|---|---|---|
| | tdate<br><br>Show the field ONLY if:<br>[covid] = '1' and [covidd iagnosis_age] = '1' and [f unctional_available] = '1' and [therapy_postcovid( 5)] = '1' and [early_interv ene_status] = '2' | referral/Individual Family Service Plan (IFSP) treatment. | Field Annotation: @HIDEBUTTON |
| 99 | school_missing_evid ence<br><br>Show the field ONLY if:<br>[covid] = '1' and [functio nal_available] = '1' and [c oviddiagnosis_age] = '2' | Is there any evidence that the patient has missed school following recovery from COVID-19 infection? | yesno, Required<br>1 Yes<br>0 No |
| 100 | school_missing_deta ils<br><br>Show the field ONLY if:<br>[covid] = '1' and [functio nal_available] = '1' and [c oviddiagnosis_age] = '2' and [school_missing_evi dence] = '1' | Enter any additional details available about school missingness (frequency, timing after infection, etc): | notes, Required |
| 101 | modified_school<br><br>Show the field ONLY if:<br>[covid] = '1' and [functio nal_available] = '1' and [c oviddiagnosis_age] = '2' | Has the patient transitioned to a modified school program? | radio, Required<br>1 Yes- home schooling<br>2 Yes- Virtual school<br>3 Yes- hybrid schedule<br>4 No<br>5 Unsure/Unavailable |
| 102 | modified_school_rea son<br><br>Show the field ONLY if:<br>[covid] = '1' and [functio nal_available] = '1' and [c oviddiagnosis_age] = '2' and [modified_school] = '1' or [modified_school] = '2' or [modified_schoo l] = '3' | Is there an indication as to why the patient transitioned to a modified school system? | radio, Required<br>1 Yes- parental preference<br>2 Yes- school mandated<br>3 Other<br>4 Unsure/No Information available |
| 103 | modified_school_ret urn<br><br>Show the field ONLY if:<br>[covid] = '1' and [functio nal_available] = '1' and [c oviddiagnosis_age] = '2' and [modified_school] = '1' or [modified_school] = '2' or [modified_schoo | Has the patient returned to full in person learning? | radio, Required<br>1 Yes<br>2 No<br>3 Unsure/Unavailable |

| | | | |
|---|---|---|---|
| | l] = '3' | | |
| 104 | school_return_timing<br><br>Show the field ONLY if:<br>[covid] = '1' and [functional_available] = '1' and [coviddiagnosis_age] = '2' and [modified_school_return] = '1' | How long after COVID-19 infection did the patient return to full in person learning? | radio, Required<br>1 Within 2 week<br>2 Within 1 month<br>3 Within 3 months<br>4 More than 3 months<br>5 Unsure/Not Available |
| 105 | school_testing<br><br>Show the field ONLY if:<br>[covid] = '1' and [functional_available] = '1' and [coviddiagnosis_age] = '2' | Starting from the period 28 days after initial COVID-19 infection, has the patient: | checkbox, Required<br>1 school_testing___1 Been referred for neuropsychological testing (either through school of otherwise)<br>2 school_testing___2 Requested an individualized education plan (IEP)<br>3 school_testing___3 Requested a 504 plan<br>4 school_testing___4 Requested private tutoring<br>5 school_testing___5 Requested any other assistance with school learning<br>6 school_testing___6 None of the above<br>7 school_testing___7 Unsure/Unavailable |
| 106 | school_testing_other<br><br>Show the field ONLY if:<br>[covid] = '1' and [functional_available] = '1' and [coviddiagnosis_age] = '2' and [school_testing(5)] = '1' | Please describe the other assistance requested: | notes, Required |
| 107 | neurotest_new<br><br>Show the field ONLY if:<br>[covid] = '1' and [functional_available] = '1' and [coviddiagnosis_age] = '2' and [school_testing(1)] = '1' | Was neuropsychological testing (either through school of otherwise) newly requested 28 days or more post COVID-19 infection? | dropdown, Required<br>1 Yes- patient never had this support prior to COVID-19 infection<br>2 No, but the level of support patient needed increased post COVID-19 infection<br>3 No, these were in place at the same level of support pre-COVID-19<br>4 Unsure/Not available |
| 108 | iep_new | Was an Individualized Education Plan (IEP) newly | dropdown, Required |

| | | | |
|---|---|---|---|
| | Show the field ONLY if: [covid] = '1' and [functional_available] = '1' and [coviddiagnosis_age] = '2' and [school_testing(2)] = '1' | requested 28 days or more post COVID-19 infection? | 1 Yes- patient never had this support prior to COVID-19 infection<br>2 No, but the level of support patient needed increased post COVID-19 infection<br>3 No, these were in place at the same level of support pre-COVID-19<br>4 Unsure/Not available |
| 109 | plan_new<br><br>Show the field ONLY if: [covid] = '1' and [functional_available] = '1' and [coviddiagnosis_age] = '2' and [school_testing(3)] = '1' | Was a 504 plan newly requested 28 days or more post COVID-19 infection? | dropdown, Required<br>1 Yes- patient never had this support prior to COVID-19 infection<br>2 No, but the level of support patient needed increased post COVID-19 infection<br>3 No, these were in place at the same level of support pre-COVID-19<br>4 Unsure/Not available |
| 110 | tutoring_new<br><br>Show the field ONLY if: [covid] = '1' and [functional_available] = '1' and [coviddiagnosis_age] = '2' and [school_testing(4)] = '1' | Was a private tutoring newly requested 28 days or more post COVID-19 infection? | dropdown, Required<br>1 Yes- patient never had this support prior to COVID-19 infection<br>2 No, but the level of support patient needed increased post COVID-19 infection<br>3 No, these were in place at the same level of support pre-COVID-19<br>4 Unsure/Not available |
| 111 | school_reported_difficult<br><br>Show the field ONLY if: [covid] = '1' and [functional_available] = '1' and [coviddiagnosis_age] = '2' | Has the patient or parent reported difficulties in school following COVID-19 infection? | yesno, Required<br>1 Yes<br>0 No |
| 112 | school_reported_describe<br><br>Show the field ONLY if: [covid] = '1' and [functional_available] = '1' and [coviddiagnosis_age] = '2' and [school_reported_difficult] = '1' | Please describe the difficulties reported: | notes, Required |
| 113 | therapy_schoolkids<br><br>Show the field ONLY if: [covid] = '1' and [functional_available] = '1' and [coviddiagnosis_age] = '2' | Has the patient required any of the following in the period starting 28 days after initial COVID infection: | checkbox, Required<br>1 therapy_schoolkids___1 Physical therapy (PT)<br>2 therapy_schoolkids___2 Occupational therapy (OT) |

| | | | |
|---|---|---|---|
| | | | 3 therapy_schoolkids___3 Speech language therapy (SLP)<br>4 therapy_schoolkids___4 Other<br>5 therapy_schoolkids___5 None |
| 114 | other_therapy_schoolkids<br><br>Show the field ONLY if:<br>[therapy_schoolkids(4)] = '1' | Please describe Other: | notes, Required |
| 115 | physical_therapy_new<br><br>Show the field ONLY if:<br>[covid] = '1' and [functional_available] = '1' and [coviddiagnosis_age] = '2' and [therapy_schoolkids(1)] = '1' | Was Physical Therapy newly requested 28 days or more post COVID-19 infection? | dropdown, Required<br>1 Yes- patient never had this support prior to COVID-19 infection<br>2 No, but the level of support patient needed increased post COVID-19 infection<br>3 No, these were in place at the same level of support pre-COVID-19<br>4 Unsure/Not available |
| 116 | physical_therapy_new_date<br><br>Show the field ONLY if:<br>[covid] = '1' and [functional_available] = '1' and [coviddiagnosis_age] = '2' and [therapy_schoolkids(1)] = '1' and [physical_therapy_new] = '1' or [physical_therapy_new] = '2' | Enter date Physical Therapy began or increased post COVID-19 infection. | text (date_mdy), Required<br>Field Annotation: @HIDEBUTTON |
| 117 | physical_therapy_new_status<br><br>Show the field ONLY if:<br>[covid] = '1' and [functional_available] = '1' and [coviddiagnosis_age] = '2' and [therapy_schoolkids(1)] = '1' and [physical_therapy_new] = '1' or [physical_therapy_new] = '2' | Enter current status of Physical Therapy treatment. | radio, Required<br>1 Ongoing<br>2 Completed<br>3 Unsure/Unavailable<br><br>Field Annotation: @HIDEBUTTON |
| 118 | physical_therapy_lastdate<br><br>Show the field ONLY if:<br>[covid] = '1' and [functional_available] = '1' and [coviddiagnosis_age] = '2' and [therapy_schoolkids( | Enter date of last Physical Therapy treatment. | text (date_mdy), Required<br>Field Annotation: @HIDEBUTTON |

| | | | |
|---|---|---|---|
| | 1)] = '1' and [physical_therapy_new] = '1' or [physical_therapy_new] = '2' and [physical_therapy_new_status] = '2' | | |
| 119 | occupational_therapy_new<br>Show the field ONLY if:<br>[covid] = '1' and [functional_available] = '1' and [coviddiagnosis_age] = '2' and [therapy_schoolkids(2)] = '1' | Was Occupational Therapy newly requested 28 days or more post COVID-19 infection? | dropdown, Required<br>1 Yes- patient never had this support prior to COVID-19 infection<br>2 No, but the level of support patient needed increased post COVID-19 infection<br>3 No, these were in place at the same level of support pre-COVID-19<br>4 Unsure/Not available |
| 120 | occupational_therapy_new_date<br>Show the field ONLY if:<br>[covid] = '1' and [functional_available] = '1' and [coviddiagnosis_age] = '2' and [therapy_schoolkids(2)] = '1' and [occupational_therapy_new] = '1' or [occupational_therapy_new] = '2' | Enter date Occupational Therapy began or increased post COVID-19 infection. | text (date_mdy), Required<br>Field Annotation: @HIDEBUTTON |
| 121 | occupational_therapy_new_status<br>Show the field ONLY if:<br>[covid] = '1' and [functional_available] = '1' and [coviddiagnosis_age] = '2' and [therapy_schoolkids(2)] = '1' and [occupational_therapy_new] = '1' or [occupational_therapy_new] = '2' | Enter current status of Occupational Therapy treatment. | radio, Required<br>1 Ongoing<br>2 Completed<br>3 Unsure/Unavailable<br>Field Annotation: @HIDEBUTTON |
| 122 | occupational_therapy_last_date<br>Show the field ONLY if:<br>[covid] = '1' and [functional_available] = '1' and [coviddiagnosis_age] = '2' and [therapy_schoolkids(2)] = '1' and [occupational_therapy_new_status] = '2' | Enter date of last Occupational Therapy treatment. | text (date_mdy), Required<br>Field Annotation: @HIDEBUTTON |
| 123 | speech_therapy_new<br>Show the field ONLY if: | Was Speech Language Therapy newly requested 28 days or more post COVID-19 infection? | dropdown, Required<br>1 Yes- patient never had this support |

| | | | |
|---|---|---|---|
| | [covid] = '1' and [functional_available] = '1' and [coviddiagnosis_age] = '2' and [therapy_schoolkids(3)] = '1' | | prior to COVID-19 infection<br>2 No, but the level of support patient needed increased post COVID-19 infection<br>3 No, these were in place at the same level of support pre-COVID-19<br>4 Unsure/Not available |
| 124 | speech_therapy_new_date<br><br>Show the field ONLY if:<br>[covid] = '1' and [functional_available] = '1' and [coviddiagnosis_age] = '2' and [therapy_schoolkids(3)] = '1' and [speech_therapy_new] = '1' or [speech_therapy_new] = '2' | Enter date Speech Language Therapy began or increased post COVID-19 infection. | text (date_mdy), Required<br>Field Annotation: @HIDEBUTTON |
| 125 | speech_therapy_new_status<br><br>Show the field ONLY if:<br>[covid] = '1' and [functional_available] = '1' and [coviddiagnosis_age] = '2' and [therapy_schoolkids(3)] = '1' and [speech_therapy_new] = '1' or [speech_therapy_new] = '2' | Enter current status of Speech Language Therapy treatment. | radio, Required<br>1 Ongoing<br>2 Completed<br>3 Unsure/Unavailable<br><br>Field Annotation: @HIDEBUTTON |
| 126 | speech_therapy_lastdate_schoolkids<br><br>Show the field ONLY if:<br>[covid] = '1' and [functional_available] = '1' and [coviddiagnosis_age] = '2' and [therapy_schoolkids(3)] = '1' and [speech_therapy_new_status] = '2' | Enter date of last Speech Language Therapy treatment. | text (date_mdy), Required<br>Field Annotation: @HIDEBUTTON |
| 127 | mental_health_history<br><br>Show the field ONLY if:<br>[covid] = '1' | Section Header: *Section 6. Mental Health History*<br><br>Does the patient have a history BEFORE their index date of diagnoses or care for mental health, psychological, or behavioral health conditions? If Yes, please indicate which category of condition (select all that apply) | checkbox, Required<br>1 mental_health_history___1 Attention deficit/hyperact[illegible] disorder<br>2 mental_health_history___2 Autism spectru[illegible]<br>3 mental_health_history___3 Intellectual/dev[illegible] disability<br>4 mental_health_history___4 Anxiety (include[illegible] repetitive disor[illegible]<br>5 mental_health_history___5 Depression<br>6 mental_health_history___6 Other mood dis[illegible] |

| | | | |
|---|---|---|---|
| | | | 6 — bipolar, dysthy[illegible]<br>7 mental_health_history___7 Other<br>9 mental_health_history___9 No, this patient mental health c[illegible] before their Ind[illegible]<br>8 mental_health_history___8 Unavailable |
| 128 | mental_health_other<br><br>Show the field ONLY if:<br>[mental_health_history(7)] = '1' | If Other, please provide detail: | notes, Required |
| 129 | mental_health_current<br><br>Show the field ONLY if:<br>[covid] = '1' | Does the patient have a history AFTER their index date of diagnoses or care for mental health, psychological, or behavioral health conditions? If Yes, please indicate which category of condition (select all that apply) | checkbox, Required<br>1 mental_health_current___1 Attention deficit/hyperac[illegible] disorder<br>2 mental_health_current___2 Autism spectru[illegible]<br>3 mental_health_current___3 Intellectual/de[illegible] disability<br>4 mental_health_current___4 Anxiety (includ[illegible] repetitive diso[illegible]<br>5 mental_health_current___5 Depression<br>6 mental_health_current___6 Other mood di[illegible] bipolar, dysthy[illegible]<br>7 mental_health_current___7 Other<br>9 mental_health_current___9 No, this patient mental health [illegible] after their Inde[illegible]<br>8 mental_health_current___8 Unavailable |
| 130 | mental_health_after_2<br><br>Show the field ONLY if:<br>[mental_health_current(7)] = '1' | If Other, please provide detail: | notes, Required |
| 131 | mental_health_history_other<br><br>Show the field ONLY if:<br>[covid] = '1' | Does the patient have any other evidence in their medical record of (diagnosed or undiagnosed) mental health, psychological, or behavioral health conditions or problems? This can be assessed by searching the chart for keywords following the guidance in the review manual:For Epic chart search (anxi OR depr OR coping OR confu OR mood OR brain OR concentra OR fatig OR worry OR relationships OR friend) | radio, Required<br>1 Yes<br>2 No<br>3 Unavailable |
| 132 | mental_health_other_descrip | If yes, please describe briefly. You may copy and paste phrases from clinical notes in the record, but be certain that you do not include any identifying | notes, Required |

| | | | |
|---|---|---|---|
| | Show the field ONLY if: [mental_health_history_other]= '1' | information or any phrases from psychotherapy records. | |
| 133 | misc_diagnosis<br><br>Show the field ONLY if: [covid] = '1' | Section Header: *Section 7. Algorithm Validation*<br>Are there any references to the patient being diagnosed with MIS-C at any time during study period? | yesno, Required<br>1 Yes<br>0 No |
| 134 | misc_terms<br><br>Show the field ONLY if: [covid] = '1' and [misc_diagnosis] = '1' | Enter the MIS-C diagnosis references in the patient's chart: | notes, Required |
| 135 | misc_date<br><br>Show the field ONLY if: [covid] = '1' and [misc_diagnosis] = '1' | Enter the first MIS-C diagnosis date: | text (date_mdy), Required<br>Field Annotation: @HIDEBUTTON |
| 136 | misc_date_2<br><br>Show the field ONLY if: [covid] = '1' and [misc_diagnosis] = '1' | Enter the second MIS-C diagnosis date, if applicable. If no second diagnosis, skip this field. | text (date_mdy)<br>Field Annotation: @HIDEBUTTON |
| 137 | u099_diagnosis<br><br>Show the field ONLY if: [covid] = '1' | Did the patient have any references to a diagnosis for "PASC" at any time during study period? | yesno, Required<br>1 Yes<br>0 No |
| 138 | pasc_terms<br><br>Show the field ONLY if: [covid] = '1' and [u099_diagnosis] = '1' | Enter the PASC diagnoses references in the patient's chart: | notes, Required |
| 139 | pasc_date<br><br>Show the field ONLY if: [covid] = '1' and [u099_diagnosis] = '1' | Enter the first PASC diagnosis date: | text (date_mdy), Required<br>Field Annotation: @HIDEBUTTON |
| 140 | pasc_date_2<br><br>Show the field ONLY if: [covid] = '1' and [u099_diagnosis] = '1' | Enter the second PASC diagnosis date, if applicable. If no second diagnosis for this patient, leave field blank. | text (date_mdy)<br>Field Annotation: @HIDEBUTTON |
| 141 | pcr_antigen_positive<br><br>Show the field ONLY if: [covid] = '1' | Did the patient have a positive SARS-CoV-2 RT PCR/ antigen test during the study period? | yesno, Required<br>1 Yes<br>0 No |
| 142 | sars_test_date<br><br>Show the field ONLY if: [covid] = '1' and [pcr_antigen_positive] = '1' | Enter the positive SARS-CoV-2 test date: | text (date_mdy), Required<br>Field Annotation: @HIDEBUTTON |
| 143 | reason_pcr_antigen_ | If no, please select the reason why: | dropdown, Required |

| | | | |
|---|---|---|---|
| | no<br>Show the field ONLY if:<br>[covid] = '1' and [pcr_antigen_positive] = '0' | | 1 -The patient never had a SARS-CoV-2 RT PCR/ antigen test<br>2 -The patient was tested but the SARS-CoV-2 RT PCR/ antigen test result was negative |
| 144 | serology_test<br>Show the field ONLY if:<br>[covid] = '1' | Did the patient have SARS-CoV-2 serology positive test result? | dropdown, Required<br>1 Yes<br>2 No, this patient never had a SARS-CoV-2 serology test<br>3 No, the patient had a SARS-CoV-2 serology test, but the results were negative<br>4 No, the patient had a SARS-CoV-2 serology test, but the results were positive for antibodies against vaccine antigens (anti-S/RBD rather than anti-N) |
| 145 | serology_test_reason<br>Show the field ONLY if:<br>[covid] = '1' and ([serology_test] = '1' or [serology_test] = '3' or [serology_test] = '4') | Select the reason why the patient was given a serology test (select all that apply): | checkbox, Required<br>1 serology_test_reason___1 Symptomatic<br>2 serology_test_reason___2 Asymptomatic<br>3 serology_test_reason___3 Concern for PAS[illegible]<br>7 serology_test_reason___7 Concern for MIS-[illegible]<br>4 serology_test_reason___4 Exposed to COVI[illegible] 19<br>5 serology_test_reason___5 Other<br>6 serology_test_reason___6 Unsure/Unavaila[illegible] |
| 146 | serology_test_date<br>Show the field ONLY if:<br>[covid] = '1' and ([serology_test] = '1' or [serology_test] = '3' or [serology_test] = '4') | Enter the date of the initial serology test: | text (date_mdy), Required<br>Field Annotation: @HIDEBUTTON |
| 147 | kawasaki_diagnosis<br>Show the field ONLY if:<br>[covid] = '1' | Did the patient have a diagnosis for Kawasaki disease assigned within 42 days of a positive RT-PCR/Antigen test? | dropdown, Required<br>1 Yes<br>2 No, this patient never had a diagnosis term for Kawasaki disease<br>3 No, this patient had a diagnosis term for Kawasaki disease but it was outside of the 42 day window of RT-PCR/Antigen test positivity |
| 148 | antigen_test_date<br>Show the field ONLY if:<br>[covid] = '1' and [kawasaki_diagnosis] = '3' | Enter the date of the first RT-PCR/Antigen positive test: | text (date_mdy), Required<br>Field Annotation: @HIDEBUTTON |

| | | | |
|---|---|---|---|
| 149 | kawasaki_terms<br><br>Show the field ONLY if:<br>[covid] = '1' and [kawasaki_diagnosis] = '1' | Enter the diagnosis terms for Kawasaki Disease found in the patient chart. | text, Required |
| 150 | kawasaki_diag_date<br><br>Show the field ONLY if:<br>[covid] = '1' and [kawasaki_diagnosis] = '3' | Enter the date of the first Kawasaki disease diagnosis: | text (date_mdy), Required<br>Field Annotation: @HIDEBUTTON |
| 151 | conditions_post<br><br>Show the field ONLY if:<br>[covid] = '1' | Did the patient have any occurrence of the following conditions, on Day 28 or later after initial COVID-19 positive diagnosis?<br><br>Select all that apply. | checkbox, Required |

| | | |
|---|---|---|
| 1 | conditions_post___1 | COVID-19 |
| 2 | conditions_post___2 | Acute respiratory distress syndrome |
| 3 | conditions_post___3 | Loss of smell |
| 4 | conditions_post___4 | Loss of taste |
| 5 | conditions_post___5 | Other changes in smell/taste |
| 6 | conditions_post___6 | Myocarditis |
| 7 | conditions_post___7 | Pericarditis |
| 8 | conditions_post___8 | Myositis |
| 9 | conditions_post___9 | Other/ill- defined heart disease |
| 10 | conditions_post___10 | Thrombophlebitis and thromboembolism |
| 11 | conditions_post___11 | Aplastic anemia |
| 12 | conditions_post___12 | Brain fog |
| 13 | conditions_post___13 | Abnormal liver enzymes |
| 14 | conditions_post___14 | Dysautonomia |
| 15 | conditions_post___15 | Fatigue |
| 19 | conditions_post___19 | Abdominal Pain |
| 20 | conditions_post___20 | Acute Kidney Injury |
| 18 | conditions_post___18 | Other (including any rheumatic, musculoskeletal, or non-stomatitis herpetic/chronic viral conditions) |
| 16 | conditions_post___16 | No- the patient never had any of these diagnoses |
| 17 | conditions_post___17 | No- the patient |

| | | | |
|---|---|---|---|
| | | | had some of these diagnoses but prior to Day 28 after initial COVID positive diagnosis |
| 152 | pasc_other<br><br>Show the field ONLY if:<br>[conditions_post(18)] = '1' | List the Other conditions and first 2 diagnosis dates of each condition: | notes, Required |
| 153 | pasc_other_smell<br><br>Show the field ONLY if:<br>[conditions_post(5)] = '1' | List the Other Changes in Smell/Taste conditions and first 2 diagnosis dates of each condition: | notes, Required |
| 154 | pasc_other_heart<br><br>Show the field ONLY if:<br>[conditions_post(9)] = '1' | List the Other/Ill-defined Heart Disease conditions and first 2 diagnosis dates of each condition: | notes, Required |
| 155 | covid_diagnosis_pasc_other<br><br>Show the field ONLY if:<br>[conditions_post(18)] = '1' | Did the physician state in the chart whether any of these Other conditions were due to PASC? | dropdown, Required<br>1 Yes, the physician stated one or more of the conditions was due to PASC<br>2 No, the physician stated that all the conditions were due to something other than PASC<br>3 Unknown/Not Available<br><br>Field Annotation: @HIDEBUTTON |
| 156 | covid_diagnosis_pasc_y_other<br><br>Show the field ONLY if:<br>[conditions_post(18)] = '1' and [covid_diagnosis_pasc_other] = '1' | Copy and paste the language from the physician notes that says the continued Other COVID diagnosis was related to PASC (make sure to remove any PHI, such as MRNs or names): | notes, Required<br>Field Annotation: @HIDEBUTTON |
| 157 | covid_date_followup<br><br>Show the field ONLY if:<br>[conditions_post(1)] = '1' | Enter the first COVID diagnosis date found during the follow-up period: | text (date_mdy), Required<br>Field Annotation: @HIDEBUTTON |
| 158 | covid_endstatus<br><br>Show the field ONLY if:<br>[conditions_post(1)] = '1' | At the end of the follow-up period, the COVID diagnosis was: | dropdown, Required<br>1 Resolved<br>2 Ongoing<br>3 Unknown/Unavailable<br><br>Field Annotation: @HIDEBUTTON |
| 159 | covid_diagnosis_pasc<br><br>Show the field ONLY if:<br>[conditions_post(1)] = '1' | Did the physician state in the chart whether this continued COVID diagnosis was due to PASC? | dropdown, Required<br>1 Yes, the physician stated this condition was due to PASC |

| | | | |
|---|---|---|---|
| | | | 2 No, the physician stated that this condition was due to something other than PASC<br>3 Unknown/Not Available<br><br>Field Annotation: @HIDEBUTTON |
| 160 | covid_diagnosis_pasc_y<br><br>Show the field ONLY if:<br>[conditions_post(1)] = '1' and [covid_diagnosis_pasc] = '1' | Copy and paste the language from the physician notes that says this continued COVID diagnosis was related to PASC (make sure to remove any PHI, such as MRNs or names): | notes, Required<br>Field Annotation: @HIDEBUTTON |
| 161 | acuteres_date_followup<br><br>Show the field ONLY if:<br>[conditions_post(2)] = '1' | Enter the first Acute respiratory distress syndrome diagnosis date found during the follow-up period: | text (date_mdy), Required<br>Field Annotation: @HIDEBUTTON |
| 162 | acuteres_date_followup_2<br><br>Show the field ONLY if:<br>[conditions_post(2)] = '1' | Enter the second Acute respiratory distress syndrome diagnosis date found during the follow-up period, if applicable: | text (date_mdy)<br>Field Annotation: @HIDEBUTTON |
| 163 | acuteres_endstatus<br><br>Show the field ONLY if:<br>[conditions_post(2)] = '1' | At the end of the follow-up period, the Acute respiratory distress syndrome was: | dropdown, Required<br>1 Resolved<br>2 Ongoing<br>3 Unknown/Unavailable<br><br>Field Annotation: @HIDEBUTTON |
| 164 | acuteres_diagnosis_pasc<br><br>Show the field ONLY if:<br>[conditions_post(2)] = '1' | Did the physician state in the chart whether the Acute respiratory distress syndrome diagnosis was due to PASC? | dropdown, Required<br>1 Yes, the physician stated this condition was due to PASC<br>2 No, the physician stated that this condition was due to something other than PASC<br>3 Unknown/Not Available<br><br>Field Annotation: @HIDEBUTTON |
| 165 | acuteres_diagnosis_pasc_y<br><br>Show the field ONLY if:<br>[conditions_post(2)] = '1' and [acuteres_diagnosis_pasc] = '1' | Copy and paste the language from the physician notes that states this Acute respiratory distress syndrome diagnosis was related to PASC: | notes, Required<br>Field Annotation: @HIDEBUTTON |
| 166 | losssmell_date_followup<br><br>Show the field ONLY if: | Enter the first Loss of smell diagnosis date found during the follow-up period: | text (date_mdy), Required<br>Field Annotation: @HIDEBUTTON |

| | | | |
|---|---|---|---|
| | [conditions_post(3)] = '1' | | |
| 167 | losssmell_date_followup_2<br><br>Show the field ONLY if:<br>[conditions_post(3)] = '1' | Enter the second Loss of smell diagnosis date found during the follow-up period, if applicable: | text (date_mdy)<br>Field Annotation: @HIDEBUTTON |
| 168 | losssmell_endstatus<br><br>Show the field ONLY if:<br>[conditions_post(3)] = '1' | At the end of the follow-up period, the Loss of smell was: | dropdown, Required<br>1 Resolved<br>2 Ongoing<br>3 Unknown/Unavailable<br><br>Field Annotation: @HIDEBUTTON |
| 169 | losssmell_diagnosis_pasc<br><br>Show the field ONLY if:<br>[conditions_post(3)] = '1' | Did the physician state in the chart whether the Loss of smell diagnosis was due to PASC? | dropdown, Required<br>1 Yes, the physician stated this condition was due to PASC<br>2 No, the physician stated that this condition was due to something other than PASC<br>3 Unknown/Not Available<br><br>Field Annotation: @HIDEBUTTON |
| 170 | losssmell_diagnosis_pasc_y<br><br>Show the field ONLY if:<br>[conditions_post(3)] = '1' and [losssmell_diagnosis_pasc] = '1' | Copy and paste the language from the physician notes that states the Loss of smell diagnosis was related to PASC: | notes, Required<br>Field Annotation: @HIDEBUTTON |
| 171 | losstaste_date_followup<br><br>Show the field ONLY if:<br>[conditions_post(4)] = '1' | Enter the first Loss of taste diagnosis date found during the follow-up period: | text (date_mdy), Required<br>Field Annotation: @HIDEBUTTON |
| 172 | losstaste_date_followup_2<br><br>Show the field ONLY if:<br>[conditions_post(4)] = '1' | Enter the second Loss of taste diagnosis date found during the follow-up period, if applicable: | text (date_mdy)<br>Field Annotation: @HIDEBUTTON |
| 173 | losstaste_endstatus<br><br>Show the field ONLY if:<br>[conditions_post(4)] = '1' | At the end of the follow-up period, the Loss of taste was: | dropdown, Required<br>1 Resolved<br>2 Ongoing<br>3 Unknown/Unavailable<br><br>Field Annotation: @HIDEBUTTON |
| 174 | losstaste_diagnosis_pasc<br><br>Show the field ONLY if: | Did the physician state in the chart whether the Loss of taste diagnosis was due to PASC? | dropdown, Required<br>1 Yes, the physician stated this condition was due to PASC |

| | | | |
|---|---|---|---|
| | [conditions_post(4)] = '1' | | 2 No, the physician stated that this condition was due to something other than PASC<br>3 Unknown/Not Available<br><br>Field Annotation: @HIDEBUTTON |
| 175 | losstaste_diagnosis_pasc_y<br><br>Show the field ONLY if:<br>[conditions_post(4)] = '1' and [losstaste_diagnosis_pasc] = '1' | Copy and paste the language from the physician notes that states the Loss of taste diagnosis was due to PASC: | notes, Required<br>Field Annotation: @HIDEBUTTON |
| 176 | lossother_date_followup<br><br>Show the field ONLY if:<br>[conditions_post(5)] = '1' | Enter the first Other Change of Smell/Taste date found during the follow-up period: | text (date_mdy), Required<br>Field Annotation: @HIDEBUTTON |
| 177 | lossother_date_followup_2<br><br>Show the field ONLY if:<br>[conditions_post(5)] = '1' | Enter the second Other Change of Smell/Taste date found during the follow-up period, if applicable: | text (date_mdy)<br>Field Annotation: @HIDEBUTTON |
| 178 | lossother_endstatus<br><br>Show the field ONLY if:<br>[conditions_post(5)] = '1' | At the end of the follow-up period, the Other Change of Smell/Taste was: | dropdown, Required<br>1 Resolved<br>2 Ongoing<br>3 Unknown/Unavailable<br><br>Field Annotation: @HIDEBUTTON |
| 179 | lossother_diagnosis_pasc<br><br>Show the field ONLY if:<br>[conditions_post(5)] = '1' | Did the physician state in the chart whether the Other Change of Smell/Taste was due to PASC? | dropdown, Required<br>1 Yes, the physician stated this condition was due to PASC<br>2 No, the physician stated that this condition was due to something other than PASC<br>3 Unknown/Not Available<br><br>Field Annotation: @HIDEBUTTON |
| 180 | lossother_diagnosis_pasc_y<br><br>Show the field ONLY if:<br>[conditions_post(5)] = '1' and [lossother_diagnosis_pasc] = '1' | Copy and paste the language from the physician notes that states the Other Change of Smell/Taste was due to PASC: | notes, Required<br>Field Annotation: @HIDEBUTTON |
| 181 | myocarditis_date_followup<br><br>Show the field ONLY if: | Enter the first Myocarditis diagnosis date found during the follow-up period: | text (date_mdy), Required<br>Field Annotation: @HIDEBUTTON |

| | | | |
|---|---|---|---|
| | [conditions_post(6)] = '1' | | |
| 182 | myocarditis_date_followup_2<br><br>Show the field ONLY if:<br>[conditions_post(6)] = '1' | Enter the second Myocarditis diagnosis date found during the follow-up period, if applicable: | text (date_mdy)<br>Field Annotation: @HIDEBUTTON |
| 183 | myocarditis_endstatus<br><br>Show the field ONLY if:<br>[conditions_post(6)] = '1' | At the end of the follow-up period, the Myocarditis was: | dropdown, Required<br>1 Resolved<br>2 Ongoing<br>3 Unknown/Unavailable<br><br>Field Annotation: @HIDEBUTTON |
| 184 | myocarditis_diagnosis_pasc<br><br>Show the field ONLY if:<br>[conditions_post(6)] = '1' | Did the physician state in the notes whether the Myocarditis diagnosis was due to PASC? | dropdown, Required<br>1 Yes, the physician stated this condition was due to PASC<br>2 No, the physician stated that this condition was due to something other than PASC<br>3 Unknown/Not Available<br><br>Field Annotation: @HIDEBUTTON |
| 185 | myocarditis_diagnosis_pasc_y<br><br>Show the field ONLY if:<br>[conditions_post(6)] = '1' and [myocarditis_diagnosis_pasc] = '1' | Copy and paste the language from the physician notes that states the Myocarditis diagnosis was due to PASC: | notes, Required<br>Field Annotation: @HIDEBUTTON |
| 186 | pericarditis_date_followup<br><br>Show the field ONLY if:<br>[conditions_post(7)] = '1' | Enter the first Pericarditis diagnosis date found during the follow-up period: | text (date_mdy), Required<br>Field Annotation: @HIDEBUTTON |
| 187 | pericarditis_date_followup_2<br><br>Show the field ONLY if:<br>[conditions_post(7)] = '1' | Enter the second Pericarditis diagnosis date found during the follow-up period, if applicable: | text (date_mdy)<br>Field Annotation: @HIDEBUTTON |
| 188 | pericarditis_endstatus<br><br>Show the field ONLY if:<br>[conditions_post(7)] = '1' | At the end of the follow-up period, the Pericarditis was: | dropdown, Required<br>1 Resolved<br>2 Ongoing<br>3 Unknown/Unavailable<br><br>Field Annotation: @HIDEBUTTON |
| 189 | pericarditis_diagnosis_pasc<br><br>Show the field ONLY if: | Did the physician state in the notes whether the Pericarditis diagnosis was due to PASC? | dropdown, Required<br>1 Yes, the physician stated this condition was due to PASC |

| | | | |
|---|---|---|---|
| | [conditions_post(7)] = '1' | | 2 No, the physician stated that this condition was due to something other than PASC<br>3 Unknown/Not Available<br><br>Field Annotation: @HIDEBUTTON |
| 190 | pericarditis_diagnosis_pasc_y<br><br>Show the field ONLY if:<br>[conditions_post(7)] = '1' and [pericarditis_diagnosis_pasc] = '1' | Copy and paste the language from the physician notes that states the Pericarditis diagnosis was due to PASC: | notes, Required<br>Field Annotation: @HIDEBUTTON |
| 191 | myositis_date_followup<br><br>Show the field ONLY if:<br>[conditions_post(8)] = '1' | Enter the first Myositis diagnosis date found during the follow-up period: | text (date_mdy), Required<br>Field Annotation: @HIDEBUTTON |
| 192 | myositis_date_followup_2<br><br>Show the field ONLY if:<br>[conditions_post(8)] = '1' | Enter the second Myositis diagnosis date found during the follow-up period, if applicable: | text (date_mdy)<br>Field Annotation: @HIDEBUTTON |
| 193 | myositis_endstatus<br><br>Show the field ONLY if:<br>[conditions_post(8)] = '1' | At the end of the follow-up period, the Myositis was: | dropdown, Required<br>1 Resolved<br>2 Ongoing<br>3 Unknown/Unavailable<br><br>Field Annotation: @HIDEBUTTON |
| 194 | myositis_diagnosis_pasc<br><br>Show the field ONLY if:<br>[conditions_post(8)] = '1' | Did the physician state in the notes whether the Myositis diagnosis was due to PASC? | dropdown, Required<br>1 Yes, the physician stated this condition was due to PASC<br>2 No, the physician stated that this condition was due to something other than PASC<br>3 Unknown/Not Available<br><br>Field Annotation: @HIDEBUTTON |
| 195 | myositis_diagnosis_pasc_y<br><br>Show the field ONLY if:<br>[conditions_post(8)] = '1' and [myositis_diagnosis_pasc] = '1' | Copy and paste the language from the notes that states the Myositis diagnosis was due to PASC: | notes, Required<br>Field Annotation: @HIDEBUTTON |
| 196 | illheart_date_followup<br><br>Show the field ONLY if: | Enter the first Other/ill-defined heart disease diagnosis date found during the follow-up period: | text (date_mdy), Required<br>Field Annotation: @HIDEBUTTON |

| | | | |
|---|---|---|---|
| | [conditions_post(9)] = '1' | | |
| 197 | illheart_date_followup_2<br><br>Show the field ONLY if:<br>[conditions_post(9)] = '1' | Enter the second Other/ill-defined heart disease diagnosis date found during the follow-up period, if applicable: | text (date_mdy)<br>Field Annotation: @HIDEBUTTON |
| 198 | illheart_endstatus<br><br>Show the field ONLY if:<br>[conditions_post(9)] = '1' | At the end of the follow-up period, the Other/ill-defined heart disease was: | dropdown, Required<br>1 Resolved<br>2 Ongoing<br>3 Unknown/Unavailable<br><br>Field Annotation: @HIDEBUTTON |
| 199 | illheart_diagnosis_pasc<br><br>Show the field ONLY if:<br>[conditions_post(9)] = '1' | Did the physician state in the notes whether the Other/ill-defined heart disease diagnosis was due to PASC? | dropdown, Required<br>1 Yes, the physician stated this condition was due to PASC<br>2 No, the physician stated that this condition was due to something other than PASC<br>3 Unknown/Not Available<br><br>Field Annotation: @HIDEBUTTON |
| 200 | illheart_diagnosis_pasc_y<br><br>Show the field ONLY if:<br>[conditions_post(9)] = '1' and [illheart_diagnosis_pasc] = '1' | Copy and paste the language from the notes that states the Other/ill-defined heart disease diagnosis was due to PASC: | notes, Required<br>Field Annotation: @HIDEBUTTON |
| 201 | thrombo_date_followup<br><br>Show the field ONLY if:<br>[conditions_post(10)] = '1' | Enter the first Thrombophlebitis/thromboembolism diagnosis date found during the follow-up period: | text (date_mdy), Required<br>Field Annotation: @HIDEBUTTON |
| 202 | thrombo_date_followup_2<br><br>Show the field ONLY if:<br>[conditions_post(10)] = '1' | Enter the second Thrombophlebitis/thromboembolism diagnosis date found during the follow-up period, if applicable: | text (date_mdy)<br>Field Annotation: @HIDEBUTTON |
| 203 | thrombo_endstatus<br><br>Show the field ONLY if:<br>[conditions_post(10)] = '1' | At the end of the follow-up period, the Thrombophlebitis/thromboembolism was: | dropdown, Required<br>1 Resolved<br>2 Ongoing<br>3 Unknown/Unavailable<br><br>Field Annotation: @HIDEBUTTON |
| 204 | thrombo_diagnosis_p | Did the physician state in the notes whether the | dropdown, Required |

| | | | |
|---|---|---|---|
| | asc<br><br>Show the field ONLY if:<br>[conditions_post(10)] = '1' | Thrombophlebitis/thromboembolism diagnosis was due to PASC? | 1 Yes, the physician stated this condition was due to PASC<br>2 No, the physician stated that this condition was due to something other than PASC<br>3 Unknown/Not Available<br><br>Field Annotation: @HIDEBUTTON |
| 205 | thrombo_diagnosis_pasc_y<br><br>Show the field ONLY if:<br>[conditions_post(10)] = '1' and [thrombo_diagnosis_pasc] = '1' | Copy and paste the language from the notes that states the Thrombophlebitis/thromboembolism diagnosis was due to PASC: | notes, Required<br>Field Annotation: @HIDEBUTTON |
| 206 | anemia_date_followup<br><br>Show the field ONLY if:<br>[conditions_post(11)] = '1' | Enter the first Aplastic anemia diagnosis date found during the follow-up period: | text (date_mdy), Required<br>Field Annotation: @HIDEBUTTON |
| 207 | anemia_date_followup_2<br><br>Show the field ONLY if:<br>[conditions_post(11)] = '1' | Enter the second Aplastic anemia diagnosis date found during the follow-up period, if applicable: | text (date_mdy)<br>Field Annotation: @HIDEBUTTON |
| 208 | anemia_endstatus<br><br>Show the field ONLY if:<br>[conditions_post(11)] = '1' | At the end of the follow-up period, the Aplastic anemia was: | dropdown, Required<br>1 Resolved<br>2 Ongoing<br>3 Unknown/Unavailable<br><br>Field Annotation: @HIDEBUTTON |
| 209 | anemia_diagnosis_pasc<br><br>Show the field ONLY if:<br>[conditions_post(11)] = '1' | Did the physician state in the notes whether the Aplastic anemia diagnosis was due to PASC? | dropdown, Required<br>1 Yes, the physician stated this condition was due to PASC<br>2 No, the physician stated that this condition was due to something other than PASC<br>3 Unknown/Not Available<br><br>Field Annotation: @HIDEBUTTON |
| 210 | anemia_diagnosis_pasc_y<br><br>Show the field ONLY if:<br>[conditions_post(11)] = '1' and [anemia_diagnosis_pasc] = '1' | Copy and paste the language from the notes that states the Aplastic anemia diagnosis was due to PASC: | notes, Required<br>Field Annotation: @HIDEBUTTON |

| | | | |
|---|---|---|---|
| 211 | brainfog_date_followup<br><br>Show the field ONLY if:<br>[conditions_post(12)] = '1' | Enter the first Brain fog diagnosis date found during the follow-up period: | text (date_mdy), Required<br>Field Annotation: @HIDEBUTTON |
| 212 | brainfog_date_followup_2<br><br>Show the field ONLY if:<br>[conditions_post(12)] = '1' | Enter the second Brain fog diagnosis date found during the follow-up period, if applicable: | text (date_mdy)<br>Field Annotation: @HIDEBUTTON |
| 213 | brainfog_endstatus<br><br>Show the field ONLY if:<br>[conditions_post(12)] = '1' | At the end of the follow-up period, the Brain fog was: | dropdown, Required<br>1 Resolved<br>2 Ongoing<br>3 Unknown/Unavailable<br><br>Field Annotation: @HIDEBUTTON |
| 214 | brainfog_diagnosis_pasc<br><br>Show the field ONLY if:<br>[conditions_post(12)] = '1' | Did the physician state in the notes whether the Brain fog diagnosis was due to PASC? | dropdown, Required<br>1 Yes, the physician stated this condition was due to PASC<br>2 No, the physician stated that this condition was due to something other than PASC<br>3 Unknown/Not Available<br><br>Field Annotation: @HIDEBUTTON |
| 215 | brainfog_diagnosis_pasc_y<br><br>Show the field ONLY if:<br>[conditions_post(12)] = '1' and [brainfog_diagnosis_pasc] = '1' | Copy and paste the language from the notes that states the Brain fog diagnosis was due to PASC: | notes, Required<br>Field Annotation: @HIDEBUTTON |
| 216 | liverenzy_date_followup<br><br>Show the field ONLY if:<br>[conditions_post(13)] = '1' | Enter the first Abnormal liver enzymes diagnosis date found during the follow-up period: | text (date_mdy), Required<br>Field Annotation: @HIDEBUTTON |
| 217 | liverenzy_date_followup_2<br><br>Show the field ONLY if:<br>[conditions_post(13)] = '1' | Enter the second Abnormal liver enzymes diagnosis date found during the follow-up period, if applicable: | text (date_mdy)<br>Field Annotation: @HIDEBUTTON |
| 218 | liverenzy_endstatus<br><br>Show the field ONLY if:<br>[conditions_post(13)] = ' | At the end of the follow-up period, the Abnormal liver enzymes were: | dropdown, Required<br>1 Resolved<br>2 Ongoing |

| | | | |
|---|---|---|---|
| | 1' | | 3 Unknown/Unavailable<br><br>Field Annotation: @HIDEBUTTON |
| 219 | liverenzy_diagnosis_pasc<br><br>Show the field ONLY if:<br>[conditions_post(13)] = '1' | Did the physician state in the notes whether the Abnormal liver enzymes diagnosis was due to PASC? | dropdown, Required<br>1 Yes, the physician stated this condition was due to PASC<br>2 No, the physician stated that this condition was due to something other than PASC<br>3 Unknown/Not Available<br><br>Field Annotation: @HIDEBUTTON |
| 220 | liverenzy_diagnosis_pasc_y<br><br>Show the field ONLY if:<br>[conditions_post(13)] = '1' and [liverenzy_diagnosis_pasc] = '1' | Copy and paste the language from the notes that states the Abnormal liver enzymes diagnosis was due to PASC: | notes, Required<br>Field Annotation: @HIDEBUTTON |
| 221 | dysautonomia_date_followup<br><br>Show the field ONLY if:<br>[conditions_post(14)] = '1' | Enter the first Dysautonomia diagnosis date found during the follow-up period: | text (date_mdy), Required<br>Field Annotation: @HIDEBUTTON |
| 222 | dysautonomia_date_followup_2<br><br>Show the field ONLY if:<br>[conditions_post(14)] = '1' | Enter the second Dysautonomia diagnosis date found during the follow-up period, if applicable: | text (date_mdy)<br>Field Annotation: @HIDEBUTTON |
| 223 | dysautonomia_endstatus<br><br>Show the field ONLY if:<br>[conditions_post(14)] = '1' | At the end of the follow-up period, the Dysautonomia was: | dropdown, Required<br>1 Resolved<br>2 Ongoing<br>3 Unknown/Unavailable<br><br>Field Annotation: @HIDEBUTTON |
| 224 | dysautonomia_diagnosis_pasc<br><br>Show the field ONLY if:<br>[conditions_post(14)] = '1' | Did the physician state in the notes whether the Dysautonomia diagnosis was due to PASC? | dropdown, Required<br>1 Yes, the physician stated this condition was due to PASC<br>2 No, the physician stated that this condition was due to something other than PASC<br>3 Unknown/Not Available<br><br>Field Annotation: @HIDEBUTTON |
| 225 | dysautonomia_diagnosis_pasc_y | Copy and paste the language from the notes that states the Dysautonomia diagnosis was due to PASC: | notes, Required<br>Field Annotation: @HIDEBUTTON |

| | | | |
|---|---|---|---|
| | Show the field ONLY if: [conditions_post(14)] = '1' and [dysautonomia_diagnosis_pasc] = '1' | | |
| 226 | fatigue_date_followup<br><br>Show the field ONLY if: [conditions_post(15)] = '1' | Enter the first Fatigue diagnosis date found during the follow-up period: | text (date_mdy), Required<br>Field Annotation: @HIDEBUTTON |
| 227 | fatigue_date_followup_2<br><br>Show the field ONLY if: [conditions_post(15)] = '1' | Enter the second Fatigue diagnosis date found during the follow-up period, if applicable: | text (date_mdy)<br>Field Annotation: @HIDEBUTTON |
| 228 | fatigue_endstatus<br><br>Show the field ONLY if: [conditions_post(15)] = '1' | At the end of the follow-up period, the Fatigue was: | dropdown, Required<br>1 Resolved<br>2 Ongoing<br>3 Unknown/Unavailable<br><br>Field Annotation: @HIDEBUTTON |
| 229 | fatigue_diagnosis_pasc<br><br>Show the field ONLY if: [conditions_post(15)] = '1' | Did the physician state in the notes whether the Fatigue diagnosis was due to PASC? | dropdown, Required<br>1 Yes, the physician stated this condition was due to PASC<br>2 No, the physician stated that this condition was due to something other than PASC<br>3 Unknown/Not Available<br><br>Field Annotation: @HIDEBUTTON |
| 230 | fatigue_diagnosis_pasc_y<br><br>Show the field ONLY if: [conditions_post(15)] = '1' and [fatigue_diagnosis_pasc] = '1' | Copy and paste the language from the notes that states the Fatigue diagnosis was due to PASC: | notes, Required<br>Field Annotation: @HIDEBUTTON |
| 231 | abdominal_date_followup<br><br>Show the field ONLY if: [conditions_post(19)] = '1' | Enter the first Abdominal pain diagnosis date found during the follow-up period: | text (date_mdy), Required<br>Field Annotation: @HIDEBUTTON |
| 232 | abdominal_date_followup_2<br><br>Show the field ONLY if: [conditions_post(19)] = '1' | Enter the second Abdominal pain diagnosis date found during the follow-up period, if applicable: | text (date_mdy)<br>Field Annotation: @HIDEBUTTON |

| | | | |
|---|---|---|---|
| 233 | abdominal_endstatus<br>Show the field ONLY if:<br>[conditions_post(19)] = '1' | At the end of the follow-up period, the Abdominal Pain was: | dropdown, Required<br>1 Resolved<br>2 Ongoing<br>3 Unknown/Unavailable<br>Field Annotation: @HIDEBUTTON |
| 234 | abdominal_diagnosis_pasc<br>Show the field ONLY if:<br>[conditions_post(19)] = '1' | Did the physician state in the notes whether the Abdominal Pain diagnosis was due to PASC? | dropdown, Required<br>1 Yes, the physician stated this condition was due to PASC<br>2 No, the physician stated that this condition was due to something other than PASC<br>3 Unknown/Not Available<br>Field Annotation: @HIDEBUTTON |
| 235 | abdominal_diagnosis_pasc_y<br>Show the field ONLY if:<br>[conditions_post(19)] = '1' and [abdominal_diagnosis_pasc] = '1' | Copy and paste the language from the notes that states the Abdominal Pain diagnosis was due to PASC: | notes, Required<br>Field Annotation: @HIDEBUTTON |
| 236 | aki_date_followup<br>Show the field ONLY if:<br>[conditions_post(20)] = '1' | Enter the first Acute Kidney Injury diagnosis date found during the follow-up period: | text (date_mdy), Required<br>Field Annotation: @HIDEBUTTON |
| 237 | aki_date_followup_2<br>Show the field ONLY if:<br>[conditions_post(20)] = '1' | Enter the second Acute Kidney Injury diagnosis date found during the follow-up period, if applicable: | text (date_mdy)<br>Field Annotation: @HIDEBUTTON |
| 238 | aki_endstatus<br>Show the field ONLY if:<br>[conditions_post(20)] = '1' | At the end of the follow-up period, the Acute Kidney Injury was: | dropdown, Required<br>1 Resolved<br>2 Ongoing<br>3 Unknown/Unavailable<br>Field Annotation: @HIDEBUTTON |
| 239 | aki_diagnosis_pasc<br>Show the field ONLY if:<br>[conditions_post(20)] = '1' | Did the physician state in the notes whether the Acute Kidney Injury diagnosis was due to PASC? | dropdown, Required<br>1 Yes, the physician stated this condition was due to PASC<br>2 No, the physician stated that this condition was due to something other than PASC<br>3 Unknown/Not Available |

| | | | |
|---|---|---|---|
| | | | Field Annotation: @HIDEBUTTON |
| 240 | aki_diagnosis_pasc_y<br><br>Show the field ONLY if:<br>[conditions_post(20)] = '1' and [aki_diagnosis_pasc] = '1' | Copy and paste the language from the notes that states the Acute Kidney Injury diagnosis was due to PASC: | notes, Required<br>Field Annotation: @HIDEBUTTON |
| 241 | abnormal_lab<br><br>Show the field ONLY if:<br>[covid] = '1' | Did the patient have at least one occurrence of the following abnormal lab tests, on Day 28 or later after initial COVID-19 positive diagnosis?<br><br>Select all that apply. | checkbox, Required<br>1 abnormal_lab___1 Thrombocytopenia<br>2 abnormal_lab___2 Elevated Troponin<br>3 abnormal_lab___3 Lymphopenia<br>4 abnormal_lab___4 Elevated CRP<br>5 abnormal_lab___5 The patient never had any of these tests<br>6 abnormal_lab___6 The patient had a test but it was done prior to Day 28 post COVID positive diagnosis<br>7 abnormal_lab___7 The patient had a test Day 28 or later, but the test results were normal |

**Instrument: RECOVER Pediatric Chart Review Form** (recover_pediatric_chart_review_form)

| | | | |
|---|---|---|---|
| 242 | reason_thrombo_test<br><br>Show the field ONLY if:<br>[covid] = '1' and [abnormal_lab(1)] = '1' | Select the reason why the patient was tested for Thrombocytopenia: | dropdown, Required<br>1 Symptomatic<br>2 Asymptomatic<br>3 Concern for PASC<br>4 Exposed<br>5 Other<br>6 Unsure/Not Available |
| 243 | date_thrombo_test<br><br>Show the field ONLY if:<br>[covid] = '1' and [abnormal_lab(1)] = '1' | Enter the Thrombocytopenia test date: | text (date_mdy), Required<br>Field Annotation: @HIDEBUTTON |
| 244 | reason_troponin_test<br><br>Show the field ONLY if:<br>[covid] = '1' and [abnormal_lab(2)] = '1' | Select the reason why the patient was tested for Elevated Troponin: | dropdown, Required<br>1 Symptomatic<br>2 Asymptomatic<br>3 Concern for PASC<br>4 Exposed<br>5 Other |

| | | | |
|---|---|---|---|
| | | | 6 Unsure/Not Available |
| 245 | date_troponin_test<br>Show the field ONLY if:<br>[covid] = '1' and [abnormal_lab(2)] = '1' | Enter the Elevated Troponin test date: | text (date_mdy), Required<br>Field Annotation: @HIDEBUTTON |
| 246 | reason_lymphopenia_test<br>Show the field ONLY if:<br>[covid] = '1' and [abnormal_lab(3)] = '1' | Select the reason why the patient was tested for Lymphopenia: | dropdown, Required<br>1 Symptomatic<br>2 Asymptomatic<br>3 Concern for PASC<br>4 Exposed<br>5 Other<br>6 Unsure/Not Available |
| 247 | date_lymphopenia_test<br>Show the field ONLY if:<br>[covid] = '1' and [abnormal_lab(3)] = '1' | Enter the Lymphopenia test date: | text (date_mdy), Required<br>Field Annotation: @HIDEBUTTON |
| 248 | reason_crp_test<br>Show the field ONLY if:<br>[covid] = '1' and [abnormal_lab(4)] = '1' | Select the reason why the patient was tested for Elevated CRP: | dropdown, Required<br>1 Symptomatic<br>2 Asymptomatic<br>3 Concern for PASC<br>4 Exposed<br>5 Other<br>6 Unsure/Not Available |
| 249 | date_crp_test<br>Show the field ONLY if:<br>[covid] = '1' and [abnormal_lab(4)] = '1' | Enter the Elevated CRP test date: | text (date_mdy), Required<br>Field Annotation: @HIDEBUTTON |
| 250 | diagnosis_post_covid<br>Show the field ONLY if:<br>[covid] = '1' | Did the patient have at least one occurrences of a diagnosis term for the following on Day 28 or later after COVID positive diagnosis?<br><br>Select all that apply. | checkbox<br>1 diagnosis_post_covid___1 Chest pain<br>2 diagnosis_post_covid___2 Hair loss<br>3 diagnosis_post_covid___3 Cough<br>4 diagnosis_post_covid___4 Cardiorespirat signs and symptoms<br>5 diagnosis_post_covid___5 Jaundice<br>6 diagnosis_post_covid___6 Generalized p<br>7 diagnosis_post_covid___7 Anxiety symptoms<br>8 diagnosis_post_covid___8 Fatigue/malais |

| | | | |
|---|---|---|---|
| | | | 9 diagnosis_post_covid___9 Diarrhea<br>10 diagnosis_post_covid___10 Fever/chills<br>11 diagnosis_post_covid___11 Skin rashes<br>12 diagnosis_post_covid___12 Headache<br>13 diagnosis_post_covid___13 None of the above<br>14 diagnosis_post_covid___14 The patient ha[illegible] condition liste[illegible] above but it occurred prior[illegible] Day 28 post COVID positive diagnosis |
| 251 | cardiorespiratory<br>Show the field ONLY if:<br>[diagnosis_post_covid(4)] = '1' | Describe or list the relevant cardiorespiratory signs and symptoms: | notes, Required |
| 252 | chestpain_date<br>Show the field ONLY if:<br>[covid] = '1' and [diagnosis_post_covid(1)] = '1' | Enter the date of the initial Chest Pain diagnosis: | text (date_mdy), Required<br>Field Annotation: @HIDEBUTTON |
| 253 | chestpain_date_2<br>Show the field ONLY if:<br>[covid] = '1' and [diagnosis_post_covid(1)] = '1' | Enter the date of the second Chest Pain diagnosis, if applicable: | text (date_mdy)<br>Field Annotation: @HIDEBUTTON |
| 254 | chestpain_endstatus<br>Show the field ONLY if:<br>[covid] = '1' and [diagnosis_post_covid(1)] = '1' | At the end of the follow-up period, the Chest Pain was: | dropdown, Required<br>1 Resolved<br>2 Ongoing<br>3 Unknown/Unavailable<br>Field Annotation: @HIDEBUTTON |
| 255 | chestpain_pasc<br>Show the field ONLY if:<br>[covid] = '1' and [diagnosis_post_covid(1)] = '1' | Did the physician state in the notes whether the Chest Pain was related to PASC? | dropdown, Required<br>1 Yes, the physician stated this condition was due to PASC<br>2 No, the physician stated that this condition was due to something other than PASC<br>3 Unknown/Not Available<br>Field Annotation: @HIDEBUTTON |
| 256 | chestpain_pasc_y<br>Show the field ONLY if:<br>[covid] = '1' and [diagnos | Copy and paste the language from the notes that states the Chest Pain diagnosis was related to PASC: | notes, Required<br>Field Annotation: @HIDEBUTTON |

| | | | |
|---|---|---|---|
| | is_post_covid(1)] = '1' and [chestpain_pasc] = '1' | | |
| 257 | hairloss_date<br><br>Show the field ONLY if:<br>[covid] = '1' and [diagnosis_post_covid(2)] = '1' | Enter the date of the initial Hair Loss diagnosis: | text (date_mdy), Required<br>Field Annotation: @HIDEBUTTON |
| 258 | hairloss_date_2<br><br>Show the field ONLY if:<br>[covid] = '1' and [diagnosis_post_covid(2)] = '1' | Enter the date of the second Hair Loss diagnosis, if applicable: | text (date_mdy)<br>Field Annotation: @HIDEBUTTON |
| 259 | hairloss_endstatus<br><br>Show the field ONLY if:<br>[covid] = '1' and [diagnosis_post_covid(2)] = '1' | At the end of the follow-up period, the Hair Loss was: | dropdown, Required<br>1 Resolved<br>2 Ongoing<br>3 Unknown/Unavailable<br><br>Field Annotation: @HIDEBUTTON |
| 260 | hairloss_pasc<br><br>Show the field ONLY if:<br>[covid] = '1' and [diagnosis_post_covid(2)] = '1' | Did the physician state in the notes whether the Hair Loss was related to PASC? | dropdown, Required<br>1 Yes, the physician stated this condition was due to PASC<br>2 No, the physician stated that this condition was due to something other than PASC<br>3 Unknown/Not Available<br><br>Field Annotation: @HIDEBUTTON |
| 261 | hairloss_pasc_y<br><br>Show the field ONLY if:<br>[covid] = '1' and [diagnosis_post_covid(2)] = '1' and [hairloss_pasc] = '1' | Copy and paste the language from the notes that states the Hair Loss diagnosis was related to PASC: | notes, Required<br>Field Annotation: @HIDEBUTTON |
| 262 | cough_date<br><br>Show the field ONLY if:<br>[covid] = '1' and [diagnosis_post_covid(3)] = '1' | Enter the date of the initial Cough diagnosis: | text (date_mdy), Required<br>Field Annotation: @HIDEBUTTON |
| 263 | cough_date_2<br><br>Show the field ONLY if:<br>[covid] = '1' and [diagnosis_post_covid(3)] = '1' | Enter the date of the second Cough diagnosis, if applicable: | text (date_mdy)<br>Field Annotation: @HIDEBUTTON |
| 264 | cough_endstatus<br><br>Show the field ONLY if:<br>[covid] = '1' and [diagnosis_post_covid(3)] = '1' | At the end of the follow-up period, the Cough was: | dropdown, Required<br>1 Resolved<br>2 Ongoing<br>3 Unknown/Unavailable |

| | | | |
|---|---|---|---|
| | | | Field Annotation: @HIDEBUTTON |
| 265 | cough_pasc<br><br>Show the field ONLY if:<br>[covid] = '1' and [diagnos is_post_covid(3)] = '1' | Did the physician state in the notes whether the Cough was related to PASC? | dropdown, Required<br>1 Yes, the physician stated this condition was due to PASC<br>2 No, the physician stated that this condition was due to something other than PASC<br>3 Unknown/Not Available<br><br>Field Annotation: @HIDEBUTTON |
| 266 | cough_pasc_y<br><br>Show the field ONLY if:<br>[covid] = '1' and [diagnos is_post_covid(3)] = '1' an d [cough_pasc] = '1' | Copy and paste the language from the notes that states the Cough diagnosis was related to PASC: | notes, Required<br>Field Annotation: @HIDEBUTTON |
| 267 | cardio_signs_date<br><br>Show the field ONLY if:<br>[covid] = '1' and [diagnos is_post_covid(4)] = '1' | Enter the date of the initial Cardiorespiratory signs and symptoms: | text (date_mdy), Required<br>Field Annotation: @HIDEBUTTON |
| 268 | cardio_signs_date_ 2<br><br>Show the field ONLY if:<br>[covid] = '1' and [diagnos is_post_covid(4)] = '1' | Enter the date of the second Cardiorespiratory signs and symptoms, if applicable: | text (date_mdy)<br>Field Annotation: @HIDEBUTTON |
| 269 | cardioresp_endstatu s<br><br>Show the field ONLY if:<br>[covid] = '1' and [diagnos is_post_covid(4)] = '1' | At the end of the follow-up period, the Cardiorespiratory signs and symptoms were: | dropdown, Required<br>1 Resolved<br>2 Ongoing<br>3 Unknown/Unavailable<br><br>Field Annotation: @HIDEBUTTON |
| 270 | cardioresp_pasc<br><br>Show the field ONLY if:<br>[covid] = '1' and [diagnos is_post_covid(4)] = '1' | Did the physician state in the notes whether the Cardiorespiratory signs and symptoms were related to PASC? | dropdown, Required<br>1 Yes, the physician stated this condition was due to PASC<br>2 No, the physician stated that this condition was due to something other than PASC<br>3 Unknown/Not Available<br><br>Field Annotation: @HIDEBUTTON |
| 271 | cardioresp_pasc_y<br><br>Show the field ONLY if:<br>[covid] = '1' and [diagnos is_post_covid(4)] = '1' an d [cardioresp_pasc] = '1' | Copy and paste the language from the notes that states the Cardiorespiratory signs and symptoms were related to PASC: | notes, Required<br>Field Annotation: @HIDEBUTTON |

| | | | |
|---|---|---|---|
| 272 | jaundice_date<br><br>Show the field ONLY if:<br>[covid] = '1' and [diagnos is_post_covid(5)] = '1' | Enter the date of the initial Jaundice diagnosis: | text (date_mdy), Required<br>Field Annotation: @HIDEBUTTON |
| 273 | jaundice_date_2<br><br>Show the field ONLY if:<br>[covid] = '1' and [diagnos is_post_covid(5)] = '1' | Enter the date of the second Jaundice diagnosis, if applicable: | text (date_mdy)<br>Field Annotation: @HIDEBUTTON |
| 274 | jaundice_endstatus<br><br>Show the field ONLY if:<br>[covid] = '1' and [diagnos is_post_covid(5)] = '1' | At the end of the follow-up period, the Jaundice was: | dropdown, Required<br>1 Resolved<br>2 Ongoing<br>3 Unknown/Unavailable<br><br>Field Annotation: @HIDEBUTTON |
| 275 | jaundice_pasc<br><br>Show the field ONLY if:<br>[covid] = '1' and [diagnos is_post_covid(5)] = '1' | Did the physician state in the notes whether the Jaundice was related to PASC? | dropdown, Required<br>1 Yes, the physician stated this condition was due to PASC<br>2 No, the physician stated that this condition was due to something other than PASC<br>3 Unknown/Not Available<br><br>Field Annotation: @HIDEBUTTON |
| 276 | jaundice_pasc_y<br><br>Show the field ONLY if:<br>[covid] = '1' and [diagnos is_post_covid(5)] = '1' an d [jaundice_pasc] = '1' | Copy and paste the language from the notes that states the Jaundice was related to PASC: | notes, Required<br>Field Annotation: @HIDEBUTTON |
| 277 | pain_date<br><br>Show the field ONLY if:<br>[covid] = '1' and [diagnos is_post_covid(6)] = '1' | Enter the date of the initial Generalized Pain diagnosis: | text (date_mdy), Required<br>Field Annotation: @HIDEBUTTON |
| 278 | pain_date_2<br><br>Show the field ONLY if:<br>[covid] = '1' and [diagnos is_post_covid(6)] = '1' | Enter the date of the second Generalized Pain diagnosis, if applicable: | text (date_mdy)<br>Field Annotation: @HIDEBUTTON |
| 279 | pain_endstatus<br><br>Show the field ONLY if:<br>[covid] = '1' and [diagnos is_post_covid(6)] = '1' | At the end of the follow-up period, the Generalized Pain was: | dropdown, Required<br>1 Resolved<br>2 Ongoing<br>3 Unknown/Unavailable<br><br>Field Annotation: @HIDEBUTTON |
| 280 | pain_pasc | Did the physician state in the notes whether the | dropdown, Required |

| | | | |
|---|---|---|---|
| | Show the field ONLY if:<br>[covid] = '1' and [diagnosis_post_covid(6)] = '1' | Generalized Pain was related to PASC? | 1 Yes, the physician stated this condition was due to PASC<br>2 No, the physician stated that this condition was due to something other than PASC<br>3 Unknown/Not Available<br><br>Field Annotation: @HIDEBUTTON |
| 281 | pain_pasc_y<br><br>Show the field ONLY if:<br>[covid] = '1' and [diagnosis_post_covid(6)] = '1' and [pain_pasc] = '1' | Copy and paste the language from the notes that states the Generalized Pain was related to PASC: | notes, Required<br>Field Annotation: @HIDEBUTTON |
| 282 | anxiety_date<br><br>Show the field ONLY if:<br>[covid] = '1' and [diagnosis_post_covid(7)] = '1' | Enter the date of the initial Anxiety symptoms: | text (date_mdy), Required<br>Field Annotation: @HIDEBUTTON |
| 283 | anxiety_date_2<br><br>Show the field ONLY if:<br>[covid] = '1' and [diagnosis_post_covid(7)] = '1' | Enter the date of the second Anxiety symptoms, if applicable: | text (date_mdy)<br>Field Annotation: @HIDEBUTTON |
| 284 | anxiety_endstatus<br><br>Show the field ONLY if:<br>[covid] = '1' and [diagnosis_post_covid(7)] = '1' | At the end of the follow-up period, the Anxiety Symptoms were: | dropdown, Required<br>1 Resolved<br>2 Ongoing<br>3 Unknown/Unavailable<br><br>Field Annotation: @HIDEBUTTON |
| 285 | anxiety_pasc<br><br>Show the field ONLY if:<br>[covid] = '1' and [diagnosis_post_covid(7)] = '1' | Did the physician state in the notes whether the Anxiety Symptoms were related to PASC? | dropdown, Required<br>1 Yes, the physician stated this condition was due to PASC<br>2 No, the physician stated that this condition was due to something other than PASC<br>3 Unknown/Not Available<br><br>Field Annotation: @HIDEBUTTON |
| 286 | anxiety_pasc_y<br><br>Show the field ONLY if:<br>[covid] = '1' and [diagnosis_post_covid(7)] = '1' and [anxiety_pasc] = '1' | Copy and paste the language from the notes that states the Anxiety symptoms were related to PASC: | notes, Required<br>Field Annotation: @HIDEBUTTON |
| 287 | fatigue_malaise_date<br><br>Show the field ONLY if: | Enter the date of the initial Fatigue/Malaise diagnosis: | text (date_mdy), Required<br>Field Annotation: @HIDEBUTTON |

| | | | |
|---|---|---|---|
| | [covid] = '1' and [diagnosis_post_covid(8)] = '1' | | |
| 288 | fatigue_malaise_date_2<br><br>Show the field ONLY if:<br>[covid] = '1' and [diagnosis_post_covid(8)] = '1' | Enter the date of the second Fatigue/Malaise diagnosis, if applicable: | text (date_mdy)<br>Field Annotation: @HIDEBUTTON |
| 289 | fatigue_malaise_endstatus<br><br>Show the field ONLY if:<br>[covid] = '1' and [diagnosis_post_covid(8)] = '1' | At the end of the follow-up period, the Fatigue/Malaise Symptoms were: | dropdown, Required<br>1 Resolved<br>2 Ongoing<br>3 Unknown/Unavailable<br><br>Field Annotation: @HIDEBUTTON |
| 290 | fatigue_malaise_pasc<br><br>Show the field ONLY if:<br>[covid] = '1' and [diagnosis_post_covid(8)] = '1' | Did the physician state in the notes whether the Fatigue/Malaise Symptoms were related to PASC? | dropdown, Required<br>1 Yes, the physician stated this condition was due to PASC<br>2 No, the physician stated that this condition was due to something other than PASC<br>3 Unknown/Not Available<br><br>Field Annotation: @HIDEBUTTON |
| 291 | fatigue_malaise_pasc_y<br><br>Show the field ONLY if:<br>[covid] = '1' and [diagnosis_post_covid(8)] = '1' and [fatigue_malaise_pasc] = '1' | Copy and paste the language from the notes that states the Fatigue/Malaise symptoms were related to PASC: | notes, Required<br>Field Annotation: @HIDEBUTTON |
| 292 | diarrhea_date<br><br>Show the field ONLY if:<br>[covid] = '1' and [diagnosis_post_covid(9)] = '1' | Enter the date of the initial Diarrhea diagnosis: | text (date_mdy), Required<br>Field Annotation: @HIDEBUTTON |
| 293 | diarrhea_date_2<br><br>Show the field ONLY if:<br>[covid] = '1' and [diagnosis_post_covid(9)] = '1' | Enter the date of the second Diarrhea diagnosis, if applicable: | text (date_mdy)<br>Field Annotation: @HIDEBUTTON |
| 294 | diarrhea_endstatus<br><br>Show the field ONLY if:<br>[covid] = '1' and [diagnosis_post_covid(9)] = '1' | At the end of the follow-up period, the Diarrhea was: | dropdown, Required<br>1 Resolved<br>2 Ongoing<br>3 Unknown/Unavailable<br><br>Field Annotation: @HIDEBUTTON |
| 295 | diarrhea_pasc | Did the physician state in the notes whether the | dropdown, Required |

| | | | |
|---|---|---|---|
| | Show the field ONLY if: [covid] = '1' and [diagnosis_post_covid(9)] = '1' | Diarrhea was related to PASC? | 1 Yes, the physician stated this condition was due to PASC<br>2 No, the physician stated that this condition was due to something other than PASC<br>3 Unknown/Not Available<br><br>Field Annotation: @HIDEBUTTON |
| 296 | diarrhea_pasc_y<br><br>Show the field ONLY if: [covid] = '1' and [diagnosis_post_covid(9)] = '1' and [diarrhea_pasc] = '1' | Copy and paste the language from the notes that states the Diarrhea was related to PASC: | notes, Required<br>Field Annotation: @HIDEBUTTON |
| 297 | fever_chills_date<br><br>Show the field ONLY if: [covid] = '1' and [diagnosis_post_covid(10)] = '1' | Enter the date of the initial Fever/Chills diagnosis: | text (date_mdy), Required<br>Field Annotation: @HIDEBUTTON |
| 298 | fever_chills_date_2<br><br>Show the field ONLY if: [covid] = '1' and [diagnosis_post_covid(10)] = '1' | Enter the date of the second Fever/Chills diagnosis, if applicable: | text (date_mdy)<br>Field Annotation: @HIDEBUTTON |
| 299 | fever_chills_endstatus<br><br>Show the field ONLY if: [covid] = '1' and [diagnosis_post_covid(10)] = '1' | At the end of the follow-up period, the Fever/Chills were: | dropdown, Required<br>1 Resolved<br>2 Ongoing<br>3 Unknown/Unavailable<br><br>Field Annotation: @HIDEBUTTON |
| 300 | fever_chills_pasc<br><br>Show the field ONLY if: [covid] = '1' and [diagnosis_post_covid(10)] = '1' | Did the physician state in the notes whether the Fever/Chills were related to PASC? | dropdown, Required<br>1 Yes, the physician stated this condition was due to PASC<br>2 No, the physician stated that this condition was due to something other than PASC<br>3 Unknown/Not Available<br><br>Field Annotation: @HIDEBUTTON |
| 301 | fever_chills_pasc_y<br><br>Show the field ONLY if: [covid] = '1' and [diagnosis_post_covid(10)] = '1' and [fever_chills_pasc] = '1' | Copy and paste the language from the notes that states the Fever/Chills were related to PASC: | notes, Required<br>Field Annotation: @HIDEBUTTON |

| | | | |
|---|---|---|---|
| 302 | skin_rash_date<br><br>Show the field ONLY if:<br>[covid] = '1' and [diagnos is_post_covid(11)] = '1' | Enter the date of the initial Skin Rash diagnosis: | text (date_mdy), Required<br>Field Annotation: @HIDEBUTTON |
| 303 | skin_rash_date_2<br><br>Show the field ONLY if:<br>[covid] = '1' and [diagnos is_post_covid(11)] = '1' | Enter the date of the second Skin Rash diagnosis, if applicable: | text (date_mdy)<br>Field Annotation: @HIDEBUTTON |
| 304 | skinrash_endstatus<br><br>Show the field ONLY if:<br>[covid] = '1' and [diagnos is_post_covid(11)] = '1' | At the end of the follow-up period, the Skin Rash was: | dropdown, Required<br>1 Resolved<br>2 Ongoing<br>3 Unknown/Unavailable<br><br>Field Annotation: @HIDEBUTTON |
| 305 | skin_rash_pasc<br><br>Show the field ONLY if:<br>[covid] = '1' and [diagnos is_post_covid(11)] = '1' | Did the physician state in the notes whether the Skin Rash was related to PASC? | dropdown, Required<br>1 Yes, the physician stated this condition was due to PASC<br>2 No, the physician stated that this condition was due to something other than PASC<br>3 Unknown/Not Available<br><br>Field Annotation: @HIDEBUTTON |
| 306 | skin_rash_pasc_y<br><br>Show the field ONLY if:<br>[covid] = '1' and [diagnos is_post_covid(11)] = '1' a nd [skin_rash_pasc] = '1' | Copy and paste the language from the notes that states the Skin Rash was related to PASC: | notes, Required<br>Field Annotation: @HIDEBUTTON |
| 307 | headache_date<br><br>Show the field ONLY if:<br>[covid] = '1' and [diagnos is_post_covid(12)] = '1' | Enter the date of the initial Headache diagnosis: | text (date_mdy), Required<br>Field Annotation: @HIDEBUTTON |
| 308 | headache_date_2<br><br>Show the field ONLY if:<br>[covid] = '1' and [diagnos is_post_covid(12)] = '1' | Enter the date of the second Headache diagnosis, if applicable: | text (date_mdy)<br>Field Annotation: @HIDEBUTTON |
| 309 | headache_endstatus<br><br>Show the field ONLY if:<br>[covid] = '1' and [diagnos is_post_covid(12)] = '1' | At the end of the follow-up period, the Headache was: | dropdown, Required<br>1 Resolved<br>2 Ongoing<br>3 Unknown/Unavailable<br><br>Field Annotation: @HIDEBUTTON |
| 310 | headache_pasc | Did the physician state in the notes whether the Headache was related to PASC? | dropdown, Required |

| | | | |
|---|---|---|---|
| | Show the field ONLY if: [covid] = '1' and [diagnosis_post_covid(12)] = '1' | | 1 Yes, the physician stated this condition was due to PASC<br>2 No, the physician stated that this condition was due to something other than PASC<br>3 Unknown/Not Available<br><br>Field Annotation: @HIDEBUTTON |
| 311 | headache_pasc_y<br><br>Show the field ONLY if: [covid] = '1' and [diagnosis_post_covid(12)] = '1' and [headache_pasc] = '1' | Copy and paste the language from the notes that states the Headache was related to PASC: | notes, Required<br>Field Annotation: @HIDEBUTTON |
| 312 | other_pasc | Section Header: *Section 7. Additional Comments*<br><br>Is there any additional information in the patient's chart about their PASC status that you think is relevant? For example, if this patient did not have PASC, enter any information you saw that might explain why they were flagged as a PASC patient (such as references to PASC or related conditions that were ruled out).<br><br>If so, please enter below, ensuring that any PHI (names, MRNs, etc) is removed. | notes, Required |
| 313 | care_everywhere | Is there any additional information in Care Everywhere about this patient's COVID or PASC status? | notes, Required |
| 314 | optional_notes | Optional- add any additional comments about this patient's chart. | notes |
| 315 | recover_pediatric_chart_review_form_complete | Section Header: *Form Status*<br><br>Complete? | dropdown<br>0 Incomplete<br>1 Unverified<br>2 Complete |
| **Instrument: Clinician Adjudication (clinician_adjudication)** | | | |
| 316 | covid_adj | Section Header: *Overview of Chart Reviewer Results*<br><br>Does this patient have COVID-19?[covid] | descriptive |
| 317 | coviddiagnosis_date_adj<br><br>Show the field ONLY if: [covid] = '1' | Enter the date the first COVID-19 diagnosis was made: [coviddiagnosis_date] | descriptive |
| 318 | coviddiagnosis_method_adj<br><br>Show the field ONLY if: [covid] = '1' | How was the diagnosis made? [coviddiagnosis_method] | descriptive |
| 319 | coviddiagnosis_mult | Did the patient have repeated COVID-19 (more than | descriptive |

| | | | |
|---|---|---|---|
| | iple_adj<br><br>Show the field ONLY if:<br>[covid] = '1' | one SARS-CoV-2 infection)?[coviddiagnosis_multiple] | |
| 320 | coviddiagnosis_multiple_dates_adj<br><br>Show the field ONLY if:<br>[covid] = '1' and [coviddiagnosis_multiple] = '1' | Additional COVID diagnosis dates: [coviddiagnosis_date_2] [coviddiagnosis_date_3] [coviddiagnosis_date_4] [coviddiagnosis_date_5] | descriptive |
| 321 | misc_diagnosis_adj<br><br>Show the field ONLY if:<br>[covid] = '1' | Are there any references to the patient being diagnosed with MIS-C at any time during study period (Index Date up to 1 year after last COVID-19 diagnosis date)?[misc_diagnosis] | descriptive |
| 322 | misc_terms_adj<br><br>Show the field ONLY if:<br>[covid] = '1' and [misc_diagnosis] = '1' | Enter the MIS-C diagnoses terms in the patient's chart:[misc_terms] | descriptive |
| 323 | misc_date_adj<br><br>Show the field ONLY if:<br>[covid] = '1' and [misc_diagnosis] = '1' | First MIS-C diagnosis date:[misc_date] | descriptive |
| 324 | misc_date_adj_2<br><br>Show the field ONLY if:<br>[covid] = '1' and [misc_diagnosis] = '1' | Second MIS-C diagnosis date (when applicable): [misc_date_2] | descriptive |
| 325 | u099_diagnosis_adj<br><br>Show the field ONLY if:<br>[covid] = '1' | Did the patient have any references to a diagnosis for "PASC" at any time during study period? [u099_diagnosis] | descriptive |
| 326 | pasc_terms_adj<br><br>Show the field ONLY if:<br>[covid] = '1' and [u099_diagnosis] = '1' | Enter the PASC diagnoses references in the patient's chart:[pasc_terms] | descriptive |
| 327 | pasc_date_adj<br><br>Show the field ONLY if:<br>[covid] = '1' and [u099_diagnosis] = '1' | Enter the first PASC diagnosis date:[pasc_date] | descriptive |
| 328 | pasc_date_adj_2<br><br>Show the field ONLY if:<br>[covid] = '1' and [u099_diagnosis] = '1' | Second PASC diagnosis date (when applicable): [pasc_date_2] | descriptive |
| 329 | pcr_antigen_positive_adj<br><br>Show the field ONLY if:<br>[covid] = '1' | Did the patient have a SARS-CoV-2 RT PCR/ antigen test positive during the study period? [pcr_antigen_positive] | descriptive |
| 330 | kawasaki_diagnosis_ | Did the patient have a diagnosis for Kawasaki | descriptive |

| | | | |
|---|---|---|---|
| | adj<br><br>Show the field ONLY if:<br>[covid] = '1' | disease assigned within 42 days of a positive RT-PCR/Antigen test?[kawasaki_diagnosis] | |
| 331 | kawasaki_terms_adj<br><br>Show the field ONLY if:<br>[covid] = '1' and [kawasaki_diagnosis] = '1' | Enter the diagnosis terms for Kawasaki Disease found in the patient chart.[kawasaki_terms] | descriptive |
| 332 | serology_test_adj<br><br>Show the field ONLY if:<br>[covid] = '1' | Did the patient have SARS-CoV-2 serology positive test result?[serology_test] | descriptive |
| 333 | serology_test_reason_adj<br><br>Show the field ONLY if:<br>[covid] = '1' and ([serology_test] = '1' or [serology_test] = '3' or [serology_test] = '4') | Select the reason why the patient was given a serology test:[serology_test_reason] | descriptive |
| 334 | serology_test_date_adj<br><br>Show the field ONLY if:<br>[covid] = '1' and ([serology_test] = '1' or [serology_test] = '3' or [serology_test] = '4') | Enter the date of the initial serology test:[serology_test_date] | descriptive |
| 335 | conditions_post_adj<br><br>Show the field ONLY if:<br>[covid] = '1' | Did the patient have any occurrence of the following conditions, on Day 28 or later after initial COVID-19 positive diagnosis? Select all that apply.[conditions_post] | descriptive |
| 336 | pasc_other_adj<br><br>Show the field ONLY if:<br>[conditions_post(18)] = '1' | List the Other conditions and first 2 diagnosis dates of each condition:[pasc_other] | descriptive, Required<br>Field Annotation: @HIDEBUTTON |
| 337 | covid_date_followup_adj<br><br>Show the field ONLY if:<br>[conditions_post(1)] = '1' | Enter the first COVID diagnosis date found during the follow-up period:[covid_date_followup] | descriptive, Required<br>Field Annotation: @HIDEBUTTON |
| 338 | covid_endstatus_adj<br><br>Show the field ONLY if:<br>[conditions_post(1)] = '1' | At the end of the follow-up period, the COVID diagnosis was:[covid_endstatus] | descriptive, Required<br>Field Annotation: @HIDEBUTTON |
| 339 | covid_diagnosis_pasc_adj<br><br>Show the field ONLY if:<br>[conditions_post(1)] = '1' | Did the physician state in the chart whether this continued COVID diagnosis was due to PASC?[covid_diagnosis_pasc] | descriptive, Required<br>Field Annotation: @HIDEBUTTON |

| | | | |
|---|---|---|---|
| 340 | covid_diagnosis_pasc_y_adj<br><br>Show the field ONLY if:<br>[conditions_post(1)] = '1' and [covid_diagnosis_pasc] = '1' | Copy and paste the language from the physician notes that says this continued COVID diagnosis was related to PASC (make sure to remove any PHI, such as MRNs or names): [covid_diagnosis_pasc_y] | descriptive, Required<br>Field Annotation: @HIDEBUTTON |
| 341 | acuteres_date_followup_adj<br><br>Show the field ONLY if:<br>[conditions_post(2)] = '1' | Enter the first and second (when applicable) Acute respiratory distress syndrome diagnosis dates found during the follow-up period:[acuteres_date_followup] [acuteres_date_followup_2] | descriptive, Required<br>Field Annotation: @HIDEBUTTON |
| 342 | acuteres_endstatus_adj<br><br>Show the field ONLY if:<br>[conditions_post(2)] = '1' | At the end of the follow-up period, the Acute respiratory distress syndrome was: [acuteres_endstatus] | descriptive, Required<br>Field Annotation: @HIDEBUTTON |
| 343 | acuteres_diagnosis_pasc_adj<br><br>Show the field ONLY if:<br>[conditions_post(2)] = '1' | Did the physician state in the chart whether the Acute respiratory distress syndrome diagnosis was due to PASC?[acuteres_diagnosis_pasc] | descriptive, Required<br>Field Annotation: @HIDEBUTTON |
| 344 | acuteres_diagnosis_pasc_y_adj<br><br>Show the field ONLY if:<br>[conditions_post(2)] = '1' and [acuteres_diagnosis_pasc] = '1' | Copy and paste the language from the physician notes that states this Acute respiratory distress syndrome diagnosis was related to PASC: [acuteres_diagnosis_pasc_y] | descriptive, Required<br>Field Annotation: @HIDEBUTTON |
| 345 | losssmell_date_followup_adj<br><br>Show the field ONLY if:<br>[conditions_post(3)] = '1' | Enter the first and second (when applicable) Loss of smell diagnosis dates found during the follow-up period:[losssmell_date_followup] [losssmell_date_followup_2] | descriptive, Required<br>Field Annotation: @HIDEBUTTON |
| 346 | losssmell_endstatus_adj<br><br>Show the field ONLY if:<br>[conditions_post(3)] = '1' | At the end of the follow-up period, the Loss of smell was:[losssmell_endstatus] | descriptive, Required<br>Field Annotation: @HIDEBUTTON |
| 347 | losssmell_diagnosis_pasc_adj<br><br>Show the field ONLY if:<br>[conditions_post(3)] = '1' | Did the physician state in the chart whether the Loss of smell diagnosis was due to PASC?[losssmell_diagnosis_pasc] | descriptive, Required<br>Field Annotation: @HIDEBUTTON |
| 348 | losssmell_diagnosis_pasc_y_adj<br><br>Show the field ONLY if:<br>[conditions_post(3)] = '1' and [losssmell_diagnosis_pasc] = '1' | Copy and paste the language from the physician notes that states the Loss of smell diagnosis was related to PASC:[losssmell_diagnosis_pasc_y] | descriptive, Required<br>Field Annotation: @HIDEBUTTON |
| 349 | losstaste_date_followup_adj<br><br>Show the field ONLY if: | Enter the first and second (when applicable) Loss of taste diagnosis dates found during the follow-up period:[losstaste_date_followup] [losstaste_date_followup_2] | descriptive, Required<br>Field Annotation: @HIDEBUTTON |

| | | | |
|---|---|---|---|
| | [conditions_post(4)] = '1' | | |
| 350 | losstaste_endstatus_adj<br><br>Show the field ONLY if:<br>[conditions_post(4)] = '1' | At the end of the follow-up period, the Loss of taste was:[losstaste_endstatus] | descriptive, Required<br>Field Annotation: @HIDEBUTTON |
| 351 | losstaste_diagnosis_pasc_adj<br><br>Show the field ONLY if:<br>[conditions_post(4)] = '1' | Did the physician state in the chart whether the Loss of taste diagnosis was due to PASC?<br>[losstaste_diagnosis_pasc] | descriptive, Required<br>Field Annotation: @HIDEBUTTON |
| 352 | losstaste_diagnosis_pasc_y_adj<br><br>Show the field ONLY if:<br>[conditions_post(4)] = '1' and [losstaste_diagnosis_pasc] = '1' | Copy and paste the language from the physician notes that states the Loss of taste diagnosis was due to PASC:[losstaste_diagnosis_pasc_y] | descriptive, Required<br>Field Annotation: @HIDEBUTTON |
| 353 | lossother_date_followup_adj<br><br>Show the field ONLY if:<br>[conditions_post(5)] = '1' | Enter the first and second (when applicable) Other Change of Smell/Taste date found during the follow-up period:[lossother_date_followup]<br>[lossother_date_followup_2] | descriptive, Required<br>Field Annotation: @HIDEBUTTON |
| 354 | lossother_endstatus_adj<br><br>Show the field ONLY if:<br>[conditions_post(5)] = '1' | At the end of the follow-up period, the Other Change of Smell/Taste was:[lossother_endstatus] | descriptive, Required<br>Field Annotation: @HIDEBUTTON |
| 355 | lossother_diagnosis_pasc_adj<br><br>Show the field ONLY if:<br>[conditions_post(5)] = '1' | Did the physician state in the chart whether the Other Change of Smell/Taste was due to PASC?<br>[lossother_diagnosis_pasc] | descriptive, Required<br>Field Annotation: @HIDEBUTTON |
| 356 | lossother_diagnosis_pasc_y_adj<br><br>Show the field ONLY if:<br>[conditions_post(5)] = '1' and [lossother_diagnosis_pasc] = '1' | Copy and paste the language from the physician notes that states the Other Change of Smell/Taste was due to PASC:[lossother_diagnosis_pasc_y] | descriptive, Required<br>Field Annotation: @HIDEBUTTON |
| 357 | myocarditis_date_followup_adj<br><br>Show the field ONLY if:<br>[conditions_post(6)] = '1' | Enter the first and second (when applicable) Myocarditis diagnosis date found during the follow-up period:[myocarditis_date_followup]<br>[myocarditis_date_followup_2] | descriptive, Required<br>Field Annotation: @HIDEBUTTON |
| 358 | myocarditis_endstatus_adj<br><br>Show the field ONLY if:<br>[conditions_post(6)] = '1' | At the end of the follow-up period, the Myocarditis was:[myocarditis_endstatus] | descriptive, Required<br>Field Annotation: @HIDEBUTTON |
| 359 | myocarditis_diagnosis_pasc_adj<br><br>Show the field ONLY if:<br>[conditions_post(6)] = '1' | Did the physician state in the notes whether the Myocarditis diagnosis was due to PASC?<br>[myocarditis_diagnosis_pasc] | descriptive, Required<br>Field Annotation: @HIDEBUTTON |

| | | | |
|---|---|---|---|
| 360 | myocarditis_diagnosis_pasc_y_adj<br><br>Show the field ONLY if:<br>[conditions_post(6)] = '1' and [myocarditis_diagnosis_pasc] = '1' | Copy and paste the language from the physician notes that states the Myocarditis diagnosis was due to PASC:[myocarditis_diagnosis_pasc_y] | descriptive, Required<br>Field Annotation: @HIDEBUTTON |
| 361 | pericarditis_date_followup_adj<br><br>Show the field ONLY if:<br>[conditions_post(7)] = '1' | Enter the first and second (when applicable) Pericarditis diagnosis date found during the follow-up period:[pericarditis_date_followup] [pericarditis_date_followup_2] | descriptive, Required<br>Field Annotation: @HIDEBUTTON |
| 362 | pericarditis_endstatus_adj<br><br>Show the field ONLY if:<br>[conditions_post(7)] = '1' | At the end of the follow-up period, the Pericarditis was:[pericarditis_endstatus] | descriptive, Required<br>Field Annotation: @HIDEBUTTON |
| 363 | pericarditis_diagnosis_pasc_adj<br><br>Show the field ONLY if:<br>[conditions_post(7)] = '1' | Did the physician state in the notes whether the Pericarditis diagnosis was due to PASC?[pericarditis_diagnosis_pasc] | descriptive, Required<br>Field Annotation: @HIDEBUTTON |
| 364 | pericarditis_diagnosis_pasc_y_adj<br><br>Show the field ONLY if:<br>[conditions_post(7)] = '1' and [pericarditis_diagnosis_pasc] = '1' | Copy and paste the language from the physician notes that states the Pericarditis diagnosis was due to PASC:[pericarditis_diagnosis_pasc_y] | descriptive, Required<br>Field Annotation: @HIDEBUTTON |
| 365 | myositis_date_followup_adj<br><br>Show the field ONLY if:<br>[conditions_post(8)] = '1' | Enter the first and second (when applicable) Myositis diagnosis date found during the follow-up period:[myositis_date_followup] [myositis_date_followup_2] | descriptive, Required<br>Field Annotation: @HIDEBUTTON |
| 366 | myositis_endstatus_adj<br><br>Show the field ONLY if:<br>[conditions_post(8)] = '1' | At the end of the follow-up period, the Myositis was:[myositis_endstatus] | descriptive, Required<br>Field Annotation: @HIDEBUTTON |
| 367 | myositis_diagnosis_pasc_adj<br><br>Show the field ONLY if:<br>[conditions_post(8)] = '1' | Did the physician state in the notes whether the Myositis diagnosis was due to PASC?[myositis_diagnosis_pasc] | descriptive, Required<br>Field Annotation: @HIDEBUTTON |
| 368 | myositis_diagnosis_pasc_y_adj<br><br>Show the field ONLY if:<br>[conditions_post(8)] = '1' and [myositis_diagnosis_pasc] = '1' | Copy and paste the language from the notes that states the Myositis diagnosis was due to PASC:[myositis_diagnosis_pasc_y] | descriptive, Required<br>Field Annotation: @HIDEBUTTON |
| 369 | illheart_date_followup_adj | Enter the first and second (when applicable) Other/ill-defined heart disease diagnosis date found during the follow-up period:[illheart_date_followup] | descriptive, Required<br>Field Annotation: @HIDEBUTTON |

| | | | |
|---|---|---|---|
| | Show the field ONLY if: [conditions_post(9)] = '1' | [illheart_date_followup_2] | |
| 370 | illheart_endstatus_adj<br>Show the field ONLY if: [conditions_post(9)] = '1' | At the end of the follow-up period, the Other/ill-defined heart disease was:[illheart_endstatus] | descriptive, Required<br>Field Annotation: @HIDEBUTTON |
| 371 | illheart_diagnosis_pasc_adj<br>Show the field ONLY if: [conditions_post(9)] = '1' | Did the physician state in the notes whether the Other/ill-defined heart disease diagnosis was due to PASC?[illheart_diagnosis_pasc] | descriptive, Required<br>Field Annotation: @HIDEBUTTON |
| 372 | illheart_diagnosis_pasc_y_adj<br>Show the field ONLY if: [conditions_post(9)] = '1' and [illheart_diagnosis_pasc] = '1' | Copy and paste the language from the notes that states the Other/ill-defined heart disease diagnosis was due to PASC:[illheart_diagnosis_pasc_y] | descriptive, Required<br>Field Annotation: @HIDEBUTTON |
| 373 | thrombo_date_followup_adj<br>Show the field ONLY if: [conditions_post(10)] = '1' | Enter the first and second (when applicable) Thrombophlebitis/thromboembolism diagnosis date found during the follow-up period:[thrombo_date_followup] [thrombo_date_followup_2] | descriptive, Required<br>Field Annotation: @HIDEBUTTON |
| 374 | thrombo_endstatus_adj<br>Show the field ONLY if: [conditions_post(10)] = '1' | At the end of the follow-up period, the Thrombophlebitis/thromboembolism was:[thrombo_endstatus] | descriptive, Required<br>Field Annotation: @HIDEBUTTON |
| 375 | thrombo_diagnosis_pasc_adj<br>Show the field ONLY if: [conditions_post(10)] = '1' | Did the physician state in the notes whether the Thrombophlebitis/thromboembolism diagnosis was due to PASC?[thrombo_diagnosis_pasc] | descriptive, Required<br>Field Annotation: @HIDEBUTTON |
| 376 | thrombo_diagnosis_pasc_y_adj<br>Show the field ONLY if: [conditions_post(10)] = '1' and [thrombo_diagnosis_pasc] = '1' | Copy and paste the language from the notes that states the Thrombophlebitis/thromboembolism diagnosis was due to PASC:[thrombo_diagnosis_pasc_y] | descriptive, Required<br>Field Annotation: @HIDEBUTTON |
| 377 | anemia_date_followup_adj<br>Show the field ONLY if: [conditions_post(11)] = '1' | Enter the first and second (when applicable) Aplastic anemia diagnosis date found during the follow-up period:[anemia_date_followup] [anemia_date_followup_2] | descriptive, Required<br>Field Annotation: @HIDEBUTTON |
| 378 | anemia_endstatus_adj<br>Show the field ONLY if: [conditions_post(11)] = ' | At the end of the follow-up period, the Aplastic anemia was:[anemia_endstatus] | descriptive, Required<br>Field Annotation: @HIDEBUTTON |

| | | | |
|---|---|---|---|
| | 1' | | |
| 379 | anemia_diagnosis_pasc_adj<br><br>Show the field ONLY if:<br>[conditions_post(11)] = '1' | Did the physician state in the notes whether the Aplastic anemia diagnosis was due to PASC?[anemia_diagnosis_pasc] | descriptive, Required<br>Field Annotation: @HIDEBUTTON |
| 380 | anemia_diagnosis_pasc_y_adj<br><br>Show the field ONLY if:<br>[conditions_post(11)] = '1' and [anemia_diagnosis_pasc] = '1' | Copy and paste the language from the notes that states the Aplastic anemia diagnosis was due to PASC:[anemia_diagnosis_pasc_y] | descriptive, Required<br>Field Annotation: @HIDEBUTTON |
| 381 | brainfog_date_followup_adj<br><br>Show the field ONLY if:<br>[conditions_post(12)] = '1' | Enter the first and second (when applicable) Brain fog diagnosis date found during the follow-up period:[brainfog_date_followup] [brainfog_date_followup_2] | descriptive, Required<br>Field Annotation: @HIDEBUTTON |
| 382 | brainfog_endstatus_adj<br><br>Show the field ONLY if:<br>[conditions_post(12)] = '1' | At the end of the follow-up period, the Brain fog was:[brainfog_endstatus] | descriptive, Required<br>Field Annotation: @HIDEBUTTON |
| 383 | brainfog_diagnosis_pasc_adj<br><br>Show the field ONLY if:<br>[conditions_post(12)] = '1' | Did the physician state in the notes whether the Brain fog diagnosis was due to PASC?[brainfog_diagnosis_pasc] | descriptive, Required<br>Field Annotation: @HIDEBUTTON |
| 384 | brainfog_diagnosis_pasc_y_adj<br><br>Show the field ONLY if:<br>[conditions_post(12)] = '1' and [brainfog_diagnosis_pasc] = '1' | Copy and paste the language from the notes that states the Brain fog diagnosis was due to PASC:[brainfog_diagnosis_pasc_y] | descriptive, Required<br>Field Annotation: @HIDEBUTTON |
| 385 | liverenzy_date_followup_adj<br><br>Show the field ONLY if:<br>[conditions_post(13)] = '1' | Enter the first and second (when applicable) Abnormal liver enzymes diagnosis date found during the follow-up period:[liverenzy_date_followup] [liverenzy_date_followup_2] | descriptive, Required<br>Field Annotation: @HIDEBUTTON |
| 386 | liverenzy_endstatus_adj<br><br>Show the field ONLY if:<br>[conditions_post(13)] = '1' | At the end of the follow-up period, the Abnormal liver enzymes were:[liverenzy_endstatus] | descriptive, Required<br>Field Annotation: @HIDEBUTTON |
| 387 | liverenzy_diagnosis_pasc_adj | Did the physician state in the notes whether the Abnormal liver enzymes diagnosis was due to PASC?[liverenzy_diagnosis_pasc] | descriptive, Required<br>Field Annotation: @HIDEBUTTON |

| | | | |
|---|---|---|---|
| | Show the field ONLY if: [conditions_post(13)] = '1' | | |
| 388 | liverenzy_diagnosis_pasc_y_adj<br>Show the field ONLY if: [conditions_post(13)] = '1' and [liverenzy_diagnosis_pasc] = '1' | Copy and paste the language from the notes that states the Abnormal liver enzymes diagnosis was due to PASC:[liverenzy_diagnosis_pasc_y] | descriptive, Required<br>Field Annotation: @HIDEBUTTON |
| 389 | dysautonomia_date_followup_adj<br>Show the field ONLY if: [conditions_post(14)] = '1' | Enter the first and second (when applicable) Dysautonomia diagnosis date found during the follow-up period:[dysautonomia_date_followup] [dysautonomia_date_followup_2] | descriptive, Required<br>Field Annotation: @HIDEBUTTON |
| 390 | dysautonomia_endstatus_adj<br>Show the field ONLY if: [conditions_post(14)] = '1' | At the end of the follow-up period, the Dysautonomia was:[dysautonomia_endstatus] | descriptive, Required<br>Field Annotation: @HIDEBUTTON |
| 391 | dysautonomia_diagnosis_pasc_adj<br>Show the field ONLY if: [conditions_post(14)] = '1' | Did the physician state in the notes whether the Dysautonomia diagnosis was due to PASC? [dysautonomia_diagnosis_pasc] | descriptive, Required<br>Field Annotation: @HIDEBUTTON |
| 392 | dysautonomia_diagnosis_pasc_y_adj<br>Show the field ONLY if: [conditions_post(14)] = '1' and [dysautonomia_diagnosis_pasc] = '1' | Copy and paste the language from the notes that states the Dysautonomia diagnosis was due to PASC: [dysautonomia_diagnosis_pasc_y] | descriptive, Required<br>Field Annotation: @HIDEBUTTON |
| 393 | fatigue_date_followup_adj<br>Show the field ONLY if: [conditions_post(15)] = '1' | Enter the first and second (when applicable) Fatigue diagnosis date found during the follow-up period: [fatigue_date_followup] [fatigue_date_followup_2] | descriptive, Required<br>Field Annotation: @HIDEBUTTON |
| 394 | fatigue_endstatus_adj<br>Show the field ONLY if: [conditions_post(15)] = '1' | At the end of the follow-up period, the Fatigue was: [fatigue_endstatus] | descriptive, Required<br>Field Annotation: @HIDEBUTTON |
| 395 | fatigue_diagnosis_pasc_adj<br>Show the field ONLY if: [conditions_post(15)] = '1' | Did the physician state in the notes whether the Fatigue diagnosis was due to PASC? [fatigue_diagnosis_pasc] | descriptive, Required<br>Field Annotation: @HIDEBUTTON |
| 396 | fatigue_diagnosis_p | Copy and paste the language from the notes that | descriptive, Required |

| | | | |
|---|---|---|---|
| | asc_y_adj<br><br>Show the field ONLY if:<br>[conditions_post(15)] = '1' and [fatigue_diagnosis_pasc] = '1' | states the Fatigue diagnosis was due to PASC:[fatigue_diagnosis_pasc_y] | Field Annotation: @HIDEBUTTON |
| 397 | abdominal_date_followup_adj<br><br>Show the field ONLY if:<br>[conditions_post(19)] = '1' | Enter the first and second (when applicable) Abdominal Pain date found during the follow-up period:[abdominal_date_followup] [abdominal_date_followup_2] | descriptive, Required<br>Field Annotation: @HIDEBUTTON |
| 398 | abdominal_endstatus_adj<br><br>Show the field ONLY if:<br>[conditions_post(19)] = '1' | At the end of the follow-up period, the Abdominal Pain was:[abdominal_endstatus] | descriptive, Required<br>Field Annotation: @HIDEBUTTON |
| 399 | abdominal_diagnosis_pasc_adj<br><br>Show the field ONLY if:<br>[conditions_post(19)] = '1' | Did the physician state in the notes whether the Abdominal Pain diagnosis was due to PASC?[abdominal_diagnosis_pasc] | descriptive, Required<br>Field Annotation: @HIDEBUTTON |
| 400 | abdominal_diagnosis_pasc_y_adj<br><br>Show the field ONLY if:<br>[conditions_post(19)] = '1' and [abdominal_diagnosis_pasc] = '1' | Copy and paste the language from the notes that states the Abdominal Pain diagnosis was due to PASC:[abdominal_diagnosis_pasc_y] | descriptive, Required<br>Field Annotation: @HIDEBUTTON |
| 401 | aki_date_followup_adj<br><br>Show the field ONLY if:<br>[conditions_post(20)] = '1' | Enter the first and second (when applicable) Acute Kidney Injury diagnosis date found during the follow-up period:[aki_date_followup] [aki_date_followup_2] | descriptive, Required<br>Field Annotation: @HIDEBUTTON |
| 402 | aki_endstatus_adj<br><br>Show the field ONLY if:<br>[conditions_post(20)] = '1' | At the end of the follow-up period, the Acute Kidney Injury was:[aki_endstatus] | descriptive, Required<br>Field Annotation: @HIDEBUTTON |
| 403 | aki_diagnosis_pasc_adj<br><br>Show the field ONLY if:<br>[conditions_post(20)] = '1' | Did the physician state in the notes whether the Acute Kidney Injury diagnosis was due to PASC?[aki_diagnosis_pasc] | descriptive, Required<br>Field Annotation: @HIDEBUTTON |
| 404 | aki_diagnosis_pasc_y_adj<br><br>Show the field ONLY if:<br>[conditions_post(20)] = '1' and [aki_diagnosis_pasc] = '1' | Copy and paste the language from the notes that states the Acute Kidney Injury diagnosis was due to PASC:[aki_diagnosis_pasc_y] | descriptive, Required<br>Field Annotation: @HIDEBUTTON |

| | | | |
|---|---|---|---|
| 405 | abnormal_lab_adj<br>Show the field ONLY if:<br>[covid] = '1' | Did the patient have at least one occurrence of the following abnormal lab tests, on Day 28 or later after initial COVID-19 positive diagnosis? Select all that apply. [abnormal_lab] | descriptive |
| 406 | reason_thrombo_test_adj<br>Show the field ONLY if:<br>[covid] = '1' and [abnormal_lab(1)] = '1' | Select the reason why the patient was tested for Thrombocytopenia:[reason_thrombo_test] | descriptive, Required |
| 407 | date_thrombo_test_adj<br>Show the field ONLY if:<br>[covid] = '1' and [abnormal_lab(1)] = '1' | Enter the Thrombocytopenia test date:[date_thrombo_test] | descriptive, Required<br>Field Annotation: @HIDEBUTTON |
| 408 | reason_troponin_test_adj<br>Show the field ONLY if:<br>[covid] = '1' and [abnormal_lab(2)] = '1' | Select the reason why the patient was tested for Elevated Troponin:[reason_troponin_test] | descriptive, Required |
| 409 | date_troponin_test_adj<br>Show the field ONLY if:<br>[covid] = '1' and [abnormal_lab(2)] = '1' | Enter the Elevated Troponin test date:[date_troponin_test] | descriptive, Required<br>Field Annotation: @HIDEBUTTON |
| 410 | reason_lymphopenia_test_adj<br>Show the field ONLY if:<br>[covid] = '1' and [abnormal_lab(3)] = '1' | Select the reason why the patient was tested for Lymphopenia:[reason_lymphopenia_test] | descriptive, Required |
| 411 | date_lymphopenia_test_adj<br>Show the field ONLY if:<br>[covid] = '1' and [abnormal_lab(3)] = '1' | Enter the Lymphopenia test date:[date_lymphopenia_test] | descriptive, Required<br>Field Annotation: @HIDEBUTTON |
| 412 | reason_crp_test_adj<br>Show the field ONLY if:<br>[covid] = '1' and [abnormal_lab(4)] = '1' | Select the reason why the patient was tested for Elevated CRP:[reason_crp_test] | descriptive, Required |
| 413 | date_crp_test_adj<br>Show the field ONLY if:<br>[covid] = '1' and [abnormal_lab(4)] = '1' | Enter the Elevated CRP test date:[date_crp_test] | descriptive, Required<br>Field Annotation: @HIDEBUTTON |
| 414 | diagnosis_post_covid_adj | Did the patient have at least one occurrences of a diagnosis term for the following on Day 28 or later after COVID positive diagnosis? Select all that apply. | descriptive |

| | | | |
|---|---|---|---|
| | Show the field ONLY if: [covid] = '1' | [diagnosis_post_covid] | |
| 415 | chestpain_date_adj<br>Show the field ONLY if: [covid] = '1' and [diagnosis_post_covid(1)] = '1' | Enter the date of the first and second Chest Pain diagnosis:[chestpain_date] [chestpain_date_2] | descriptive, Required<br>Field Annotation: @HIDEBUTTON |
| 416 | chestpain_endstatus_adj<br>Show the field ONLY if: [covid] = '1' and [diagnosis_post_covid(1)] = '1' | At the end of the follow-up period, the Chest Pain was:[chestpain_endstatus] | descriptive, Required<br>Field Annotation: @HIDEBUTTON |
| 417 | chestpain_pasc_adj<br>Show the field ONLY if: [covid] = '1' and [diagnosis_post_covid(1)] = '1' | Did the physician state in the notes whether the Chest Pain was related to PASC?[chestpain_pasc] | descriptive, Required<br>Field Annotation: @HIDEBUTTON |
| 418 | chestpain_pasc_y_adj<br>Show the field ONLY if: [covid] = '1' and [diagnosis_post_covid(1)] = '1' and [chestpain_pasc] = '1' | Copy and paste the language from the notes that states the Chest Pain diagnosis was related to PASC:[chestpain_pasc_y] | descriptive, Required<br>Field Annotation: @HIDEBUTTON |
| 419 | hairloss_date_adj<br>Show the field ONLY if: [covid] = '1' and [diagnosis_post_covid(2)] = '1' | Enter the date of the first and second Hair Loss diagnosis:[hairloss_date] [hairloss_date_2] | descriptive, Required<br>Field Annotation: @HIDEBUTTON |
| 420 | hairloss_endstatus_adj<br>Show the field ONLY if: [covid] = '1' and [diagnosis_post_covid(2)] = '1' | At the end of the follow-up period, the Hair Loss was:[hairloss_endstatus] | descriptive, Required<br>Field Annotation: @HIDEBUTTON |
| 421 | hairloss_pasc_adj<br>Show the field ONLY if: [covid] = '1' and [diagnosis_post_covid(2)] = '1' | Did the physician state in the notes whether the Hair Loss was related to PASC?[hairloss_pasc] | descriptive, Required<br>Field Annotation: @HIDEBUTTON |
| 422 | hairloss_pasc_y_adj<br>Show the field ONLY if: [covid] = '1' and [diagnosis_post_covid(2)] = '1' and [hairloss_pasc] = '1' | Copy and paste the language from the notes that states the Hair Loss diagnosis was related to PASC:[hairloss_pasc_y] | descriptive, Required<br>Field Annotation: @HIDEBUTTON |
| 423 | cough_date_adj<br>Show the field ONLY if: [covid] = '1' and [diagnosis_post_covid(3)] = '1' | Enter the date of the first and second Cough diagnosis:[cough_date] [cough_date_2] | descriptive, Required<br>Field Annotation: @HIDEBUTTON |
| 424 | cough_endstatus_ad | At the end of the follow-up period, the Cough was: | descriptive, Required |

| | | | |
|---|---|---|---|
| | j<br><br>Show the field ONLY if:<br>[covid] = '1' and [diagnos is_post_covid(3)] = '1' | [cough_endstatus] | Field Annotation: @HIDEBUTTON |
| 425 | cough_pasc_adj<br><br>Show the field ONLY if:<br>[covid] = '1' and [diagnos is_post_covid(3)] = '1' | Did the physician state in the notes whether the Cough was related to PASC?[cough_pasc] | descriptive, Required<br>Field Annotation: @HIDEBUTTON |
| 426 | cough_pasc_y_adj<br><br>Show the field ONLY if:<br>[covid] = '1' and [diagnos is_post_covid(3)] = '1' an d [cough_pasc] = '1' | Copy and paste the language from the notes that states the Cough diagnosis was related to PASC:[cough_pasc_y] | descriptive, Required<br>Field Annotation: @HIDEBUTTON |
| 427 | cardio_signs_date_a dj<br><br>Show the field ONLY if:<br>[covid] = '1' and [diagnos is_post_covid(4)] = '1' | Enter the date of the first and second Cardiorespiratory signs and symptoms:[cardio_signs_date] [cardio_signs_date_2] | descriptive, Required<br>Field Annotation: @HIDEBUTTON |
| 428 | cardioresp_endstatu s_adj<br><br>Show the field ONLY if:<br>[covid] = '1' and [diagnos is_post_covid(4)] = '1' | At the end of the follow-up period, the Cardiorespiratory signs and symptoms were:[cardioresp_endstatus] | descriptive, Required<br>Field Annotation: @HIDEBUTTON |
| 429 | cardioresp_pasc_ad j<br><br>Show the field ONLY if:<br>[covid] = '1' and [diagnos is_post_covid(4)] = '1' | Did the physician state in the notes whether the Cardiorespiratory signs and symptoms were related to PASC?[cardioresp_pasc] | descriptive, Required<br>Field Annotation: @HIDEBUTTON |
| 430 | cardioresp_pasc_y_a dj<br><br>Show the field ONLY if:<br>[covid] = '1' and [diagnos is_post_covid(4)] = '1' an d [cardioresp_pasc] = '1' | Copy and paste the language from the notes that states the Cardiorespiratory signs and symptoms were related to PASC:[cardioresp_pasc_y] | descriptive, Required<br>Field Annotation: @HIDEBUTTON |
| 431 | jaundice_date_adj<br><br>Show the field ONLY if:<br>[covid] = '1' and [diagnos is_post_covid(5)] = '1' | Enter the date of the first and second Jaundice diagnosis:[jaundice_date] [jaundice_date_2] | descriptive, Required<br>Field Annotation: @HIDEBUTTON |
| 432 | jaundice_endstatus_ adj<br><br>Show the field ONLY if:<br>[covid] = '1' and [diagnos is_post_covid(5)] = '1' | At the end of the follow-up period, the Jaundice was:[jaundice_endstatus] | descriptive, Required<br>Field Annotation: @HIDEBUTTON |
| 433 | jaundice_pasc_adj<br><br>Show the field ONLY if: | Did the physician state in the notes whether the Jaundice was related to PASC?[jaundice_pasc] | descriptive, Required<br>Field Annotation: @HIDEBUTTON |

| | | | |
|---|---|---|---|
| | [covid] = '1' and [diagnosis_post_covid(5)] = '1' | | |
| 434 | jaundice_pasc_y_adj<br><br>Show the field ONLY if:<br>[covid] = '1' and [diagnosis_post_covid(5)] = '1' and [jaundice_pasc] = '1' | Copy and paste the language from the notes that states the Jaundice was related to PASC:[jaundice_pasc_y] | descriptive, Required<br>Field Annotation: @HIDEBUTTON |
| 435 | pain_date_adj<br><br>Show the field ONLY if:<br>[covid] = '1' and [diagnosis_post_covid(6)] = '1' | Enter the date of the first and second Generalized Pain diagnosis:[pain_date] [pain_date_2] | descriptive, Required<br>Field Annotation: @HIDEBUTTON |
| 436 | pain_endstatus_adj<br><br>Show the field ONLY if:<br>[covid] = '1' and [diagnosis_post_covid(6)] = '1' | At the end of the follow-up period, the Generalized Pain was:[pain_endstatus] | descriptive, Required<br>Field Annotation: @HIDEBUTTON |
| 437 | pain_pasc_adj<br><br>Show the field ONLY if:<br>[covid] = '1' and [diagnosis_post_covid(6)] = '1' | Did the physician state in the notes whether the Generalized Pain was related to PASC?[pain_pasc] | descriptive, Required<br>Field Annotation: @HIDEBUTTON |
| 438 | pain_pasc_y_adj<br><br>Show the field ONLY if:<br>[covid] = '1' and [diagnosis_post_covid(6)] = '1' and [pain_pasc] = '1' | Copy and paste the language from the notes that states the Generalized Pain was related to PASC:[pain_pasc_y] | descriptive, Required<br>Field Annotation: @HIDEBUTTON |
| 439 | anxiety_date_adj<br><br>Show the field ONLY if:<br>[covid] = '1' and [diagnosis_post_covid(7)] = '1' | Enter the date of the first and second Anxiety symptoms:[anxiety_date] [anxiety_date_2] | descriptive, Required<br>Field Annotation: @HIDEBUTTON |
| 440 | anxiety_endstatus_adj<br><br>Show the field ONLY if:<br>[covid] = '1' and [diagnosis_post_covid(7)] = '1' | At the end of the follow-up period, the Anxiety Symptoms were:[anxiety_endstatus] | descriptive, Required<br>Field Annotation: @HIDEBUTTON |
| 441 | anxiety_pasc_adj<br><br>Show the field ONLY if:<br>[covid] = '1' and [diagnosis_post_covid(7)] = '1' | Did the physician state in the notes whether the Anxiety Symptoms were related to PASC?[anxiety_pasc] | descriptive, Required<br>Field Annotation: @HIDEBUTTON |
| 442 | anxiety_pasc_y_adj<br><br>Show the field ONLY if:<br>[covid] = '1' and [diagnosis_post_covid(7)] = '1' and [anxiety_pasc] = '1' | Copy and paste the language from the notes that states the Anxiety symptoms were related to PASC:[anxiety_pasc_y] | descriptive, Required<br>Field Annotation: @HIDEBUTTON |
| 443 | fatigue_malaise_dat | Enter the date of the first and second | descriptive, Required |

| | | | |
|---|---|---|---|
| | e_adj<br><br>Show the field ONLY if:<br>[covid] = '1' and [diagnosis_post_covid(8)] = '1' | Fatigue/Malaise diagnosis:[fatigue_malaise_date] [fatigue_malaise_date_2] | Field Annotation: @HIDEBUTTON |
| 444 | fatigue_malaise_endstatus_adj<br><br>Show the field ONLY if:<br>[covid] = '1' and [diagnosis_post_covid(8)] = '1' | At the end of the follow-up period, the Fatigue/Malaise Symptoms were:[fatigue_malaise_endstatus] | descriptive, Required<br>Field Annotation: @HIDEBUTTON |
| 445 | fatigue_malaise_pasc_adj<br><br>Show the field ONLY if:<br>[covid] = '1' and [diagnosis_post_covid(8)] = '1' | Did the physician state in the notes whether the Fatigue/Malaise Symptoms were related to PASC?[fatigue_malaise_pasc] | descriptive, Required<br>Field Annotation: @HIDEBUTTON |
| 446 | fatigue_malaise_pasc_y_adj<br><br>Show the field ONLY if:<br>[covid] = '1' and [diagnosis_post_covid(8)] = '1' and [fatigue_malaise_pasc] = '1' | Copy and paste the language from the notes that states the Fatigue/Malaise symptoms were related to PASC:[fatigue_malaise_pasc_y] | descriptive, Required<br>Field Annotation: @HIDEBUTTON |
| 447 | diarrhea_date_adj<br><br>Show the field ONLY if:<br>[covid] = '1' and [diagnosis_post_covid(9)] = '1' | Enter the date of the first and second Diarrhea diagnosis:[diarrhea_date] [diarrhea_date_2] | descriptive, Required<br>Field Annotation: @HIDEBUTTON |
| 448 | diarrhea_endstatus_adj<br><br>Show the field ONLY if:<br>[covid] = '1' and [diagnosis_post_covid(9)] = '1' | At the end of the follow-up period, the Diarrhea was:[diarrhea_endstatus] | descriptive, Required<br>Field Annotation: @HIDEBUTTON |
| 449 | diarrhea_pasc_adj<br><br>Show the field ONLY if:<br>[covid] = '1' and [diagnosis_post_covid(9)] = '1' | Did the physician state in the notes whether the Diarrhea was related to PASC?[diarrhea_pasc] | descriptive, Required<br>Field Annotation: @HIDEBUTTON |
| 450 | diarrhea_pasc_y_adj<br><br>Show the field ONLY if:<br>[covid] = '1' and [diagnosis_post_covid(9)] = '1' and [diarrhea_pasc] = '1' | Copy and paste the language from the notes that states the Diarrhea was related to PASC:[diarrhea_pasc_y] | descriptive, Required<br>Field Annotation: @HIDEBUTTON |
| 451 | fever_chills_date_adj<br><br>Show the field ONLY if:<br>[covid] = '1' and [diagnosis_post_covid(10)] = '1' | Enter the date of the first and second Fever/Chills diagnosis:[fever_chills_date] [fever_chills_date_2] | descriptive, Required<br>Field Annotation: @HIDEBUTTON |
| 452 | fever_chills_endsta | At the end of the follow-up period, the Fever/Chills | descriptive, Required |

| | | | |
|---|---|---|---|
| | tus_adj<br><br>Show the field ONLY if:<br>[covid] = '1' and [diagnosis_post_covid(10)] = '1' | were:[fever_chills_endstatus] | Field Annotation: @HIDEBUTTON |
| 453 | fever_chills_pasc_adj<br><br>Show the field ONLY if:<br>[covid] = '1' and [diagnosis_post_covid(10)] = '1' | Did the physician state in the notes whether the Fever/Chills were related to PASC?[fever_chills_pasc] | descriptive, Required<br>Field Annotation: @HIDEBUTTON |
| 454 | fever_chills_pasc_y_adj<br><br>Show the field ONLY if:<br>[covid] = '1' and [diagnosis_post_covid(10)] = '1' and [fever_chills_pasc] = '1' | Copy and paste the language from the notes that states the Fever/Chills were related to PASC:[fever_chills_pasc_y] | descriptive, Required<br>Field Annotation: @HIDEBUTTON |
| 455 | skin_rash_date_adj<br><br>Show the field ONLY if:<br>[covid] = '1' and [diagnosis_post_covid(11)] = '1' | Enter the date of the first and second Skin Rash diagnosis:[skin_rash_date] [skin_rash_date_2] | descriptive, Required<br>Field Annotation: @HIDEBUTTON |
| 456 | skinrash_endstatus_adj<br><br>Show the field ONLY if:<br>[covid] = '1' and [diagnosis_post_covid(11)] = '1' | At the end of the follow-up period, the Skin Rash was:[skinrash_endstatus] | descriptive, Required<br>Field Annotation: @HIDEBUTTON |
| 457 | skin_rash_pasc_adj<br><br>Show the field ONLY if:<br>[covid] = '1' and [diagnosis_post_covid(11)] = '1' | Did the physician state in the notes whether the Skin Rash was related to PASC?[skin_rash_pasc] | descriptive, Required<br>Field Annotation: @HIDEBUTTON |
| 458 | skin_rash_pasc_y_adj<br><br>Show the field ONLY if:<br>[covid] = '1' and [diagnosis_post_covid(11)] = '1' and [skin_rash_pasc] = '1' | Copy and paste the language from the notes that states the Skin Rash was related to PASC:[skin_rash_pasc_y] | descriptive, Required<br>Field Annotation: @HIDEBUTTON |
| 459 | headache_date_adj<br><br>Show the field ONLY if:<br>[covid] = '1' and [diagnosis_post_covid(12)] = '1' | Enter the date of the first and second Headache diagnosis:[headache_date] [headache_date_2] | descriptive, Required<br>Field Annotation: @HIDEBUTTON |
| 460 | headache_endstatus_adj<br><br>Show the field ONLY if:<br>[covid] = '1' and [diagnosis_post_covid(12)] = '1' | At the end of the follow-up period, the Headache was:[headache_endstatus] | descriptive, Required<br>Field Annotation: @HIDEBUTTON |
| 461 | headache_pasc_adj | Did the physician state in the notes whether the | descriptive, Required |

| | | | |
|---|---|---|---|
| | Show the field ONLY if: [covid] = '1' and [diagnos is_post_covid(12)] = '1' | Headache was related to PASC?[headache_pasc] | Field Annotation: @HIDEBUTTON |
| 462 | headache_pasc_y_ad j<br><br>Show the field ONLY if: [covid] = '1' and [diagnos is_post_covid(12)] = '1' a nd [headache_pasc] = '1' | Copy and paste the language from the notes that states the Headache was related to PASC: [headache_pasc_y] | descriptive, Required<br>Field Annotation: @HIDEBUTTON |
| 463 | other_pasc_adj | Is there any additional information in the patient's chart about their PASC status that you think is relevant? If so, please enter below, ensuring that any PHI (names, MRNs, etc) is removed. [other_pasc] | descriptive |
| 464 | care_everywhere_ad j | Is there any additional information in Care Everywhere about this patient's PASC status? [care_everywhere] | descriptive, Required |
| 465 | optional_notes_adj | Optional- add any additional comments about this patient's chart. [optional_notes] | descriptive |
| 466 | long_covid_def | Section Header: *Long Covid Definition*<br><br>According its federal definition, Long COVID is defined as signs, symptoms, and conditions that continue or develop after initial COVID-19 or SARS-CoV-2 infection. The signs, symptoms, and conditions are present four weeks [28 days] or more after the initial phase of infection; may be multisystemic; and may present with a relapsing- remitting pattern and progression or worsening over time, with the possibility of severe and life-threatening events even months or years after infection. Long COVID is not one condition. It represents many potentially overlapping entities, likely with different biological causes and different sets of risk factors and outcomes<br><br>According to this definition, do you think this patient has Long COVID? | dropdown<br>1 Definitely<br>2 Probably<br>3 Possibly<br>4 No |
| 467 | long_covid_def_2 | According its federal definition, Long COVID is defined as signs, symptoms, and conditions that continue or develop after initial COVID-19 or SARS-CoV-2 infection. The signs, symptoms, and conditions are present four weeks [28 days] or more after the initial phase of infection; may be multisystemic; and may present with a relapsing- remitting pattern and progression or worsening over time, with the possibility of severe and life-threatening events even months or years after infection. Long COVID is not one condition. It represents many potentially overlapping entities, likely with different biological causes and different sets of risk factors and outcomes | dropdown<br>1 Definitely<br>2 Probably<br>3 Possibly<br>4 No |

| | | | |
|---|---|---|---|
| | | According to this definition, do you think this patient has Long COVID? | |
| 468 | long_covid_findings<br><br>Show the field ONLY if:<br>[long_covid_def] = '1' or [long_covid_def] = '2' or [long_covid_def] = '3' | If you answered definitely, probably, or possibly, list the findings that you think merit the Long Covid diagnosis. | notes |
| 469 | rules1 | Section Header: *Algorithm Rules to define PASC:*<br><br>Rules SARS-CoV-2 Infection Rule Fact of Infection Date of Infection Positive viral PCR Conclusive Yes Positive viral antigen Conclusive Yes Positive nucleocapsid serology Conclusive No Positive spike/nonspecific serology No No Specific COVID19 diagnosis Conclusive Yes (±) Complication COVID19 diagnosis Conclusive No? History COVID19 diagnosis Probable No Exposure COVID19 diagnosis No No Negative viral laboratory test Ruled out Yes | descriptive |
| 470 | rules2 | Plausible Medical Event Rule Attribution PASC, 2+ diagnoses Conclusive MIS-C, 2+ diagnoses Conclusive PASC, 1 diagnosis Probable MIS-C, 1 diagnosis Probable Post-viral sequela Probable Inflammatory disorder Probable Cardiac disorder, non-congenital Probable Respiratory disorder Possible Nonspecific disorder, enriched in COVID-19 patients Possible | descriptive |
| 471 | rules3 | Mechanism Attribution Fact references SARS-CoV-2 Conclusive Rare fact, enriched in COVID-19 Probable Fact attributed to rare cause¶, enriched in COVID-19 Probable Common fact§, enriched in COVID-19, following COVID-19 Probable Common fact§, enriched in COVID-19 Possible Negative viral testing Ruled out for 90 days ¶Rare Facts Include: Rheumatic conditions Non-stomatitis herpetic/chronic viral conditions - reactivation Non-murmur/non-congenital cardiac conditions - exclude silent conditions that are likely incidentally ascertained post-COVID-19 § Common facts include: Two or more diagnoses within the same cluster on different dates, each ≥28 days after the cohort entry date and separated by at least 28 days, of: Abdominal pain - new-onset only, washout after 2019-01-01 Abnormal liver enzymes Acute kidney injury Acute respiratory distress syndrome Arrythmias Cardiovascular signs and symptoms Changes in taste and smell Chest pain Cognitive function Fatigue and malaise Fever Fluid/electrolyte disturbance Generalized pain Hair loss Headache Heart disease (incl. pericarditis) Musculoskeletal Myocarditis Myositis Respiratory signs and symptoms Skin Thrombophlebitis/thromboembolism | descriptive |

| | | | |
|---|---|---|---|
| 472 | clinical_pasc_determination | Section Header: *Determination of PASC: Based on the information that was collected by the chart reviewer, use the above algorithm rules to determine whether this patient had PASC.*<br>Based on the algorithm logic, does this patient have PASC? | dropdown, Required<br>1 Conclusive- Results establish the presence of PASC with highest confidence<br>2 Probable- Results indicate the presence of PASC is significantly more likely than not<br>3 Possible- Results suggest that PASC may be present<br>4 No Evidence- Results provide no information about the presence of PASC<br>5 Ruled Out- Results indicate that PASC is likely not present |
| 473 | pasc_definite_clin_adj<br>Show the field ONLY if:<br>[clinical_pasc_determination] = '1' | Please explain why this patient definitely has PASC: | notes, Required |
| 474 | pasc_probable_clin_adj<br>Show the field ONLY if:<br>[clinical_pasc_determination] = '2' | Please explain why this patient probably has PASC: | notes, Required |
| 475 | pasc_possible_clin_adj<br>Show the field ONLY if:<br>[clinical_pasc_determination] = '3' | Please explain why this patient possibly has PASC: | notes, Required |
| 476 | pasc_negative_clin_adj<br>Show the field ONLY if:<br>[clinical_pasc_determination] = '4' | Please explain why this patient did not have PASC: | notes, Required |
| 477 | comments_clin_adj | Section Header: *Additional Comments*<br>Please add any additional comments. | notes |
| 478 | clinician_adjudication_complete | Section Header: *Form Status*<br>Complete? | dropdown<br>0 Incomplete<br>1 Unverified<br>2 Complete |

**Instrument: Algorithm comparison (algorithm_comparison)** **[collapsed]**
