## Supplemental Tables for "EHR-based Case Identification of Pediatric Long COVID: A Report from the RECOVER EHR Cohort"

**Supplemental Materials**

**Table S1.** Demographics of children with and without Long COVID based on the computable phenotype definition.

|  | **Chart Review Long COVID Positive (N=239)** | **Chart Review Long COVID Negative (N=412)** | **Overall (N=651)** |
| --- | --- | --- | --- |

| **Approx. CED age (years)** |  |  |  |
| --- | --- | --- | --- |
| Mean (SD) | 10.6 (6.04) | 9.82 (6.44) | 10.1 (6.30) |
| Median [Min, Max] | 11.6 [0, 21.0] | 10.3 [0.0700, 21.0] | 10.9 [0, 21.0] |
| **CED Age Group (years)** |  |  |  |
| <1 | 13 (5.4%) | 36 (8.7%) | 49 (7.5%) |
| 01 to 04 | 46 (19.2%) | 100 (24.3%) | 146 (22.4%) |
| 05 to 09 | 40 (16.7%) | 67 (16.3%) | 107 (16.4%) |
| 10 to 15 | 90 (37.7%) | 108 (26.2%) | 198 (30.4%) |
| 16 to 20 | 50 (20.9%) | 101 (24.5%) | 151 (23.2%) |
| **Patient Sex** |  |  |  |
| Male | 111 (46.4%) | 203 (49.3%) | 314 (48.2%) |
| Female | 128 (53.6%) | 209 (50.7%) | 337 (51.8%) |
| **Race** |  |  |  |
| Asian/Native Hawaiian/Pacific Islander | 8 (3.3%) | 18 (4.4%) | 26 (4.0%) |
| Black or African American | 38 (15.9%) | 77 (18.7%) | 115 (17.7%) |
| White | 137 (57.3%) | 220 (53.4%) | 357 (54.8%) |
| Multiple race | 7 (2.9%) | 14 (3.4%) | 21 (3.2%) |
| Other/Unknown | 49 (20.5%) | 83 (20.1%) | 132 (20.3%) |
| **Ethnicity** |  |  |  |
| Hispanic or Latino | 64 (26.8%) | 113 (27.4%) | 177 (27.2%) |
| Not Hispanic or Latino | 161 (67.4%) | 263 (63.8%) | 424 (65.1%) |
| Other/Unknown | 14 (5.9%) | 36 (8.7%) | 50 (7.7%) |
| **Payer** |  |  |  |
| Private/commercial | 125 (52.3%) | 157 (38.1%) | 282 (43.3%) |
| Public (Medicaid/SCHIP) | 93 (38.9%) | 165 (40.0%) | 258 (39.6%) |
| Other/Unknown | 21 (8.8%) | 90 (21.8%) | 111 (17.1%) |

*Note. CP = computable phenotype. *=at time of COVID-19 infection.*

**Table S2.** Statistics comparing CP and chart review identification of Long COVID presented separately for patients younger and older than 12 years of age.

|  | **Accuracy** | **Sensitivity** | **Specificity** | **PPV** | **NPV** | **F1** |
| --- | --- | --- | --- | --- | --- | --- |
| **Under 12 years of age**  **N = 367** | 0.602 | 0.619 | 0.593 | 0.443 | 0.749 | 0.517 |
| **12 years of age & older**  **N = 284** | 0.651 | 0.690 | 0.626 | 0.549 | 0.754 | 0.612 |

NPV = Negative predictive value. PPV = Positive predictive value

**Table S3.** Start and end dates associated with each era of infection.

| **Era** | **Start Date** | **End Date** |
| --- | --- | --- |
| Ancestral | Prior to 2020-09-30 | 2020-09-30 |
| Alpha | 2020-10-01 | 2021-05-30 |
| Delta | 2021-06-01 | 2021-11-30 |
| Omicron | 2021-11-30 | 2023-01-02 |

**Table S4.** Statistics comparing CP and chart review identification of Long COVID presented separately by era of infection.

|  | **Accuracy** | **Sensitivity** | **Specificity** | **PPV** | **NPV** | **F1** |
| --- | --- | --- | --- | --- | --- | --- |
| Ancestral  N = 43 | 0.558 | 0.562 | 0.556 | 0.429 | 0.682 | 0.486 |
| Alpha  N = 194 | 0.598 | 0.544 | 0.635 | 0.506 | 0.670 | 0.524 |
| Delta  N = 142 | 0.676 | 0.656 | 0.691 | 0.615 | 0.727 | 0.635 |
| Omicron  N = 272 | 0.625 | 0.771 | 0.561 | 0.435 | 0.848 | 0.557 |

NPV = Negative predictive value. PPV = Positive predictive value

**Table S5.** Statistics comparing CP and chart review identification of Long COVID presented separately by the number of clusters identified by the CP.

|  | Accuracy | Sensitivity | Specificity | PPV | NPV | F1 |
| --- | --- | --- | --- | --- | --- | --- |
| 0 Clusters  N = 236 | 0.801 | 0.228 | 0.983 | 0.812 | 0.800 | 0.356 |
| 1 Cluster  N = 209 | 0.469 | 0.611 | 0.394 | 0.346 | 0.659 | 0.442 |
| 2 Clusters  N = 99 | 0.495 | 0.791 | 0.268 | 0.453 | 0.625 | 0.576 |
| 3+ Clusters  N = 107 | 0.654 | 0.970 | 0.125 | 0.650 | 0.714 | 0.778 |

NPV = Negative predictive value. PPV = Positive predictive value.
